# Mpox clinical features and varicella-zoster virus coinfection in the Democratic Republic of Congo: a systematic review and meta-analysis (1970–2024)

**DOI:** 10.1101/2025.05.02.25326902

**Authors:** Fabrice Zobel Lekeumo Cheuyem, Constantine Tanywe Asahngwa, William Ndjidda Bakari, Chabeja Achangwa, Jessy Goupeyou-Youmsi, Brian Ngongheh Ajong, Claude Axel Minkandi, Mohamadou Adama, Jonathan Hangi Ndungo, Jude Tsafack Zefack, Solange Dabou, Badou Guianga, Saralees Nadarajah

**Affiliations:** Department of Public Health, Faculty of Medicine and Biomedical Sciences, The University of Yaoundé 1, Yaoundé, Cameroon; Division of Health Policy and Research, Nkafu Policy Institute, Denis and Lenora Foretia Foundation, Yaoundé, Cameroon; Periodontology Department, Faculty of Medicine, Pharmacy and Odontology, University Cheikh Anta Diop of Dakar, Dakar, Senegal; Department of Public Health, Faculty of Medical Sciences, University of West Indies, Cave Hill, Barbados; Health Emergencies Programme, World Health Organization (WHO), Kinshasa, Democratic Republic of Congo; National Multisectoral Programme to Fight against Maternal and Child Mortality, Ministry of Public Health, Yaoundé, Cameroon; Higher Institute of Medical Techniques, Bunia, Democratic Republic of Congo; Department of Family Health, Ministry of Public Health, Yaoundé, Cameroon; Social Epidemiology Lab, Wuppertal, Germany; Department of Global Health and Bioethics, Euclid University, Bangui, Central African Republic; Department of Mathematics, University of Manchester, Manchester, UK

**Author notes:** **Corresponding author’s address:** Fabrice Zobel Lekeumo Cheuyem, Department of Public Health, Faculty of Medicine & Biomedical Sciences, The University of Yaounde 1, Yaoundé, Cameroon.

**Keywords:** epidemiology, monkeypox, varicella-zoster virus, HIV, Clinical manifestation, Democratic Republic of Congo, systematic review, meta-analysis

## Abstract

**Background:** Monkeypox (Mpox) remains endemic in the Democratic Republic of Congo (DRC), with inc. Despite five decades of outbreaks, gaps persist in understanding clinical patterns and coinfections with varicella-zoster virus (VZV) and HIV in this high-burden setting.

**Methods:** We conducted a systematic review and meta-analysis (1970–2024) of Mpox cases in the DRC, extracting data from PubMed, ScienceDirect, and Google Scholar. Pooled prevalence rates were calculated using fixed and random-effects models, with subgroup analyses by time period, region, setting, study design and participant characteristics.

**Results:** Among 1,841 confirmed Mpox cases, VZV coinfection was 9.69% (95% CI: 1.33–18.06; n = 8), with higher rates in Kivu (33.33%) versus Equateur (11.10%). The VZV pooled prevalence rate among 64131 suspected Mpox cases was 16.73% (95% CI: 5.36-28.10; n = 8), with *I*^*2*^ = 99.4% (*p* □ 0.001). HIV coinfection was low (0.52%, 95% CI: 0.18–0.87) but elevated in South Kivu (1.64%). Among confirmed cases, rash (99.97%), painful lesions (78.17%), and Malaise (77.14%) dominated clinical presentation and underscored their diagnostic importance in the case definition. A similar clinical pattern of Mpox was observed among suspected cases, featuring a near-universal presentation of rash (99.43%) and fever (98.91%). Heterogeneity was high (*I*^*2*^ *> 90%*) for most outcomes.

**Conclusion:** Mpox in the DRC presents with consistent rash-centered symptomatology but high VZV coinfection rates complicate diagnosis. The low HIV coinfection suggests distinct transmission dynamics from global outbreaks. Findings underscore the need for integrated VZV/Mpox diagnostics and context-specific surveillance in endemic regions.

## Background

Monkeypox (Mpox), a zoonotic disease caused by the *Simian Orthopoxvirus*, has emerged as a significant public health concern, particularly in endemic regions of Central and West Africa [1,2]. The Democratic Republic of the Congo (DRC) remains the epicenter of human Mpox cases, where it has been responsible for 4.18% mortality (95% CI: 0.29-8.08) among confirmed cases since the first case was discovered in 1970 [3]. The Mpox virus belongs to the same genus as the smallpox virus (variola virus) and the virus used in the smallpox vaccine (vaccinia virus), known as the *Orthopoxvirus* genus. Scientists have identified two main types (clades) of the Mpox virus: Clade I (previously called the Congo Basin clade) and Clade II (previously called the West African clade). Clade I, which is common in the DRC, tends to cause more severe illness, has a higher mortality rate (up to 10%), and spreads more easily than Clade II. The recent global spread of a subtype of Clade II (Clade IIb) in 2022-2023, largely through sexual contact between people, demonstrated the virus’s ability to cause widespread outbreaks [4,5]. However, in the DRC, Mpox mainly spreads from animals to humans, with occasional human outbreaks linked to animals like rodents and other small mammals [6,7].

The clinical presentation of Mpox often resembles that of smallpox, though typically less severe. Classic symptoms include fever, lymphadenopathy, and a characteristic vesiculopustular rash that progresses through macular, papular, vesicular, and pustular stages before crusting over [8,9]. However, significant variability in clinical manifestations has been reported across different outbreaks and demographic groups [4,10–14]. This variability poses challenges for accurate clinical diagnosis, particularly in resource-limited settings where laboratory confirmation may be unavailable [15]. The situation is further complicated by the co-circulation of varicella-zoster virus (VZV), which produces similar cutaneous manifestations, leading to potential misdiagnosis and underreporting of Mpox cases [10,16,17]. Studies suggest that up to 40% of suspected Mpox cases in the DRC may actually represent VZV infections, underscoring the critical need for improved diagnostic capacity in endemic regions [4].

The emergence of the human immunodeficiency virus (HIV) as a comorbid condition in Mpox patients has introduced new complexities to the epidemiological landscape [18,19]. The extensive connectivity, coupled with frequent cross-border migration and commercial sex work along transit routes, facilitates regional transmission of diseases like Mpox and HIV [20]. Preliminary data suggest much lower HIV-Mpox coinfection rates (□4%) in this endemic setting, potentially reflecting distinct transmission dynamics [6]. Although the effects of HIV on the immune response to Clade I Mpox are not well-established, the weakened immune system associated with HIV likely increases the risk of severe illness and potential death in individuals with coinfection if they do not receive appropriate medical care [19,21]. Furthermore, the DRC’s strained healthcare infrastructure, ongoing conflict in eastern regions, and limited access to diagnostics create substantial barriers to accurate surveillance and case reporting [3,15].

Despite being endemic in the DRC for over five decades, critical gaps persist in our understanding of Mpox epidemiology in this region. The lack of standardized case definitions, inconsistent diagnostic approaches, and limited genomic surveillance have hindered accurate burden estimation and outbreak response. Recent advances in molecular diagnostics, including multiplex polymerase chain rection (PCR) assays capable of differentiating Mpox from VZV, offer new opportunities to improve case detection [10]. However, these tools often remain inaccessible in most endemic areas of the DRC [22]. These challenges can result in delayed diagnosis, inadequate treatment, and increased mortality from Mpox [23]. Additionally, the potential for Mpox virus evolution and adaptation to human-to-human transmission - as demonstrated by the 2022 global outbreak -underscores the urgent need for enhanced surveillance in endemic regions to detect any shifts in transmission patterns or clinical presentation [24].

To inform the development of more accurate case definitions tailored to the DRC context and provide evidence to guide clinical management and public health interventions in this high burden setting, there is a need to generate updated findings on the clinical and paraclinical presentation of the disease. The study is particularly timely given the recent expansion of Mpox vaccine trials in endemic countries and the need for data-driven strategies to optimize their deployment [25–28]. This systematic review and meta-analysis aimed to synthesize five decades (1970-2024) of evidence on Mpox clinical presentation and coinfection patterns in the DRC. By analyzing data from both confirmed and suspected cases, we sought to: (1) characterize the full spectrum of clinical manifestations associated with endemic Clade I Mpox, and (2) quantify the burden of VZV and HIV coinfections.

## Methods

### Study design

This systematic review and meta-analysis evaluated the clinical presentation and co-infection patterns of Mpox cases in the DRC between 1970 and 2024. The study was reported based on the Preferred Reporting Items for Systematic Review and Meta-analysis (PRISMA) guidelines [29].

### Study setting

The Democratic Republic of the Congo (DRC), the largest country in Sub-Saharan Africa, spans approximately 2,345,409 km^2^ [30]. It shares borders with nine countries (Angola, Burundi, Central African Republic, Republic of the Congo, Rwanda, South Sudan, Tanzania, Uganda, and Zambia) [20]. The DRC comprises 26 provinces, including Kinshasa (the capital), Nord-Kivu, Sud-Kivu, Equateur, Tshuapa, and Bas-Huélé among others [3]. The health system is structured into three tiers: central, intermediate, and operational levels, with 519 health zones, 393 general referral hospitals, and 8,504 health areas delivering primary care [31]. With a population exceeding 105 million, the DRC has a median age of 16.7 years, a growth rate of 3.1%, and a life expectancy of 61.6 years [32]. The country faces recurrent infectious disease outbreaks, including Mpox, Ebola, cholera, measles and yellow fever, exacerbated by humanitarian crises linked to protracted civil unrest and armed conflict [33]. Socio-culturally, over 93% of the population identifies as Christian, with Catholics representing 42% [34]. These demographic, health, and geopolitical factors collectively shape the DRC’s unique challenges in disease surveillance and outbreak response [35].

### Eligibility criteria

This systematic review and meta-analysis included all published studies reporting Mpox clinical manifestations and VZV coinfection in the DRC, irrespective of study design. We excluded studies with unclear outcome definitions and removed duplicate publications during screening. Only English-and French-language articles were eligible, with no restrictions on publication date.

### Article searching strategy

Published research was identified through a systematic search of digital repositories, including PubMed, ScienceDirect, and Google Scholar. The search strategy involved examining titles and abstracts using a combination of keywords and Medical Subject Headings (MeSH). Boolean operators (“AND” and “OR”) were employed to refine the search, with terms including (monkeypox) OR (monkeypox virus) OR (human monkeypox) OR (Mpox) OR (mpox) OR (MPX) OR (epidemiology) OR (surveillance) OR (characteristics) OR (clinical characteristics) OR (severe) AND (DRC) OR (Democratic Republic of Congo) OR (Zaire). A manual search for additional publications not indexed in these databases was conducted to ensure comprehensive coverage. Furthermore, the reference lists of included studies were screened for relevant articles. The final search was completed on February 27, 2025

### Data extraction

Data were extracted from all eligible articles using a pre-designed Microsoft Excel 2016 form to collect study characteristics. This form captured the first author’s name, study year, region, study design, study type, type of participant, setting, number of confirmed VZV cases, frequency of each clinical manifestation, and the number of suspected and confirmed Mpox cases. Two authors independently assessed the relevance and quality of each article. Any disagreements between the reviewers were resolved through discussion with a third author to reach a consensus

### Data quality assessment

The Joanna Briggs Institute (JBI) quality assessment tool was used to evaluate the quality of studies included [36]. Risk of bias was assessed using nine or ten criteria, depending on the study design. (1) For cross-sectional studies, criteria included: appropriateness of the sampling frame, use of a suitable sampling technique, adequate sample size, description of study subjects and setting, sufficient data analysis, use of valid methods for identifying conditions and measurements, use of appropriate statistical analysis, and an adequate response rate (≥60%). (2) For case series, criteria included: standardized measurement and valid identification of the condition for all participants, consecutive and complete inclusion of participants, reporting of participant demographics and clinical information, reporting of outcomes or follow-up results, reporting of the presenting site(s)/clinic(s) demographic information, and statistical analysis appropriate for case series studies. Each criterion was scored as 1 (yes) or 0 (no or unclear). The overall risk of bias was categorized as low (>50%), moderate (>25-50%), or high (≤25%).

### Outcome measurement

The primary outcomes of this systematic review and meta-analysis were clinical manifestations of Mpox cases and the coinfection rate of VZV and Mpox. Secondary outcomes included the HIV and Mpox coinfection rate and the prevalence of VZV among suspected Mpox cases. The VZV and Mpox coinfection rate was calculated by dividing the number of confirmed VZV cases by the number of confirmed Mpox cases. The prevalence of VZV among suspected Mpox cases was calculated by dividing the number of confirmed VZV cases by the total number of suspected Mpox cases. Similarly, the HIV and Mpox coinfection rate was assessed by dividing the number of confirmed HIV cases by the number of confirmed Mpox cases. For clinical features among suspected and confirmed Mpox cases, the proportion of each manifestation was determined by dividing its frequency by the number of suspected or confirmed Mpox cases, respectively.

### Operational definition

A suspected Mpox case was defined as an individual presenting with a vesicular or pustular rash characterized by deep-seated, firm pustules, and at least one of the following: fever preceding the rash, lymphadenopathy (inguinal, axillary, or cervical), or pustules or crusts on the palms or soles. A case was considered laboratory-confirmed Mpox if at least one specimen tested positive for *Orthopoxvirus* using a specific assay or Mpox-specific real-time PCR, or if Mpox was isolated in culture. A case was defined as laboratory-confirmed VZV if at least one specimen yielded a positive result in a real-time PCR assay targeting the VZV-specific DNA signature [10]. Similarly, a case was defined as laboratory-confirmed HIV if at least one specimen showed a positive result in an HIV antigen-specific assay or HIV-specific real-time PCR [5,37].

### Statistical analysis and synthesis

The *I*^*2*^ statistic was used to assess the heterogeneity between studies, categorizing heterogeneity as low (<25%), moderate (25-75%), or high (>75%). Subgroup analyses were then conducted based on study period, region, setting, participant type, study design, and disease burden, with a random-effects model employed when heterogeneity exceeded 50%. Furthermore, meta-regression explored whether study characteristics explained the variability in results. For the prevalence of VZV or HIV among suspected or confirmed Mpox cases, the time frames were defined based on major developments in healthcare systems: (1) 1970–1990: limited healthcare infrastructure in endemic regions; (2) 1991–2010: improvements in healthcare access and disease surveillance; (3) 2011–2024: strengthened global health initiatives and response systems [3]. Only study variables with meaningful and practical categories were included in the analyses. Univariable and multivariable meta-regression models were used to assess whether varied according to the selected explanatory variable categories. Statistical significance was set at a p-value of <0.05. All analyses were performed using the ‘meta’ package in R Statistics version 4.4.2 [38].

### Publication bias and sensitivity test

Publication bias was assessed visually using a funnel plot. The symmetry of the inverted funnel shape suggested the absence of publication bias. The trim-and-fill method was used to adjust for potential missing studies [39]. Sensitivity analysis was conducted by iteratively excluding one study at a time to assess the robustness of the findings.

## Results

The database search yielded 157 records, and an additional 4 were identified from other sources, resulting in a total of 161 records. After removing 9 duplicates, 151 unique records underwent title and abstract screening, followed by full-text assessment for eligibility. Ultimately, 20 study reports met the inclusion criteria and were included in the meta-analysis (Fig. 1).

**Fig. 1.**
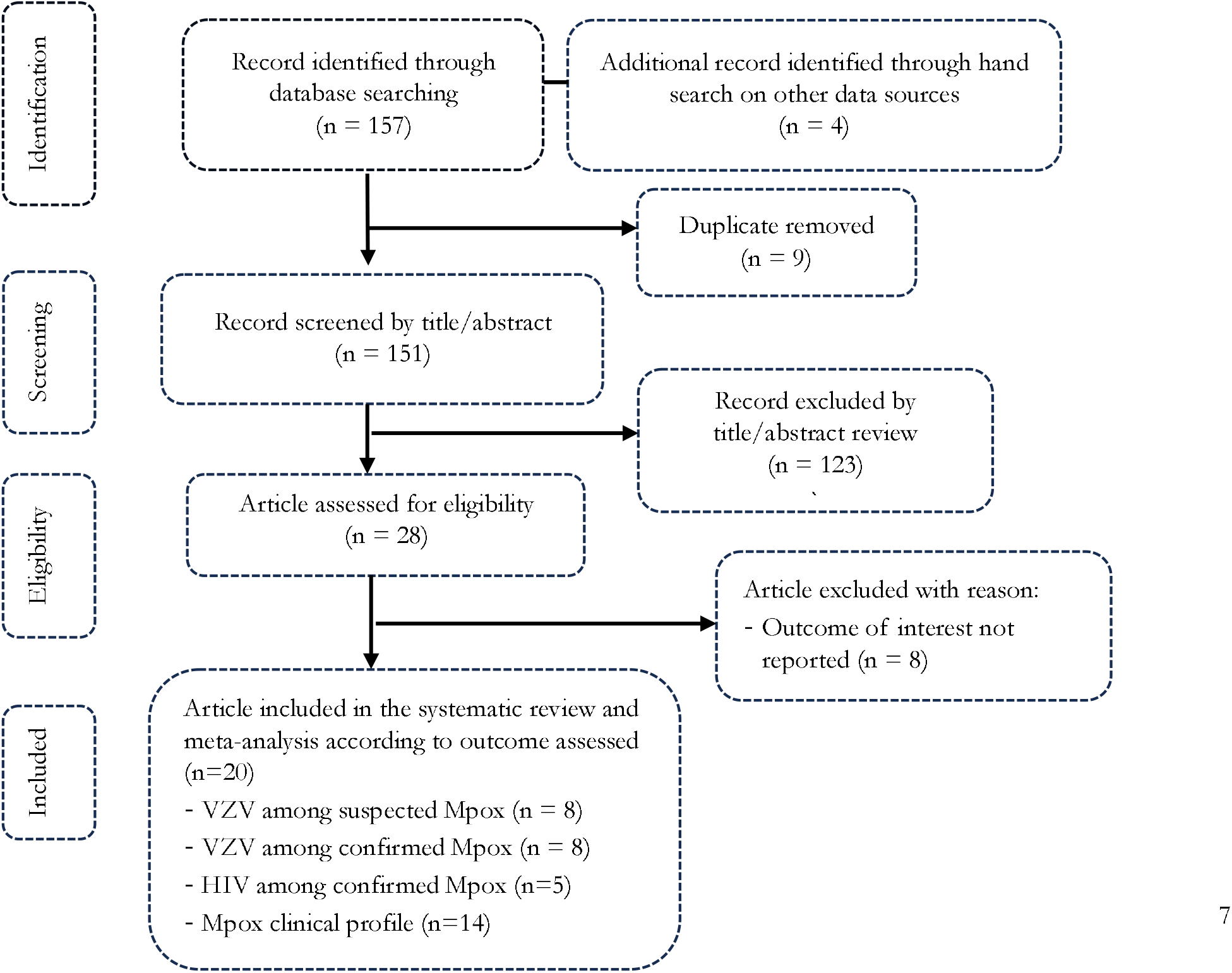
PRISMA diagram flow of studies included in the meta-analysis (VZV: Varicella-zona virus; HIV: Human immunodeficiency virus)

### Studies selection

### Characteristics of studies included

Twenty studies, conducted between 1970 and 2024 in community and hospital settings across the DRC, were included in this comprehensive analysis. Data collection in most (n = 19) utilized surveillance and investigation, targeting the general population and healthcare workers to describe VZV, HIV, and Mpox coinfection and the clinical features of suspected and confirmed Mpox cases (Additional Files 1, Supplementary Table 1)

### Varicella zona prevalence among suspected Mpox cases

The national pooled prevalence rate of the VZV cases among 64131 suspected Mpox cases was 16.73% (95% CI: 5.36-28.10; n = 8), with *I*^2^ = 99.4% (*p* ⍰ 0.001) (Fig. 2).

**Fig. 2.**
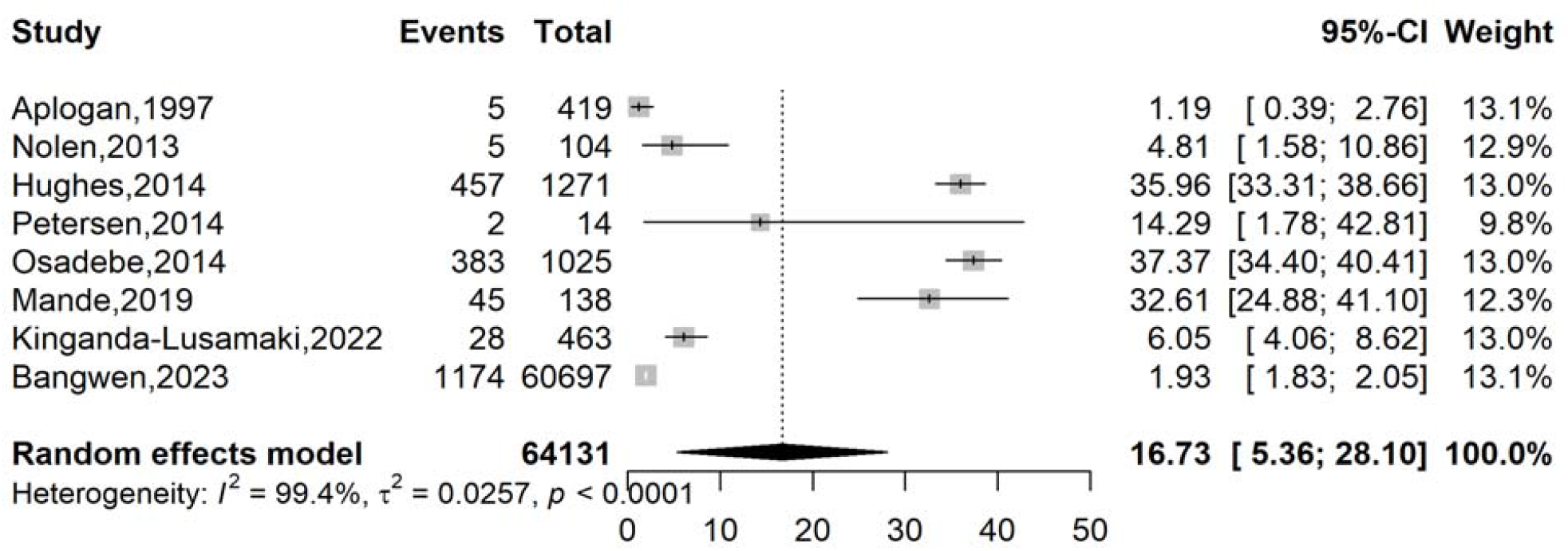
Pooled estimate of the prevalence rate of varicella-zoster virus isolated among suspected Mpox cases in DRC, 1970-2024

The subgroup analysis reveals a notable increase in the suspected Mpox event rate from 1.19% (95% CI: 0.39-2.76; n = 1 study) in 1991-2010 to 19.06% (95% CI: 6.96-31.17; n = 7 studies) in 2011-2024. Substantial heterogeneity (*p* □ 0.001) existed across most subgroups, indicating considerable variation in event rates between studies. Regionally, the highest proportion was observed in the Northeastern region (32.61%; 95% CI: 24.88-41.10; n = 1 study), and among healthcare workers (14.29%; 95% CI: 1.78-42.81; n = 1 study), while setting and participant type also demonstrated varying event rates (Table 1 and Additional Files 2, Supplementary Fig. 1-5).

**Table 1.** Subgroup meta-analysis of varicella-zoster virus pooled estimate proportion among Mpox suspected case in DRC, 1970-2024.

| Subgroup | Suspected Mpox cases | Event rate<sup>1</sup> (%) | 95% CI limits<sup>1</sup> |  | Number of studies | Heterogeneity statistic<sup>1</sup> |  |
| --- | --- | --- | --- | --- | --- | --- | --- |
| | | | Lower | Upper | | $I^2$ (%) | $p$-value |
| **Study period<sup>2</sup>** |  |  |  |  |  |  |  |
| 1991-2010 | 419 | 1.19 | 0.39 | 2.76 | 1 | - | - |
| 2011-2024 | 63712 | 19.06 | 6.96 | 31.17 | 7 | 99.5 | □0.001 |
| **Region<sup>3</sup>** |  |  |  |  |  |  |  |
| Northeastern | 138 | 32.61 | 24.88 | 41.10 | 1 | - | - |
| Central/Western | 2833 | 18.93 | 3.31 | 34.54 | 5 | 99.6 | □0.001 |
| Nationwide | 61160 | 3.84 | 0.00 | 7.86 | 2 | 92.7 | □0.001 |
| **Setting** |  |  |  |  |  |  |  |
| Community | 3434 | 18.96 | 6.75 | 31.16 | 7 | 99.4 | □0.001 |
| Online | 60697 | 1.93 | 1.83 | 28.10 | 1 | - | - |
| **Participant** |  |  |  |  |  |  |  |
| General population | 64013 | 19.05 | 4.76 | 33.33 | 6 | 99.6 | □0.001 |
| General population and HCWs | 104 | 4.81 | 1.58 | 10.86 | 1 | - | - |
| HCWs | 14 | 14.29 | 1.78 | 42.81 | 1 | - | - |
| **Disease burden (suspected Mpox cases)** |  |  |  |  |  |  |  |
| □400 | 256 | 17.25 | 0.00 | 34.69 | 3 | 94.8 | □0.001 |
| ≥400 | 63875 | 16.46 | 0.25 | 32.67 | 5 | 99.7 | □0.001 |

<sup>1</sup> Random effects model; CI: Confidence Interval; <sup>2</sup> 1970–1990: Limited healthcare infrastructure in endemic regions; 1991–2010: Improvements in healthcare access and disease surveillance; 2011–2024: Strengthened global health initiatives and response systems; <sup>3</sup> Northeastern=Bas-Uélé, Central/Western= Tshuapa and Kasai oriental; HCW: Healthcare Worker

### Varicella zona virus and Mpox coinfection

A total of 1841 confirmed Mpox cases were examined, with the pooled coinfection rate of VZV estimated at 9.69% (95% CI: 1.33-18.06; n = 8 studies) in the country. Significant heterogeneity was observed across these studies (*I*^*2*^ = 96.9%, *p* < 0.001) (Fig. 3).

**Fig. 3.**
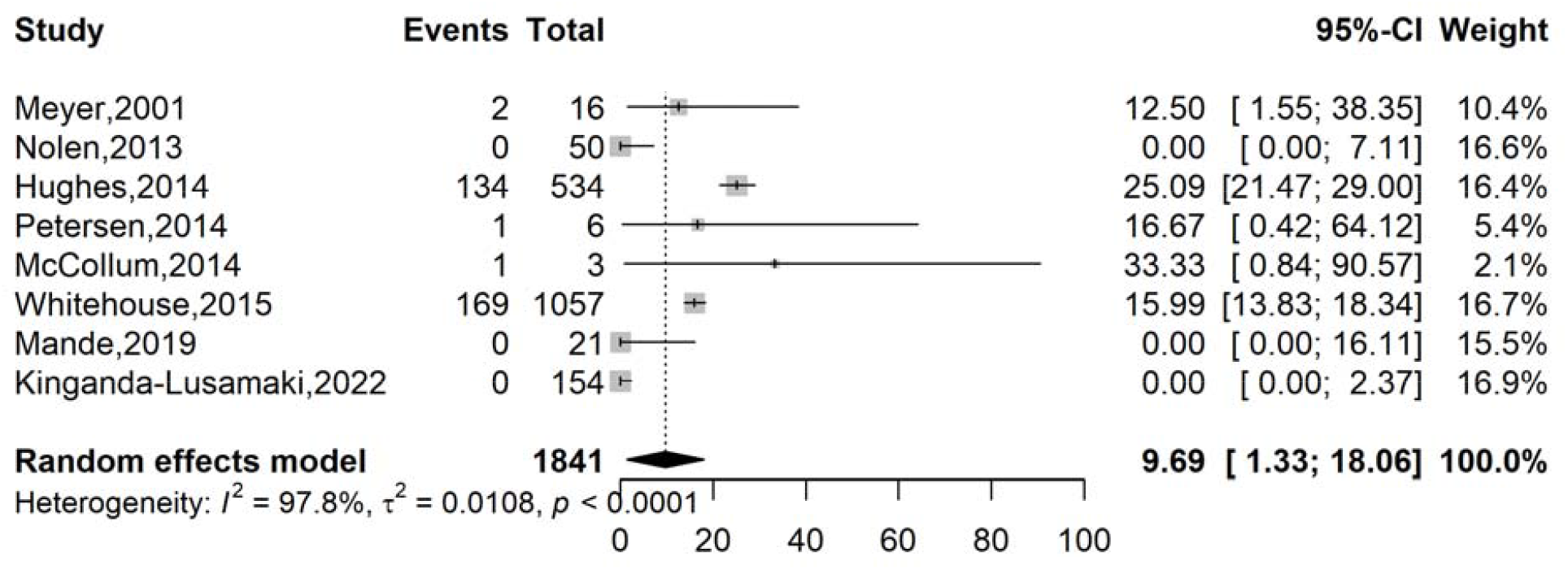
Pooled estimate of the coinfection proportion of varicella-zoster virus among confirmed Mpox cases in DRC, 1970-2024

The subgroup analysis of coinfection cases indicates relatively stable event rates across the study periods of 1991-2010 (12.50%; 95% CI: 1.55-38.35; n = 1 study) and 2011-2024 (9.47%; 95% CI: 0.11-18.82; n = 6), although the latter period benefits from more extensive data (n = 7 studies). Significant heterogeneity (*p* < 0.001) was prevalent across several subgroups, suggesting considerable variability in event rates between studies. Regionally, Equateur reported an event rate of 11.10%; (95% CI : 1.69-20.51; n = 6) while Kivu regions showed a higher rate (33.33%; 95% CI : 0.84-90.7) based on limited data (n = 1). Similarly, participant type and disease burden categories exhibited varying event rates, often accompanied by substantial heterogeneity (Table 2 and Additional Files 3, Fig.1-5).

**Table 2.** Subgroup meta-analysis of the pooled estimate proportion varicella-zoster virus and Mpox coinfection in DRC, 1970-2024.

| Subgroup | Confirmed Mpox cases | Event rate<sup>1</sup> (%) | 95% CI limits<sup>1</sup> |  | Number of studies | Heterogeneity statistic<sup>1</sup> |  |
| --- | --- | --- | --- | --- | --- | --- | --- |
|  |  |  | Lower | Upper |  | I<sup>2</sup>(%) | p-value |
| **Study period<sup>2</sup>** |  |  |  |  |  |  |  |
| 1991-2010 | 16 | 12.50 | 1.55 | 38.35 | 1 | - | - |
| 2011-2024 | 1825 | 9.47 | 0.11 | 18.82 | 7 | 98.1 | □0.001 |
| **Region<sup>3</sup>** |  |  |  |  |  |  |  |
| Equateur | 1684 | 11.10 | 1.69 | 20.51 | 6 | 96.7 | □0.001 |
| Kivu | 3 | 33.33 | 0.84 | 90.7 | 1 | - | - |
| Nationwide | 154 | 0.00 | 0.00 | 2.37 | 1 | - | - |
| **Participant** |  |  |  |  |  |  |  |
| General population | 1785 | 11.33 | 1.42 | 21.24 | 6 | 98.4 | □0.001 |
| General population and HCWs | 50 | 0 | 0.00 | 7.11 | 1 | - | - |
| HCWs | 6 | 16.67 | 0.42 | 64.12 | 1 | - | - |
| **Disease burden (confirmed cases)** |  |  |  |  |  |  |  |
| □40 | 46 | 2.51 | 0.00 | 8.19 | 4 | 29.1* | 0.237 |
| ≥40 | 1795 | 10.21 | 0.00 | 22.35 | 4 | 99.1 | □0.001 |

<sup>1</sup>Random effects model; CI: Confidence interval; <sup>2</sup>1970–1990: Limited healthcare infrastructure in endemic regions; 1991-2010: Improvements in healthcare access and disease surveillance; 2011-2024: Strengthened global health initiatives and response systems; <sup>3</sup> Equateur = Equateur, Tshuapa and Bas-Uélé; Kivu = North and South Kivu; \* Fixed effect model applied

### HIV and Mpox coinfection

The HIV coinfection rate among 1652 confirmed Mpox cases was 0.52% (95% CI: 0.18-0.87) with a low heterogeneity (*I*^2^ = 39.2%, *p* = 0.160) between studies included (n = 5) (Fig. 4).

**Fig. 4.**
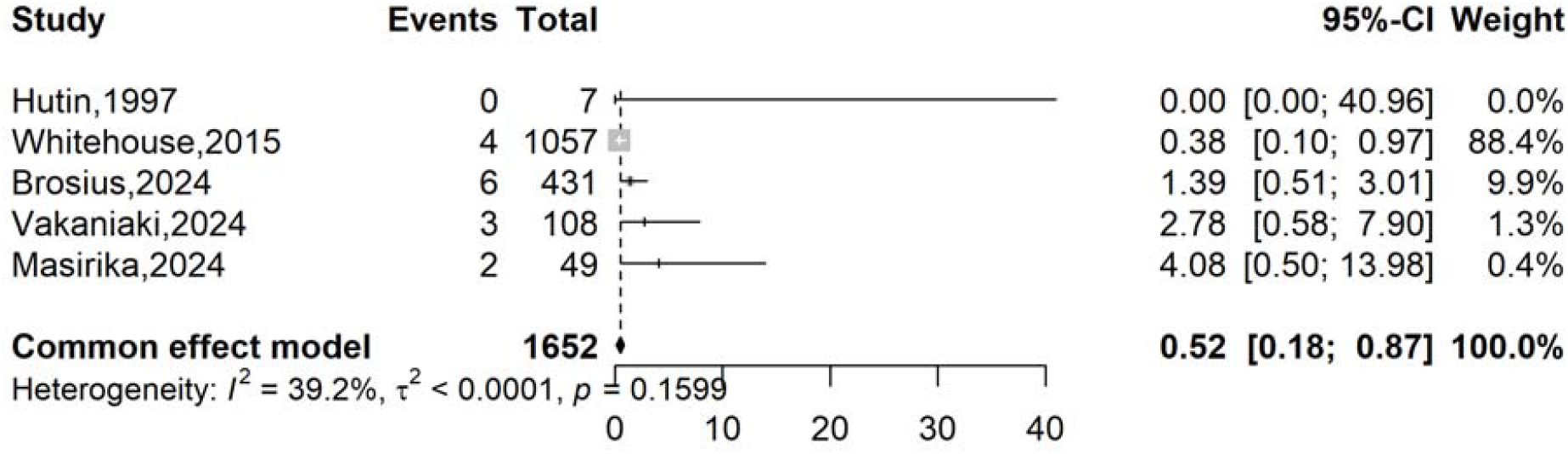
Pooled estimate of the coinfection proportion of HIV among confirmed Mpox cases in DRC, 1970-2024

The meta-analysis of HIV and Mpox coinfection in the DRC indicates a generally low pooled proportion, with a slight increase observed in 2011-2024 (0.52%; 95% CI: 0.18-0.87; n = 4 studies) compared to earlier (0.00%; 95% CI: 0.00-40.96; n = 1 study). Regional variations showed a higher proportion in South Kivu (1.64%; 95% CI: 0.61-2.66; n = 3 studies). Furthermore, a higher coinfection proportion was found in settings with a lower Mpox burden (3.01%; 95% CI: 0.34-5.68; n = 3 studies) (Table 3, Additional Files 4, Supplementary Fig. 1-5).

**Table 3.** Subgroup meta-analysis of the pooled estimate proportion of HIV and Mpox coinfection in DRC, 1970-2024.

| Subgroup | Confirmed cases | Event rate<sup>1</sup> (%) | 95% CI limits<sup>1</sup> |  | Number of studies | Heterogeneity statistic<sup>1</sup> |  |
| --- | --- | --- | --- | --- | --- | --- | --- |
|  |  |  | Lower | Upper |  | I<sup>2</sup>(%) | p-value |
| **Study period<sup>2</sup>** |  |  |  |  |  |  |  |
| 1991-2010 | 7 | 0.00 | 0.00 | 40.96 | 1 | - | - |
| 2011-2024 | 1645 | 0.52 | 0.18 | 0.87 | 4 | 54.4 | 0.087 |
| **Region<sup>3</sup>** |  |  |  |  |  |  |  |
| South Kivu | 588 | 1.64 | 0.61 | 2.66 | 3 | 0.0 | 0.483 |
| Other | 1064 | 0.38 | 0.01 | 0.75 | 2 | 0.0 | 0.965 |
| **Study design** |  |  |  |  |  |  |  |
| Cross-sectional | 431 | 1.39 | 0.51 | 3.01 | 1 | - | - |
| Surveillance and Investigation | 1221 | 0.43 | 0.06 | 0.79 | 4 | 24.1 | 0.267 |
| **Study setting** |  |  |  |  |  |  |  |
| Community | 1221 | 0.43 | 0.06 | 0.79 | 4 | 24.1 | 0.267 |
| Hospital | 431 | 1.39 | 0.51 | 3.01 | 1 | - | - |
| **Disease burden (confirmed cases)** |  |  |  |  |  |  |  |
| □110 | 164 | 3.01 | 0.34 | 5.68 | 3 | 0.0 | 0.866 |
| ≥110 | 1488 | 0.48 | 0.13 | 0.83 | 2 | 65.5 | 0.089 |

<sup>1</sup> Fixed effects model; CI: Confidence interval; <sup>2</sup>1970–1990: Limited healthcare infrastructure in endemic regions; 1991-2010: Improvements in healthcare access and disease surveillance; 2011-2024: Strengthened global health initiatives and response systems; <sup>3</sup> Equateur: Equateur, Tshuapa and Bas-Uélé; Kivu: North and South Kivu;

### Subgroup analysis

### Mpox clinical profile

This comprehensive analysis of confirmed Mpox cases in the DRC reveals distinct clinical patterns, with rash being the most universal manifestation (99.97%; 95% CI: 99.85-100.00%), followed by painful lesions (78.17%) and malaise (77.14%). Notably, systemic symptoms such as fever (67.94%) and lymphadenopathy (71.99%) were also prevalent, while genital lesions (60.97%) and oral lesions (44.22%) demonstrated substantial occurrence. The data shows high heterogeneity (*I*^*2*^ > 90% for most symptoms, *p* < 0.001), reflecting variability across studies. Severe manifestations like convulsions (0.23%), hemorrhagic lesions (2.78%), and hypotension (1.39%) were rare but documented (Table 4, Fig. 5 and Additional Files 5, Supplementary Fig.1).

**Table 4.**
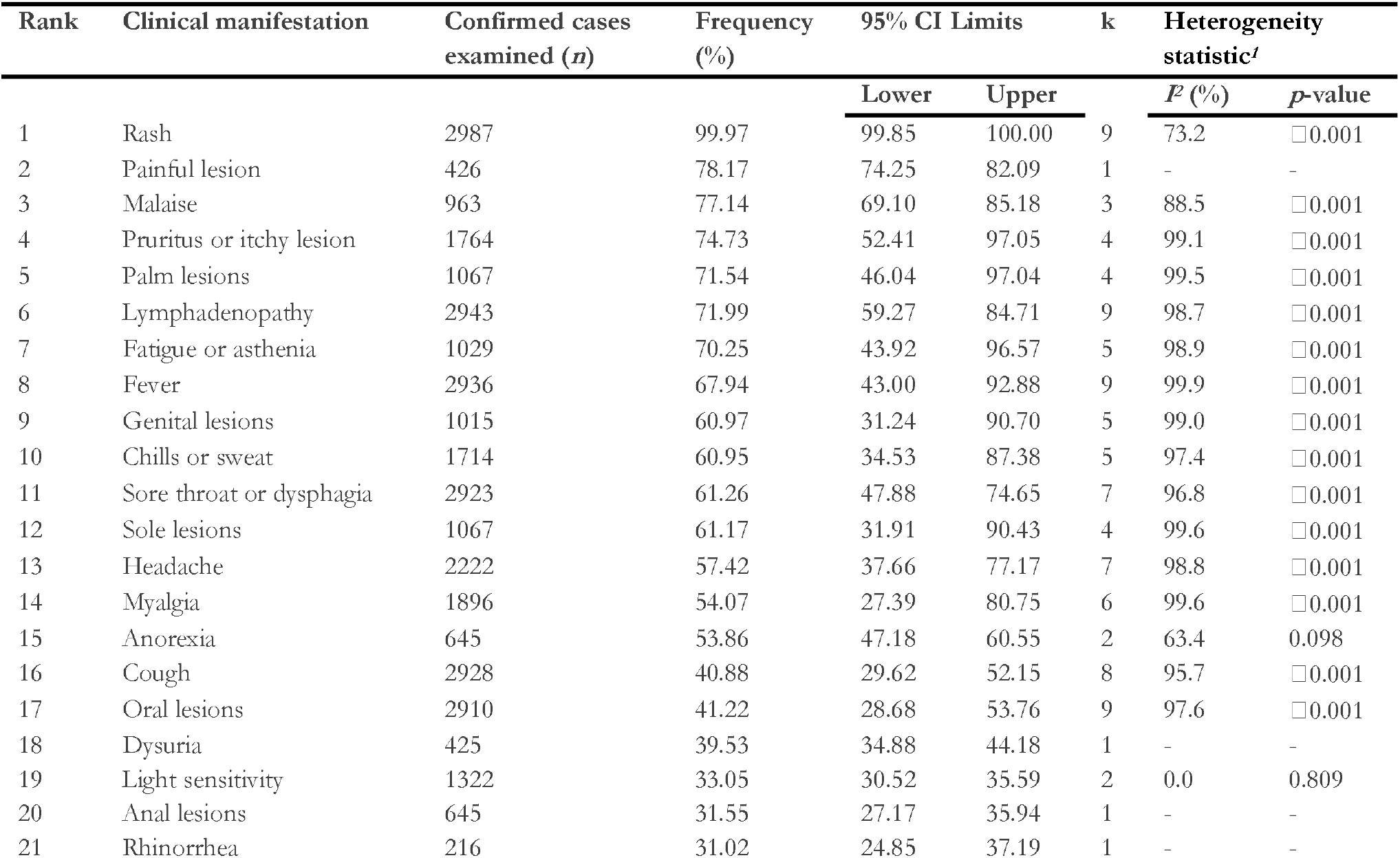
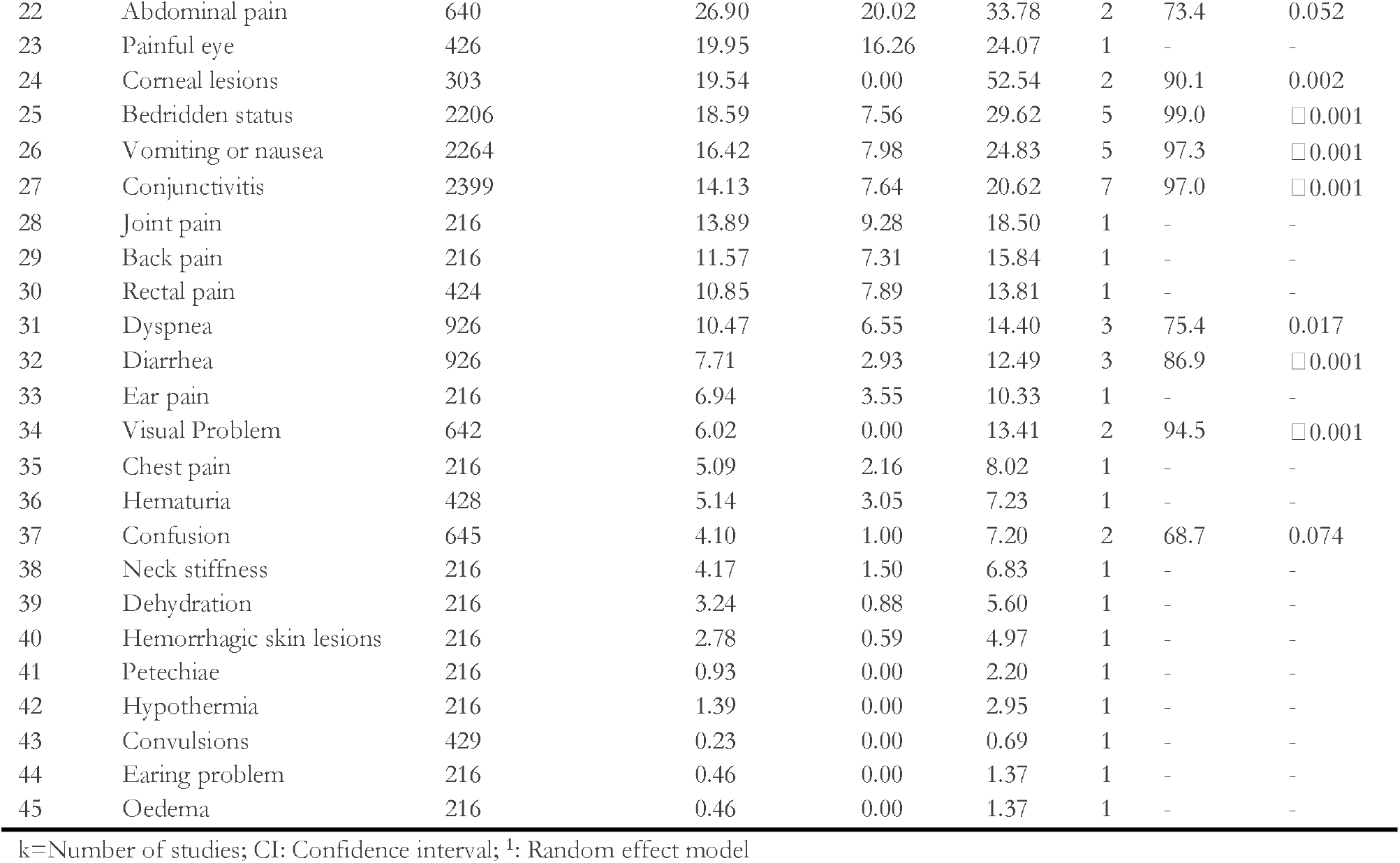
Clinical pattern of confirmed Mpox cases in DRC, 1970-2024.

| Rank | Clinical manifestation | Confirmed cases examined (*n*) | Frequency (%) | 95% CI Limits |  | k | Heterogeneity statistic<sup>†</sup> |  |
| --- | --- | --- | --- | --- | --- | --- | --- | --- |
| | | | | Lower | Upper | | $I^2$ (%) | *p*-value |
| 1 | Rash | 2987 | 99.97 | 99.85 | 100.00 | 9 | 73.2 | □ 0.001 |
| 2 | Painful lesion | 426 | 78.17 | 74.25 | 82.09 | 1 | - | - |
| 3 | Malaise | 963 | 77.14 | 69.10 | 85.18 | 3 | 88.5 | □ 0.001 |
| 4 | Pruritus or itchy lesion | 1764 | 74.73 | 52.41 | 97.05 | 4 | 99.1 | □ 0.001 |
| 5 | Palm lesions | 1067 | 71.54 | 46.04 | 97.04 | 4 | 99.5 | □ 0.001 |
| 6 | Lymphadenopathy | 2943 | 71.99 | 59.27 | 84.71 | 9 | 98.7 | □ 0.001 |
| 7 | Fatigue or asthenia | 1029 | 70.25 | 43.92 | 96.57 | 5 | 98.9 | □ 0.001 |
| 8 | Fever | 2936 | 67.94 | 43.00 | 92.88 | 9 | 99.9 | □ 0.001 |
| 9 | Genital lesions | 1015 | 60.97 | 31.24 | 90.70 | 5 | 99.0 | □ 0.001 |
| 10 | Chills or sweat | 1714 | 60.95 | 34.53 | 87.38 | 5 | 97.4 | □ 0.001 |
| 11 | Sore throat or dysphagia | 2923 | 61.26 | 47.88 | 74.65 | 7 | 96.8 | □ 0.001 |
| 12 | Sole lesions | 1067 | 61.17 | 31.91 | 90.43 | 4 | 99.6 | □ 0.001 |
| 13 | Headache | 2222 | 57.42 | 37.66 | 77.17 | 7 | 98.8 | □ 0.001 |
| 14 | Myalgia | 1896 | 54.07 | 27.39 | 80.75 | 6 | 99.6 | □ 0.001 |
| 15 | Anorexia | 645 | 53.86 | 47.18 | 60.55 | 2 | 63.4 | 0.098 |
| 16 | Cough | 2928 | 40.88 | 29.62 | 52.15 | 8 | 95.7 | □ 0.001 |
| 17 | Oral lesions | 2910 | 41.22 | 28.68 | 53.76 | 9 | 97.6 | □ 0.001 |
| 18 | Dysuria | 425 | 39.53 | 34.88 | 44.18 | 1 | - | - |
| 19 | Light sensitivity | 1322 | 33.05 | 30.52 | 35.59 | 2 | 0.0 | 0.809 |
| 20 | Anal lesions | 645 | 31.55 | 27.17 | 35.94 | 1 | - | - |
| 21 | Rhinorrhea | 216 | 31.02 | 24.85 | 37.19 | 1 | - | - |
| 22 | Abdominal pain | 640 | 26.90 | 20.02 | 33.78 | 2 | 73.4 | 0.052 |
| 23 | Painful eye | 426 | 19.95 | 16.26 | 24.07 | 1 | - | - |
| 24 | Corneal lesions | 303 | 19.54 | 0.00 | 52.54 | 2 | 90.1 | 0.002 |
| 25 | Bedridden status | 2206 | 18.59 | 7.56 | 29.62 | 5 | 99.0 | □ 0.001 |
| 26 | Vomiting or nausea | 2264 | 16.42 | 7.98 | 24.83 | 5 | 97.3 | □ 0.001 |
| 27 | Conjunctivitis | 2399 | 14.13 | 7.64 | 20.62 | 7 | 97.0 | □ 0.001 |
| 28 | Joint pain | 216 | 13.89 | 9.28 | 18.50 | 1 | - | - |
| 29 | Back pain | 216 | 11.57 | 7.31 | 15.84 | 1 | - | - |
| 30 | Rectal pain | 424 | 10.85 | 7.89 | 13.81 | 1 | - | - |
| 31 | Dyspnea | 926 | 10.47 | 6.55 | 14.40 | 3 | 75.4 | 0.017 |
| 32 | Diarrhea | 926 | 7.71 | 2.93 | 12.49 | 3 | 86.9 | □ 0.001 |
| 33 | Ear pain | 216 | 6.94 | 3.55 | 10.33 | 1 | - | - |
| 34 | Visual Problem | 642 | 6.02 | 0.00 | 13.41 | 2 | 94.5 | □ 0.001 |
| 35 | Chest pain | 216 | 5.09 | 2.16 | 8.02 | 1 | - | - |
| 36 | Hematuria | 428 | 5.14 | 3.05 | 7.23 | 1 | - | - |
| 37 | Confusion | 645 | 4.10 | 1.00 | 7.20 | 2 | 68.7 | 0.074 |
| 38 | Neck stiffness | 216 | 4.17 | 1.50 | 6.83 | 1 | - | - |
| 39 | Dehydration | 216 | 3.24 | 0.88 | 5.60 | 1 | - | - |
| 40 | Hemorrhagic skin lesions | 216 | 2.78 | 0.59 | 4.97 | 1 | - | - |
| 41 | Petechiae | 216 | 0.93 | 0.00 | 2.20 | 1 | - | - |
| 42 | Hypothermia | 216 | 1.39 | 0.00 | 2.95 | 1 | - | - |
| 43 | Convulsions | 429 | 0.23 | 0.00 | 0.69 | 1 | - | - |
| 44 | Earing problem | 216 | 0.46 | 0.00 | 1.37 | 1 | - | - |
| 45 | Oedema | 216 | 0.46 | 0.00 | 1.37 | 1 | - | - |

k=Number of studies; CI: Confidence interval; <sup>1</sup>: Random effect model

**Fig. 5.**
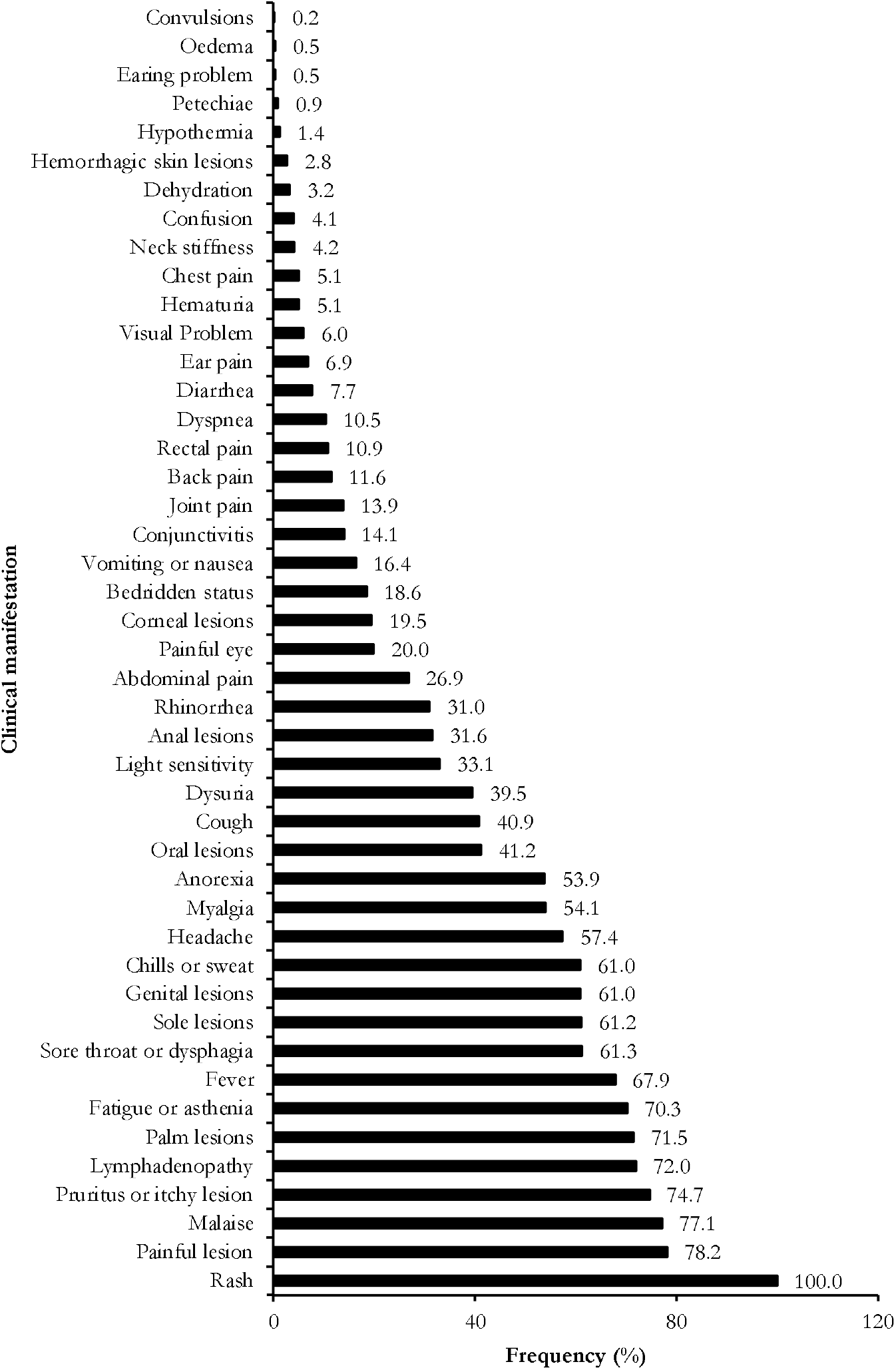
Trend of clinical manifestations observed among confirmed Mpox cases in DRC,1970-2024

An almost similar clinical pattern of Mpox was observed among suspected cases, featuring a near-universal presentation of rash (99.43%, 95% CI: 98.92–99.94) and fever (98.91%, 95% CI: 98.32–99.50). Painful lesions (91.17%) and lymphadenopathy (68.93%) were also prevalent, while systemic symptoms such as fatigue (58.11%) and myalgia (49.49%) occurred moderately. Notably, conjunctivitis (9.92%) and diarrhea (10.91%) were infrequent but non-negligible. Heterogeneity was high for most symptoms (*I*^*2*^ > 90%, *p* < 0.001). Rare but severe features (e.g., bedridden status: 18.44%) and demographic-specific patterns (e.g., genital lesions: 34.00%) were also described (Table 5, Fig. 6 and Additional Files 6, Supplementary Fig. 1).

**Table 5.** Clinical pattern of suspected Mpox cases in DRC, 1970-2024.

| Rank | Clinical manifestation | Suspected cases examined (*n*) | Frequency (%) | 95% CI |  | k | Heterogeneity statistic<sup>1</sup> |  |
| --- | --- | --- | --- | --- | --- | --- | --- | --- |
|  |  |  |  | Lower | Upper |  | *I*<sup>2</sup> (%)<sup>a</sup> | *p*-value |
| 1 | Rash | 1120 | 99.43 | 98.92 | 99.94 | 5 | 89.2 | <0.001 |
| 2 | Fever | 1035 | 98.91 | 98.32 | 99.50 | 5 | 97.5 | <0.001 |
| 3 | Palm lesions | 749 | 91.17 | 89.14 | 93.20 | 2 | 25.4 | 0.265 |
| 4 | Conjunctivitis | 1031 | 9.92 | 8.14 | 11.69 | 3 | 96.6 | <0.001 |
| 5 | Sole lesions | 750 | 81.22 | 78.43 | 84.02 | 2 | 0.0 | 0.564 |
| 6 | Chills or sweat | 795 | 78.86 | 76.04 | 81.69 | 2 | 86.3 | 0.007 |
| 7 | Diarrhea | 339 | 10.91 | 7.60 | 14.23 | 1 | - | - |
| 8 | Lymphadenopathy | 1440 | 68.93 | 66.63 | 71.23 | 5 | 96.5 | <0.001 |
| 9 | Malaise | 720 | 62.92 | 59.39 | 66.44 | 1 | - | - |
| 10 | Fatigue | 1032 | 58.11 | 55.77 | 60.45 | 3 | 99.7 | <0.001 |
| 11 | Sore throat or dysphagia | 1369 | 53.40 | 50.89 | 55.91 | 4 | 98.0 | <0.001 |
| 12 | Pruritus or itchy lesion | 734 | 51.63 | 48.02 | 55.25 | 1 | - | - |
| 13 | Myalgia | 292 | 49.49 | 47.12 | 51.85 | 3 | 99.7 | <0.001 |
| 14 | Cough | 1366 | 37.13 | 34.65 | 39.62 | 4 | 96.5 | <0.001 |
| 15 | Genital lesions | 877 | 34.00 | 31.35 | 36.65 | 3 | 99.4 | <0.001 |
| 16 | Oral lesions | 1372 | 32.83 | 30.53 | 35.14 | 4 | 98.7 | <0.001 |
| 17 | Light sensitivity | 743 | 27.16 | 23.93 | 30.39 | 1 | - | - |
| 18 | Back pain | 51 | 31.37 | 18.64 | 44.11 | 1 | - | - |
| 19 | Vomiting or nausea | 788 | 20.15 | 17.37 | 22.92 | 2 | 93.5 | <0.001 |
| 20 | Bedridden status | 743 | 18.44 | 15.65 | 21.23 | 1 | - | - |
| 21 | Headache | 1021 | 52.40 | 10.40 | 94.37 | 3 | 99.7 | <0.001 |
| 22 | Rhinorrhea | 51 | 21.57 | 10.28 | 32.86 | 1 | - | - |
| 23 | Abdominal pain | 3 | 33.33 | 0.00 | 86.68 | 1 | - | - |
| 24 | Alopecia | 88 | 3.41 | 0.00 | 7.20 | 1 | - | - |

k=Number of studies; CI: Confidence interval; <sup>1</sup>: Random effect model

**Table 6.** Meta-regression to explore sources of heterogeneity in the pooled estimate of varicella-zoster virus infection among suspected and confirmed Mpox cases in the DRC, 1970-2024.

| Epidemiological estimate | Moderator | Univariate |  | Multivariate |  |
| --- | --- | --- | --- | --- | --- |
| | | Unadjusted coefficient ($\beta$) | *p*-value | Adjusted coefficient ($\beta$) | *p*-value |
| VZV among suspected Mpox cases |  |  |  |  |  |
| Study year | Years | 0.0532 | 0.489 | 0.2247 | <0.001 |
| Region | Regional vs. Nationwide | -1.5582 | 0.192 | 4.7148 | <0.001 |
| Participant | General population vs. Other | 0.2986 | 0.833 | 0.5377 | 0.577 |
| Disease burden<sup>†</sup> | □400 vs. ≥400 | 0.5960 | 0.627 | -1.2899 | 0.164 |
| VZV among confirmed Mpox cases |  |  |  |  |  |
| Study year | Years | -0.1049 | 0.293 | 0.0046 | 0.959 |
| Region | Regional vs. Nationwide | 4.1257 | 0.006 | 4.2220 | 0.018 |
| Participant | General population vs. Other | 0.8139 | 0.554 | 1.2466 | 0.266 |
| Disease burden | □40 vs. ≥40 | 0.7141 | 0.5589 | 0.0275 | 0.976 |
| HIV among confirmed Mpox cases |  |  |  |  |  |
| Study year | Years | -0.0030 | 0.964 | -0.1592 | 0.063 |
| Region | South Kivu vs. Others | 1.3090 | 0.087 | 3.608 | 0.002 |
| Study design<sup>†</sup> | SIR vs. Cross-sectional | 0.3004 | 0.826 | - | - |
| Setting | Hospital vs. Community | -0.3004 | 0.826 | -0.863 | 0.159 |
| Disease burden<sup>†</sup> | □110 vs. ≥110 | 1.5627 | 0.033 | - | - |

<sup>†</sup> Redundant predictor dropped from the model; VZV: Varicella-zona virus; HIV: Human immunodeficiency virus; Others included Sankuru and Tshuapa Regions; SIR: Surveillance and investigation report (Case series)

**Fig. 6.**
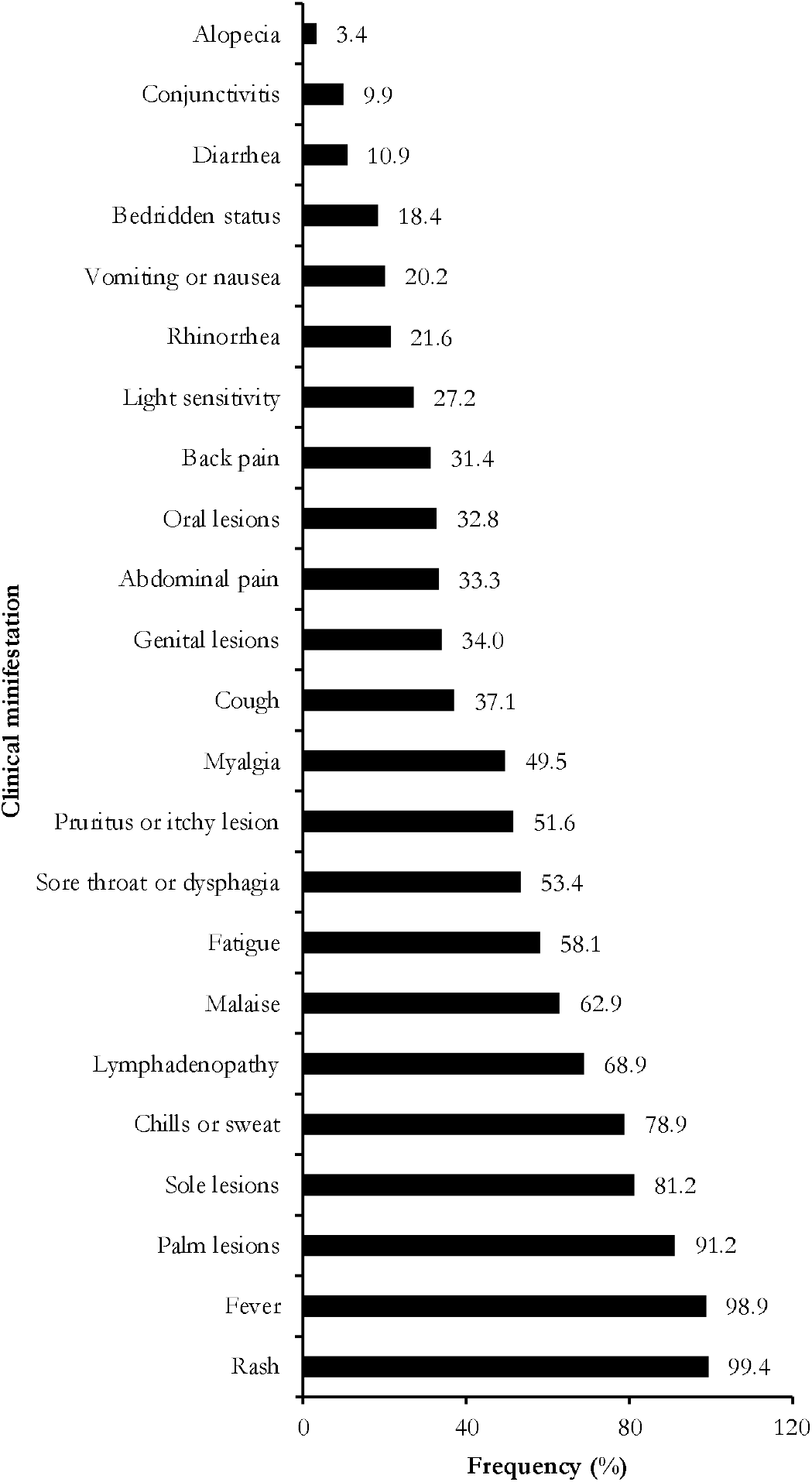
Trend of clinical manifestations observed among suspected Mpox cases in DRC,1970-2024

### Meta-regression analysis

The multivariate analysis of suspected Mpox cases revealed a significant temporal increase in VZV prevalence (β = 0.2247, *p* < 0.001), while regional studies exhibited significantly higher VZV estimates compared to nationwide ones (β=4.7148, *p* <0.001). For confirmed Mpox cases, regional studies had higher VZV estimates (β = 4.2220, *p* = 0.018). South Kivu had significantly higher HIV co-infection rates among confirmed Mpox cases compared to other regions (β = 3.608, *p* = 0.002). Participant type (*p* = 0.577 and p=0.266), disease burden for suspected cases (*p* = 0.164), study year for confirmed cases (*p* = 0.959), study design (*p* = 0.826), and setting (*p* = 0.159) did not significantly explain heterogeneity at both uni-and multivariate analysis (Table 5).

### Publication bias

The funnel plot asymmetry suggested a risk of publication bias for our study outcomes. The trim-and-fill analysis suggested four potentially missing studies and not significantly reducing the pooled VZV prevalence among suspected Mpox cases from 16.7% (95% CI: 5.36-28.10) to 2.73% (95% CI: 0.00–18.47). After adjusting for three potentially missing studies, the pooled varicella-zoster virus and Mpox coinfection rate was 4.00% (95% CI: 0.00-13.85), not significantly different from the initial pooled estimate of 9.69% (95% CI: 1.33-18.06). Regarding HIV and Mpox coinfection, two potentially missing studies were suggested by the method, there was no significant difference between the initial rate of 0.52% (95% CI: 0.18-0.87) to 0.70% (95% CI: 0.00-1.60). (Additional files 2,3 and 4, Supplementary Fig. 6 and 7).

### Sensitivity test analysis

Sensitivity analysis of the pooled prevalence estimate of VZV among suspected Mpox cases demonstrated stable pooled estimates (95% CI: 1.88–2.17%) upon the exclusion of individual studies, supporting the robustness of the primary findings. However, omitting the Bangwen *et al*. study [17] resulted in an outlier effect (8.60%; 95% CI: 7.78–9.43), indicating that this study disproportionately influenced the meta-analysis, possibly due to unique sample characteristics or methodological differences. Sensitivity analysis of the pooled VZV and Mpox coinfection rate also revealed stable estimates (95% CI: 0.47–3.88 events/100 observations) when most studies were excluded. Conversely, excluding the Kinganda-Lusamaki *et al*. study [16] inflated the estimate to 11.66 events/100 observations, highlighting its outlier influence. The pooled estimates of the HIV and Mpox coinfection rate were not significantly influenced by the omission of any individual study included in the meta-analysis (Additional Files 2, 3, and 4, Supplementary Fig. 9, 9, and 6, respectively).

## Discussion

This systematic review and meta-analysis were conducted to determine the coinfection rate of Mpox and VZV, and the clinical profile of Mpox cases in the world’s most endemic focus. It synthesized data from 20 studies to provide a comprehensive overview in the DRC from 1970 to 2024.

### Varicella zona virus-Mpox coinfection

Our meta-analysis, examining 1,841 confirmed Mpox cases in the DRC, revealed a pooled VZV coinfection rate of 9.69% (95% CI: 1.33–18.06). This finding was marked by significant heterogeneity (*I*^*2*^ = 96.9%, *p* < 0.001), indicating substantial variability across the included studies. Subgroup analyses offered further insights into these variations. Coinfection rates showed temporal stability, with similar rates observed in 1991–2010 (12.50%) and 2011–2024 (9.47%), despite expanded surveillance efforts in the latter period. However, regional disparities were evident. The Kivu region exhibited a higher coinfection rate of 33.33% (though based on a single study), compared to the 11.10% rate in the Equateur region (across six studies).

Several factors likely contribute to the observed significant VZV-Mpox coinfection rate. The clinical similarities between VZV and Mpox rashes may lead to diagnostic overlap and misclassification, particularly in settings where PCR confirmation is limited [40]. The temporal stability of coinfection rates, despite improved diagnostics after 2010, suggests the influence of persistent underlying ecological drivers, such as deforestation and human encroachment [3].

Our findings were lower than those obtained from studies conducted in Nigeria, which reported that 27-81% of confirmed Mpox cases were coinfected with VZV [28,41]. Meanwhile, a study conducted in Belgium reported no cases of VZV among confirmed Mpox cases, suggesting that VZV did not cocirculate in the population at risk for Mpox during the Belgian 2022 outbreak, and also that Mpox does not commonly trigger reactivation of latent VZV in adult men [42].

Our results might indicate that VZV coinfection was responsible for more severity and complications among confirmed Mpox cases, as described in a study conducted in Nigeria (coinfected patients had more complications than Mpox-only infected cases (56.3% vs. 22.5%, *p* = 0.015) [41]. In addition, in some epidemiological contexts, rodents infected with the poxvirus could transmit the disease through VZV skin lesions and cause coinfection. Based on that hypothesis, the authors concluded that varicella infection is a risk factor for the acquisition of Mpox [43].

These findings have important implications for clinical practice and public health strategies in the DRC. They highlight the need for dual diagnostic approaches, potentially incorporating PCR and serological assays to accurately differentiate between VZV and Mpox coinfections and single infections [12]. Moreover, coinfected patients may require closer attention during hospital stays, as they may exhibit greater severity and more complications [41]. Furthermore, our regional findings underscore the importance of targeted interventions, such as prioritized vaccination efforts for both VZV and Mpox, in high-burden zones like Kivu [43].

### HIV-Mpox coinfection

The HIV coinfection rate among 1652 confirmed Mpox cases was 0.52% (95% CI: 0.18–0.87) with low heterogeneity (*I*^*2*^ = 39.2%, *p* = 0.160) between the five included studies. In contrast, a study reported a much higher proportion of HIV coinfection among confirmed Mpox cases (35%) within a gender-specific group from developed countries [21]. In this regard, a meta-analysis found that individuals with both HIV and Mpox had a higher hospitalization rate than those with Mpox only (OR = 1.85; 95% CI 0.918–3.719; *p* = 0.085). This provides substantial evidence that HIV and Mpox coinfection has a negative impact on clinical outcomes, including patient death [18]. Similar conclusions were drawn in a report from Nigeria [19].

### Mpox clinical pattern

The clinical presentation was dominated by rash (99.97%), painful lesions (78.17%), and malaise (77.14%), suggesting the integration of these symptoms in Mpox case definitions. However, the high heterogeneity (*I*^*2*^ > 90%) across studies underscores significant variability in clinical reporting, raising questions about the reliability of symptom-based diagnosis in resource-limited settings [1].

The near-universal prevalence of rash (99.97%) aligns with WHO and CDC case definitions, which prioritize rash as a cardinal symptom for suspected Mpox [1,9]. Despite being reported in only one study, the high frequency of painful lesions (78.17%) highlights a key diagnostic marker for differentiating Mpox in endemic regions. This finding supports the inclusion of painful rash in clinical algorithms in DRC guidelines [1].

The prevalence of systemic symptoms (fever: 67.94%; lymphadenopathy: 71.99%) further complicates differential diagnosis, as these overlap with malaria, VZV, and other febrile illnesses endemic to the DRC [44,45]. Notably, the lower fever rate compared to rash suggests that afebrile rash presentations should not exclude Mpox, challenging historical definitions requiring fever as a mandatory criterion [46].

This study highlights the need to expand clinical criteria to capture atypical presentations [47]. The extreme heterogeneity (*I*^*2*^ > 90%) in symptom reporting underscores the importance of standardizing clinical assessments across healthcare tiers and implementing dual-pathogen testing (Mpox/VZV PCR) to improve diagnostic accuracy, especially given the overlapping symptoms with other endemic diseases [10]. These results advocate for context-adapted surveillance protocols that account for regional variability in HIV prevalence and healthcare access, with particular attention to conflict-affected zones like Kivu where diagnostic challenges are amplified, ultimately calling for investments in healthcare worker training and decentralized laboratory capacity to strengthen early detection and outbreak response in these high-burden settings [48,49].

## Strength and Limitations

This review has several limitations. The small number of included studies (□10) constrained us to rely only on the funnel plot to assess publication bias, as the traditional Egger and Begg’s tests could not be performed. Additionally, the small sample sizes in some subgroup analyses resulted in wide confidence intervals, limiting the precision of the findings. Despite these limitations, this systematic review and meta-analysis provides valuable insights into Mpox coinfection with other viral diseases, including VZV and HIV, in the DRC. The findings highlight the need for continued public health surveillance, research, and interventions to reduce and prevent disease complications among infected patients.

## Conclusions

This systematic review and meta-analysis highlight the distinct clinical profile of Mpox in the DRC, characterized by near-universal rash presentation and frequent systemic symptoms, revealing gaps in current case definitions and surveillance systems. Despite the low HIV coinfection rate observed, such patients should benefit from particular attention. However, the high heterogeneity in symptom reporting and significant VZV coinfection rates underscore the urgent need for standardized, context-adapted diagnostic protocols that integrate dual-pathogen testing and expand criteria to capture atypical presentations. These findings call for targeted investments in healthcare worker training, laboratory capacity, and regionally tailored surveillance strategies to improve early detection and outbreak response in Africa’s highest-burden settings, where Mpox remains an ongoing public health threat.

## Data Availability

All data generated or analyzed during this study are included in this published article and supplemental material.

## Abbreviations

*CI*: Confidence interval
*DRC*: Democratic Republic of Congo
*HCW*: Healthcare worker
*HIV/AIDS*: Human immunodeficiency virus/acquired immunodeficiency syndrome
*MeSH:*: Medical subject headings
Mpox: Monkeypox
*PCR*: Polymerase Chain Reaction
*PRISMA*: Preferred reporting items for systematic reviews and meta-analysis
*VZV:*: Varicella-zona virus

## Declarations

**Author contributions:** F.Z.L.C. conceived the original idea of the study. F.Z.L.C and W.N.B. conducted the literature search. F.Z.L.C., C.A. and C.T.A. selected the studies, extracted the relevant information, and synthesized the data. F.Z.L.C. performed the analyses and wrote the first draft of the manuscript. All authors critically reviewed and revised successive drafts of the manuscript. All authors read and approved the final manuscript.

**Ethical Approval Statement** : Not applicable

**Consent for publication**: Not applicable.

**Availability of data and materials**: The sources of data supporting this systematic review are available in the reference. All data generated or analyzed during this study are included in this published article and supplemental material.

**Competing interests**: All authors declare no conflicts of interest and have approved the final version of the article.

**Funding source**: This research did not receive any specific grant from funding agencies in the public, commercial or not-for-profit sectors.

## References

1. Mpox. WHO, Geneva. 2024. https://www.who.int/news-room/fact-sheets/detail/mpox. Accessed 2025 Apr 27.

2. Djuicy DD, Sadeuh-Mba SA, Bilounga CN, Yonga MG, Tchatchueng-Mbougua JB, Essima GD, et al. Concurrent Clade I and Clade II Monkeypox Virus Circulation, Cameroon, 1979–2022. Emerg Infect Dis. 2024;30(3):433–43.

3. Cheuyem FZL, Zemsi A, Ndungo JH, Achangwa C, Takpando-le-grand DR, Goupeyou-Youmsi JM, et al. Mpox Severity and Mortality in the Most Endemic Focus in Africa: A Systematic Review and Meta-Analysis (1970-2024). medRxiv; 2025. doi: 10.1101/2025.04.07.25325410v1.

4. Osadebe L, Hughes CM, Shongo Lushima R, Kabamba J, Nguete B, Malekani J, et al. Enhancing case definitions for surveillance of human monkeypox in the Democratic Republic of Congo. Kasper M, editor. PLoS Negl Trop Dis. 2017;11(9):e0005857.

5. Brosius I, Vakaniaki EH, Mukari G, Munganga P, Tshomba JC, Vos ED, et al. Epidemiological and clinical features of mpox during the clade Ib outbreak in South Kivu, Democratic Republic of the Congo: a prospective cohort study. The Lancet. 2025;405(10478):547–59.

6. Masirika Lm, Udahemuka Jc, Ndishimye P, Martinez Gs, Kelvin P, Nadine Mb, et al. Epidemiology, clinical characteristics, and transmission patterns of a novel Mpox (Monkeypox) outbreak in eastern Democratic Republic of the Congo (DRC): an observational, cross-sectional cohort study. medRxiv; 2024. doi: 10.1101/2024.03.05.24303395v1.

7. Dieu-Merci. KY, Singa Valentin. B, Ley. Bl, Antoine. Et, Guild. AK, Lwanga. K, et al. Mpox: epidemiological profile and factors associated with its emergence in the Isangi territory, Democratic Republic of Congo. Int J Appl Sci Eng Rev. 2024;5(5):27–43.

8. Coronavirus (COVID-19) Dashboard. WHO, Geneva. 2022. https://covid19.who.int. Accessed 2022 Oct 20.

9. CDC. Signs and Symptoms of Mpox. CDC, Atlanta. 2024. https://www.cdc.gov/mpox/signs-symptoms/index.html. Accessed 2025 Apr 27.

10. Whitehouse ER, Bonwitt J, Hughes CM, Lushima RS, Likafi T, Nguete B, et al. Clinical and Epidemiological Findings from Enhanced Monkeypox Surveillance in Tshuapa Province, Democratic Republic of the Congo During 2011–2015. J Infect Dis. 2021;223(11):1870–8.

11. Hughes CM, Liu L, Davidson WB, Radford KW, Wilkins K, Monroe B, et al. A Tale of Two Viruses: Coinfections of Monkeypox and Varicella Zoster Virus in the Democratic Republic of Congo. Am J Trop Med Hyg. 2021;104(2):604–11.

12. Pittman PR, Martin JW, Kingebeni PM, Tamfum JJM, Mwema G, Wan Q, et al. Clinical characterization and placental pathology of mpox infection in hospitalized patients in the Democratic Republic of the Congo. Bowman N, editor. PLoS Negl Trop Dis. 2023;17(4):e0010384.

13. Kibungu EM, Vakaniaki EH, Kinganda-Lusamaki E, Kalonji-Mukendi T, Pukuta E, Hoff NA, et al. Clade I–Associated Mpox Cases Associated with Sexual Contact, the Democratic Republic of the Congo. Emerg Infect Dis. 2024;30(1):172–6.

14. Mukadi-Bamuleka D, Kinganda-Lusamaki E, Mulopo-Mukanya N, Amuri-Aziza A, O’Toole Á, Modadra-Madakpa B, et al. First imported Cases of MPXV Clade Ib in Goma, Democratic Republic of the Congo: Implications for Global Surveillance and Transmission Dynamics. medRxiv; 2024. doi: 10.1101/2024.09.12.24313188.

15. Mpox - Democratic Republic of the Congo. WHO, Geneva. 2024. https://www.who.int/emergencies/disease-outbreak-news/item/2024-DON522. Accessed 2025 Apr 27.

16. Kinganda-Lusamaki E, Baketana LK, Ndomba-Mukanya E, Bouillin J, Thaurignac G, Aziza AA, et al. Use of Mpox Multiplex Serology in the Identification of Cases and Outbreak Investigations in the Democratic Republic of the Congo (DRC). Pathogens. 2023;12(7):916.

17. Bangwen E, Diavita R, Vos ED, Vakaniaki EH, Nundu SS, Mutombo A, et al. Suspected and confirmed mpox cases in DR Congo: a retrospective analysis of national epidemiological and laboratory surveillance data, 2010–23. The Lancet. 2025;405(10476):408–19.

18. Taha AM, Elrosasy A, Mahmoud AM, Saed SAA, Moawad WAE, Hamouda E, et al. The effect of HIV and mpox co-infection on clinical outcomes: Systematic review and meta-analysis. HIV Med. 2024;25(8):897–909.

19. Mmerem JI, Johnson SM, Iroezindu MO. Monkeypox and chickenpox co-infection in a person living with Human Immunodeficiency Virus. J Infect Dev Ctries. 2024;18(07):1152–6.

20. Tati G. Territory and border crossing for livelihoods among (voluntary and forced) migrants from DRC to Swaziland: the re-imagining of a borderless spatial system. In: Udelsmann Rodrigues C, Tomàs J, editors. Crossing African Borders: Migration and Mobility. Lisboa: Centro de Estudos Internacionais; 2012. p. 126–44. (ebook’IS).

21. Liu BM, Rakhmanina NY, Yang Z, Bukrinsky MI. Mpox (Monkeypox) Virus and Its Co-Infection with HIV, Sexually Transmitted Infections, or Bacterial Superinfections: Double Whammy or a New Prime Culprit? Viruses. 2024;16(5):784.

22. Doshi RH, Guagliardo SAJ, Dzabatou-Babeaux A, Likouayoulou C, Ndakala N, Moses C, et al. Strengthening of Surveillance during Monkeypox Outbreak, Republic of the Congo, 2017. Emerg Infect Dis. 2018 ;24(6) :1158–60.

23. Musuka G, Moyo E, Tungwarara N, Mhango M, Pierre G, Saramba E, et al. A critical review of mpox outbreaks, risk factors, and prevention efforts in Africa: lessons learned and evolving practices. IJID Reg. 2024;12(2024):100402.

24. Martins-Filho PR, Dorea FCM, Sena LOC, Bezerra GVB, Teixeira DCP, Damaso CR, et al. First reports of monkeypox and varicella-zoster virus coinfection in the global human monkeypox outbreak in 2022. Travel Med Infect Dis. 2023; 51:102510.

25. Du M, Niu B, Liu J. The research and development landscape for mpox vaccines. Lancet Infect Dis. 2025;25(4):e198–9.

26. Shafaati M, Forghani S, Shahsavand Davoudi A, Samiee R, Mohammadi K, Akbarpour S, et al. Current advances and challenges in mpox vaccine development: a global landscape. Ther Adv Vaccines Immunother. 2025;13:25151355251314339.

27. Petrichko S, Kindrachuk J, Nkamba D, Halbrook M, Merritt S, Kalengi H, et al. Mpox Vaccine Acceptance, Democratic Republic of the Congo. Emerg Infect Dis. 2024;30(12):2614–9.

28. Stephen R, Alele F, Olumoh J, Tyndall J, Okeke MI, Adegboye O. The epidemiological trend of monkeypox and monkeypox-varicella zoster viruses co-infection in North-Eastern Nigeria. Front Public Health. 2022. doi: 10.3389/fpubh.2022.1066589.

29. Moher D, Liberati A, Tetzlaff J, Altman DG, Group TP. Preferred Reporting Items for Systematic Reviews and Meta-Analyses: The PRISMA Statement. PLOS Med. 2009;6(7):e1000097.

30. Congo, Democratic Republic of the. In: The World Factbook. Central Intelligence Agency; 2023. https://www.cia.gov/the-world-factbook/countries/congo-democratic-republic-of-the/. xAccessed 2023 Oct 30.

31. République démocratique du Congo - Plan national de développement sanitaire recadré pour la période 2019-2022□: Vers la Couverture sanitaire universelle. https://www.ilo.org/dyn/natlex/natlex4.detail?p_isn=111796&p_lang=fr. Accessed 2023 Oct 30.

32. Democratic Republic of the Congo. WHO, Geneva. 2024. https://data.who.int/countries/180. Accessed 2025 Mar 27.

33. Amani A, Bene ACM, Lungoyo CL, Mukoka AK, Cheuyem FZL, Mpinganjira S, et al. The Struggle to Vaccinate: Unveiling the Reality of the first year of Covid-19 Vaccination in the Democratic Republic of Congo. medRxiv; 2024. doi: 10.1101/2024.01.03.24300795v1

34. Overview. World Bank. 2023. https://www.worldbank.org/en/country/drc/overview. Accessed 2023 Oct 30.

35. Democratic Republic of the Congo situation. UNHCR, Global Focus, Geneva. 2024. https://reporting.unhcr.org/operational/situations/democratic-republic-congo-situation. Accessed 2025 Apr 6.

36. JBI Critical Appraisal Tools. The Joanna Briggs Institute, Adelaide. 2017. Available from: https://jbi.global/critical-appraisal-tools. Accessed 2024 Dec 3.

37. Hutin YJF, Williams RJ, Malfait P, Pebody R, Loparev VN, Ropp SL, et al. Outbreak of Human Monkeypox, Democratic Republic of Congo, 1996 to 1997. Emerg Infect Dis. 2001;7(3):434–8.

38. R Core Team. R: A Language and Environment for Statistical Computing. R Foundation for Statistical Computing, Vienna, Austria. 2024. Available from: https://www.R-project.org/. Accessed 2024 Dec 19.

39. Shi L, Lin L. The trim-and-fill method for publication bias: practical guidelines and recommendations based on a large database of meta-analyses. Medicine (Baltimore). 2019;98(23):e15987.

40. Bourner J, Garcia-Gallo E, Mbrenga F, Boum Y, Nakouné E, Paterson A, et al. Challenges in clinical diagnosis of Clade I Mpox: Highlighting the need for enhanced diagnostic approaches. PLoS Negl Trop Dis. 2024;18(6):e0012087.

41. Mmerem JI, Umenzekwe CC, Johnson SM, Onukak AE, Chika-Igwenyi NM, Chukwu SK, et al. Mpox and Chickenpox Coinfection: Case Series from Southern Nigeria. J Infect Dis. 2024;229(Supplement_2): S260–4.

42. Coppens J, Liesenborghs L, Vercauteren K, Van Esbroeck M, Van Dijck C. No Varicella Zoster Virus Infection among Mpox Cases in Antwerp, Belgium. Am J Trop Med Hyg. 2023;109(6):1282–3.

43. Grose C. Surveillance of Nigerian children suggests that varicella may be a risk factor for acquisition of monkeypox. Front Public Health. 2023;11:1140956.

44. Endy TP. Viral Febrile Illnesses and Emerging Pathogens. In: Hunter’s Tropical Medicine and Emerging Infectious Diseases. Elsevier; 2020. p. 325–50.

45. Acute respiratory infections complicated by malaria (previously undiagnosed disease) - Democratic Republic of the Congo. WHO, Geneva. 2024. https://www.who.int/emergencies/disease-outbreak-news/item/2024-DON547. Accessed 2025 Apr 27.

46. Breman JG, Kalisa-Ruti null, Steniowski MV, Zanotto E, Gromyko AI, Arita I. Human monkeypox, 1970-79. Bull World Health Organ. 1980;58(2):165–82.

47. Mpox Virus Infection 2022 Case Definition. 2024. https://ndc.services.cdc.gov/case-definitions/monkeypox-virus-infection-2023/. Accessed 2025 Apr 28.

48. Basimane Bisimwa P, Koyaweda GW, Bihehe Masemo D, Ayagirwe RBB, Birindwa AB, Bisimwa PN, et al. High prevalence of hepatitis B and HIV among women survivors of sexual violence in South Kivu province, eastern Democratic Republic of Congo. PloS One. 2024;19(7):e0292473.

49. Duhant A, Kusinza B, Tantet C, Bisimwa B, Gare M, Masemo B, et al. HIV-1 infection in South Kivu (Democratic Republic of Congo): high genotypic resistance to antiretrovirals. J Antimicrob Chemother. 2023;78(7):1732–9.

