## Additional Files 2 for "Mpox clinical features and varicella-zoster virus coinfection in the Democratic Republic of Congo: a systematic review and meta-analysis (1970–2024)"

### Subgroup analysis

Year of study

**Event rate (%)**

**VZV cases proportion (%)**

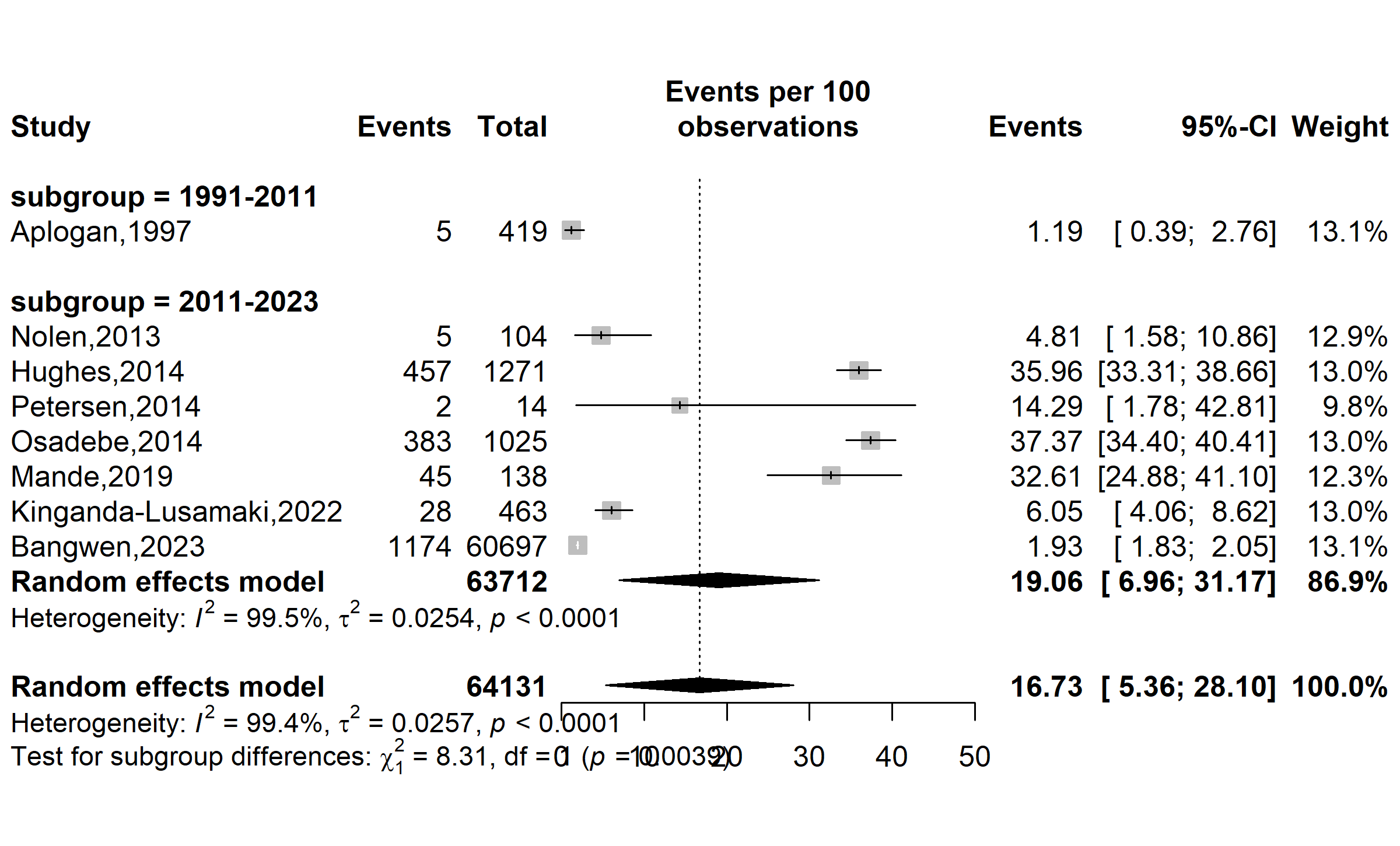

**1991-2010**

Supplementary Fig. 1 Subgroup analysis of the varicella-zoster virus prevalence rate among suspected Mpox cases in DRC, 1970-2024 *(based on policy or healthcare system changes: 1970–1990: Limited healthcare infrastructure in endemic regions;1991-2010: Improvements in healthcare access and disease surveillance; 2011-2024: Strengthened global health initiatives and response systems; VZV: Varicella-Zoster Virus)*

Study location

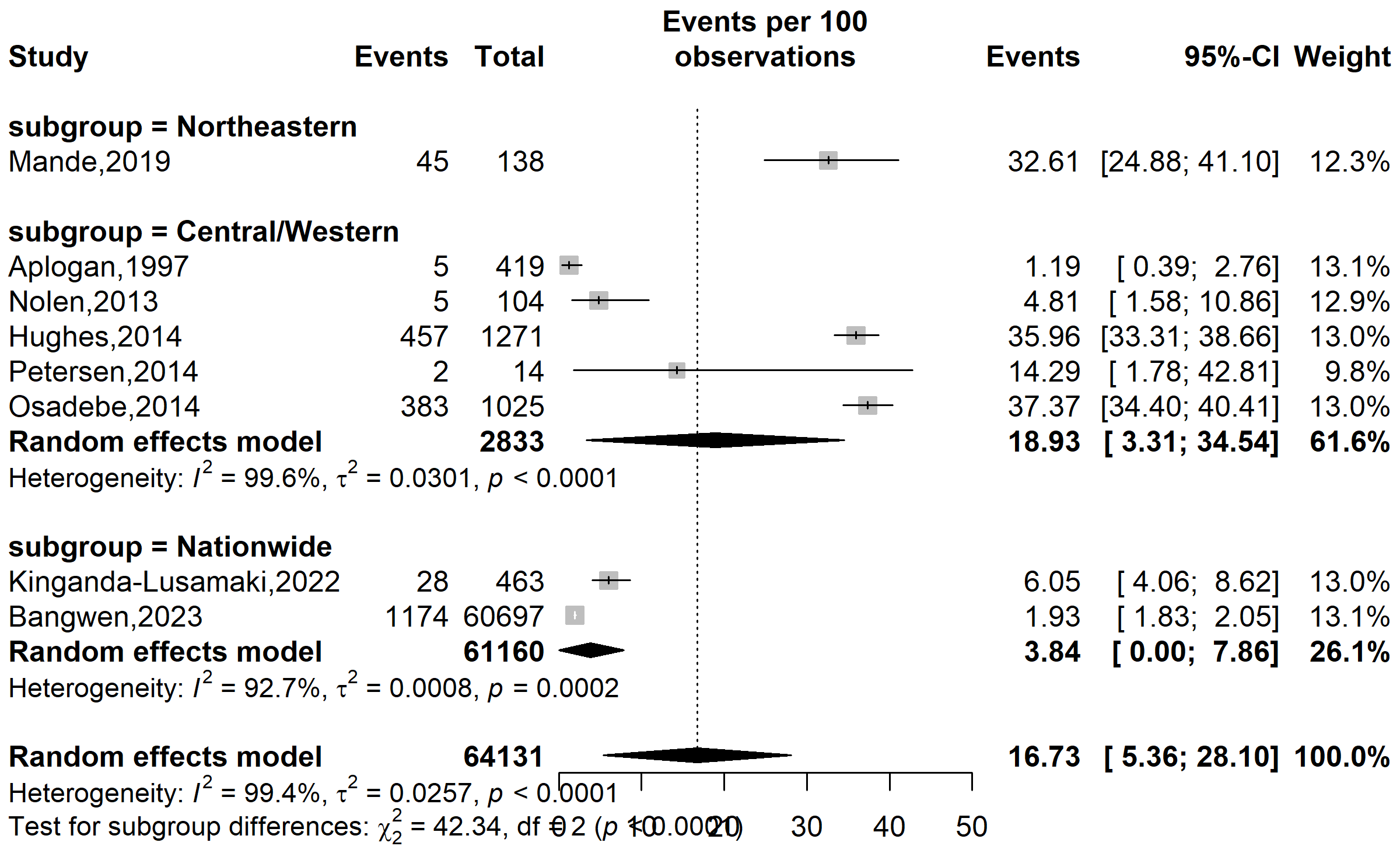

**Event rate (%)**

**VZV cases proportion (%)**

Supplementary Fig. 2 Subgroup analysis of the varicella-zoster virus prevalence rate among suspected Mpox cases in DRC, 1970-2024 *(based on geographical location Northeastern: Bas-Uélé, Central/Western: Tshuapa and Kasai oriental; VZV: Varicella-Zoster Virus)*

Study setting

**VZV cases proportion (%)**

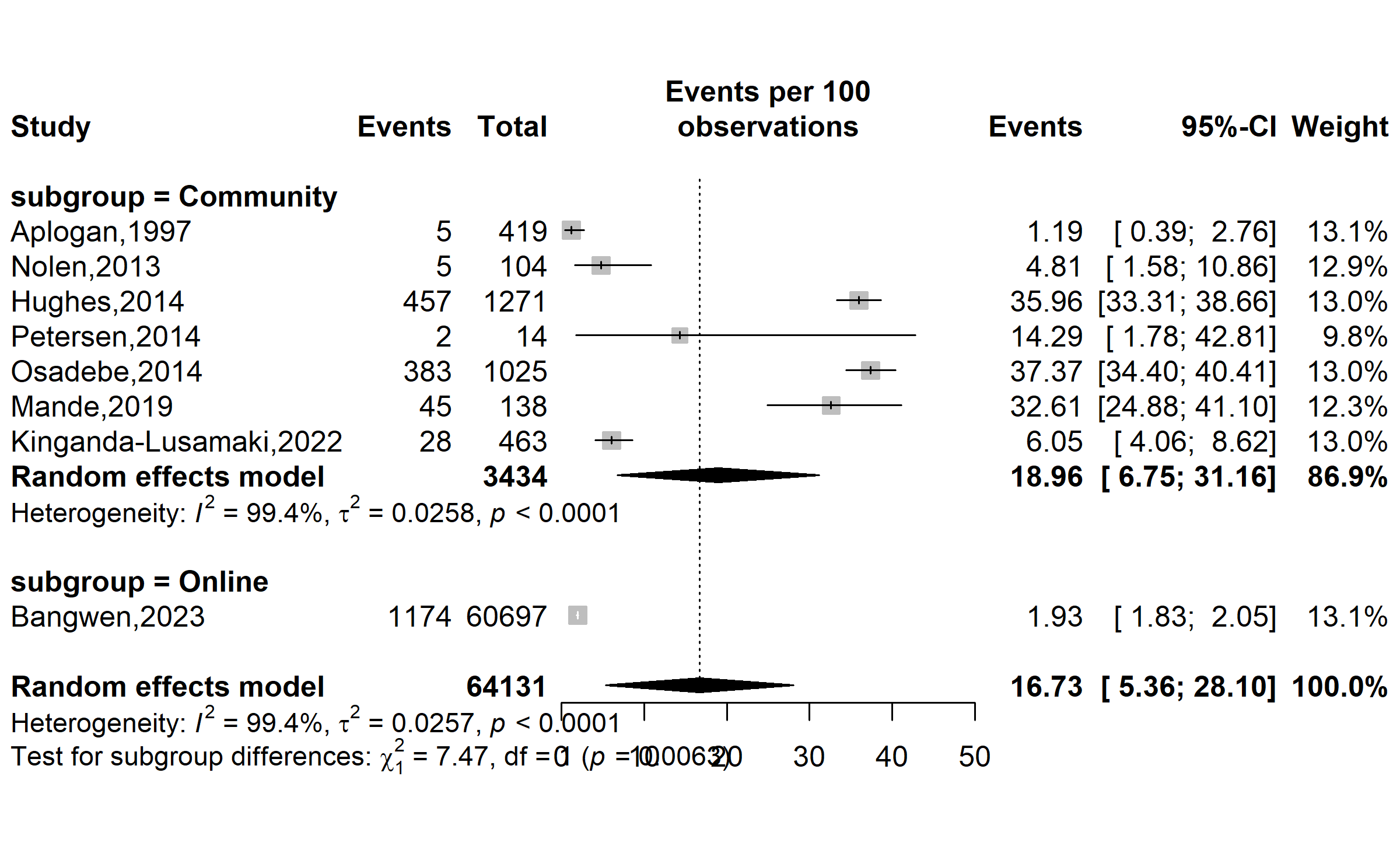

**Event rate (%)**

Supplementary Fig. 3 Subgroup analysis of the varicella-zoster virus prevalence rate among suspected Mpox cases in DRC, 1970-2024 *(based on the setting of data collection; VZV: Varicella-Zoster Virus)*

Type of participants

**Event rate (%)**

**VZV cases proportion (%)**

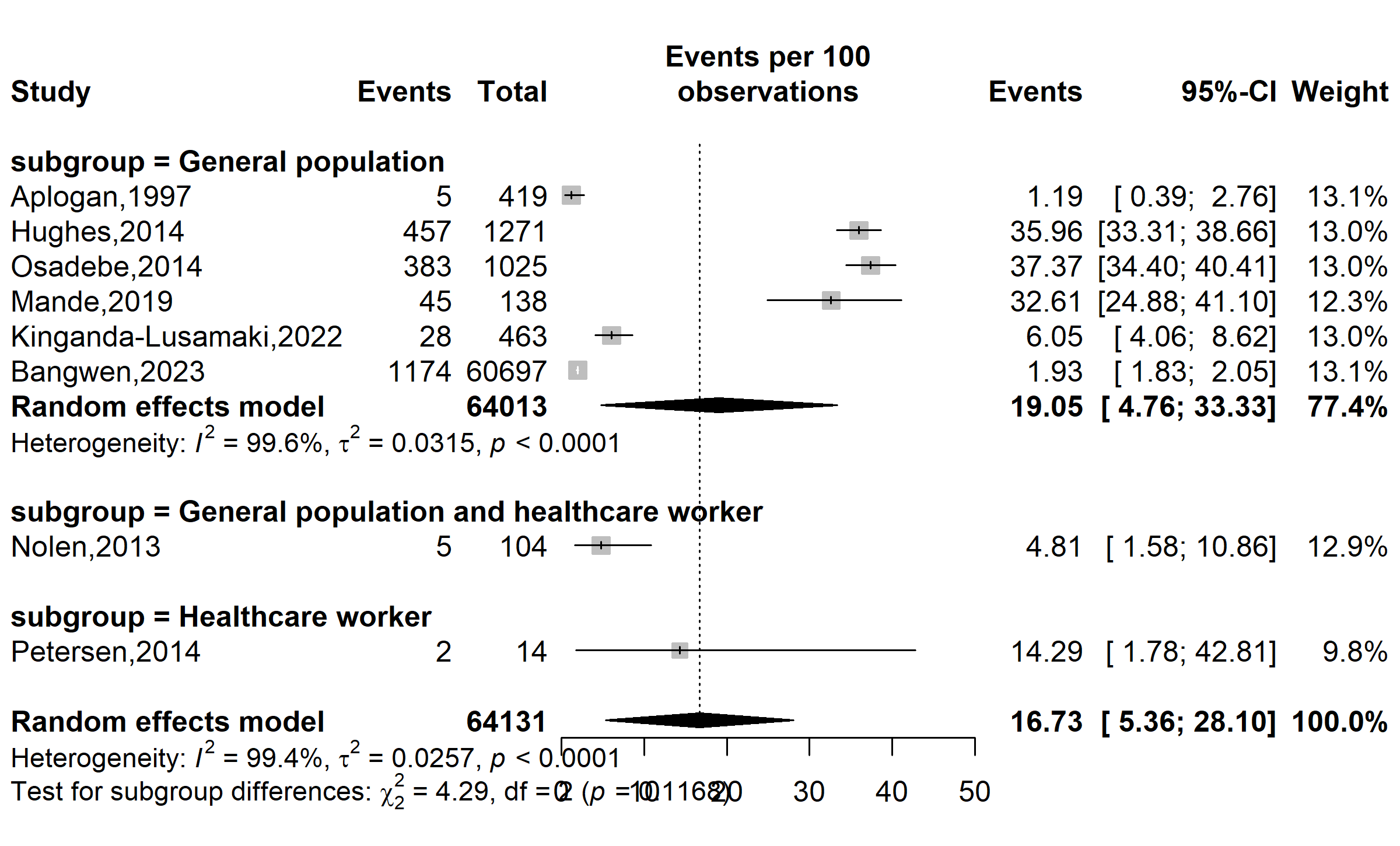

Supplementary Fig. 4 Subgroup analysis of the varicella-zoster virus prevalence rate among suspected Mpox cases in DRC, 1970-2024 *(based on the type of study participants; VZV: Varicella-Zoster Virus)*

Disease burden

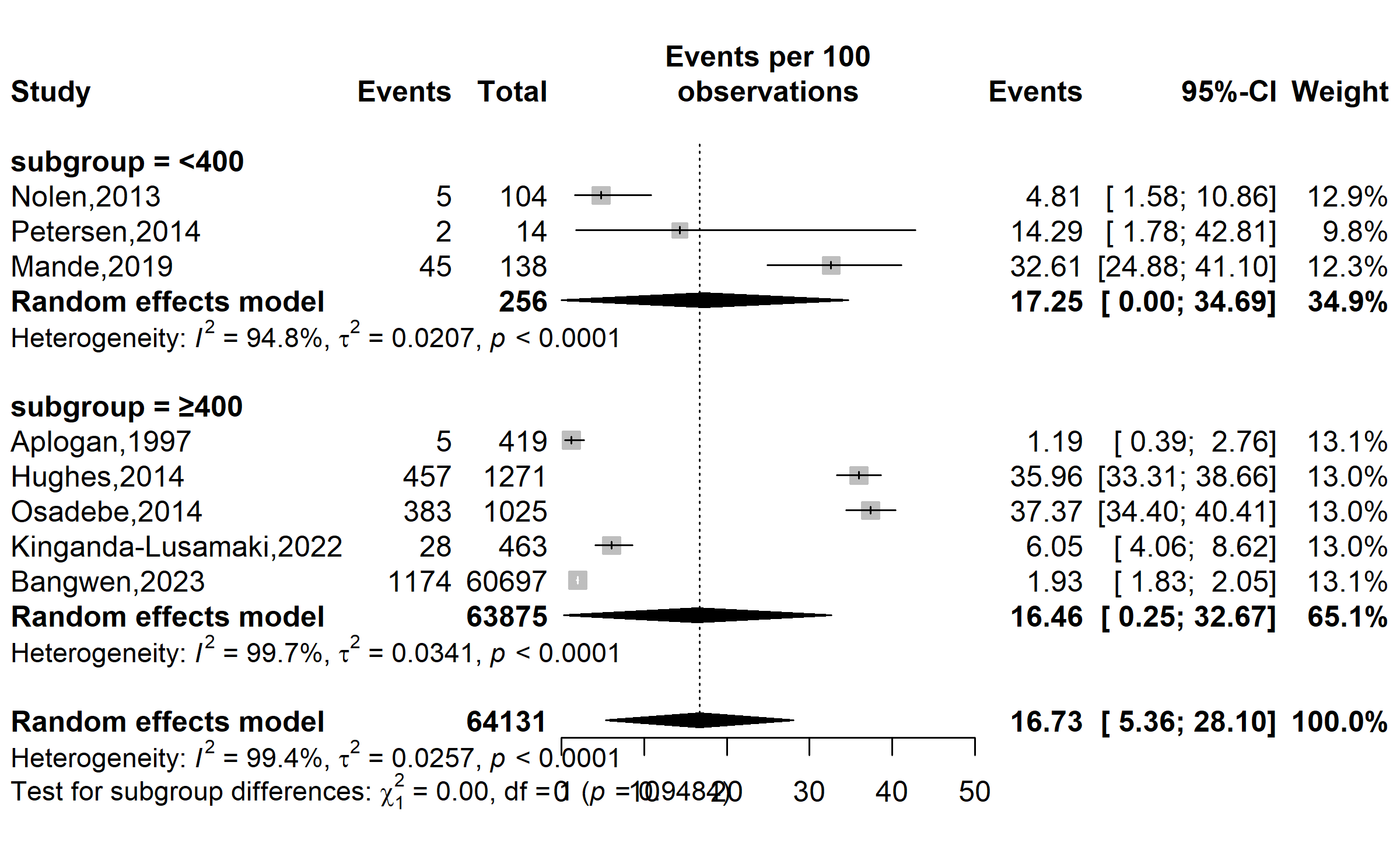

**VZV cases proportion (%)**

**Event rate (%)**

Supplementary Fig. 5 Subgroup analysis of the varicella-zoster virus prevalence rate among suspected Mpox cases in DRC, 1970-2024 *(based on the median suspected Mpox prevalence; VZV: Varicella-Zoster Virus)*

### Publication bias assessment

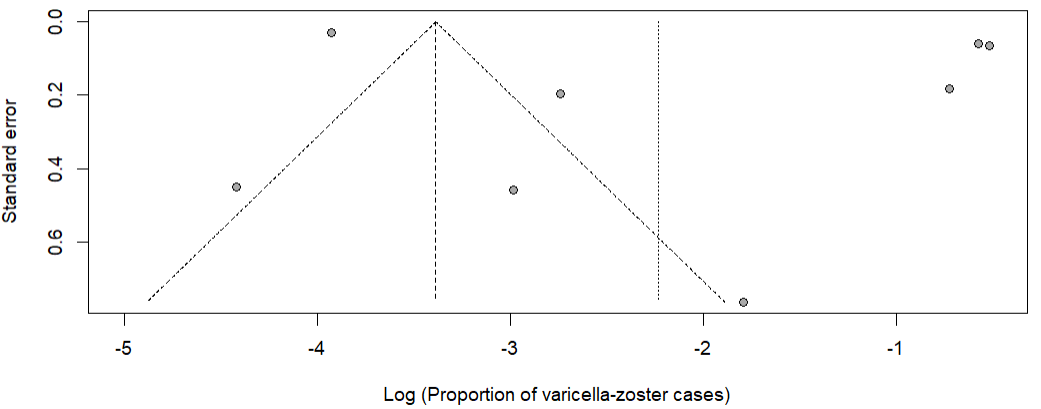

Supplementary Fig. 6 Funnel plot with pseudo 95% confidence limits and tests assessing the publication bias studies included

**VZV cases proportion (%)**

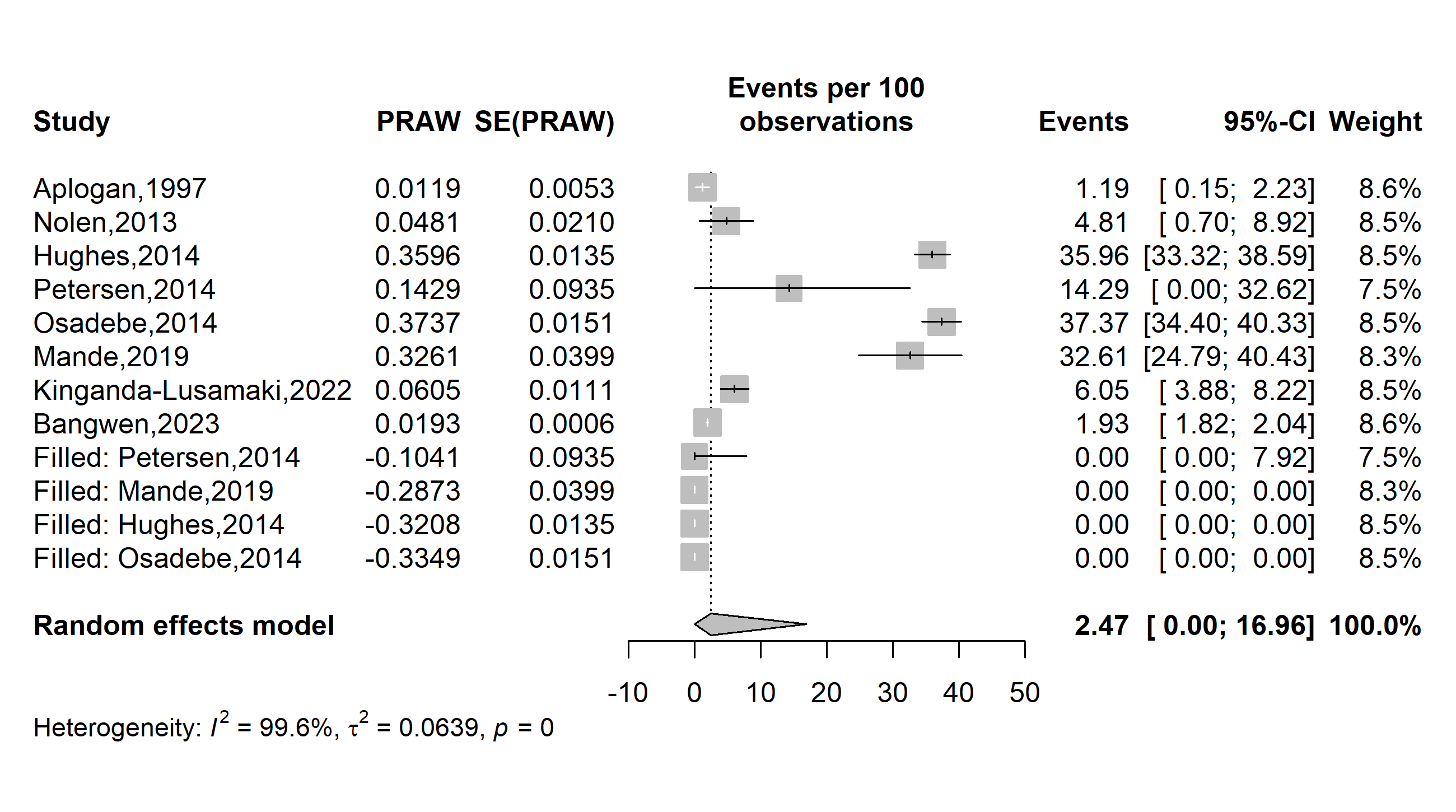

**Event rate (%)**

Supplementary Fig. 7 Funnel plot of the trim-and-fill method addressing publication bias *(VZV: Varicella-zoster virus)*

### Sensitivity analysis

**VZV proportion (%)**

**Event rate (%)**

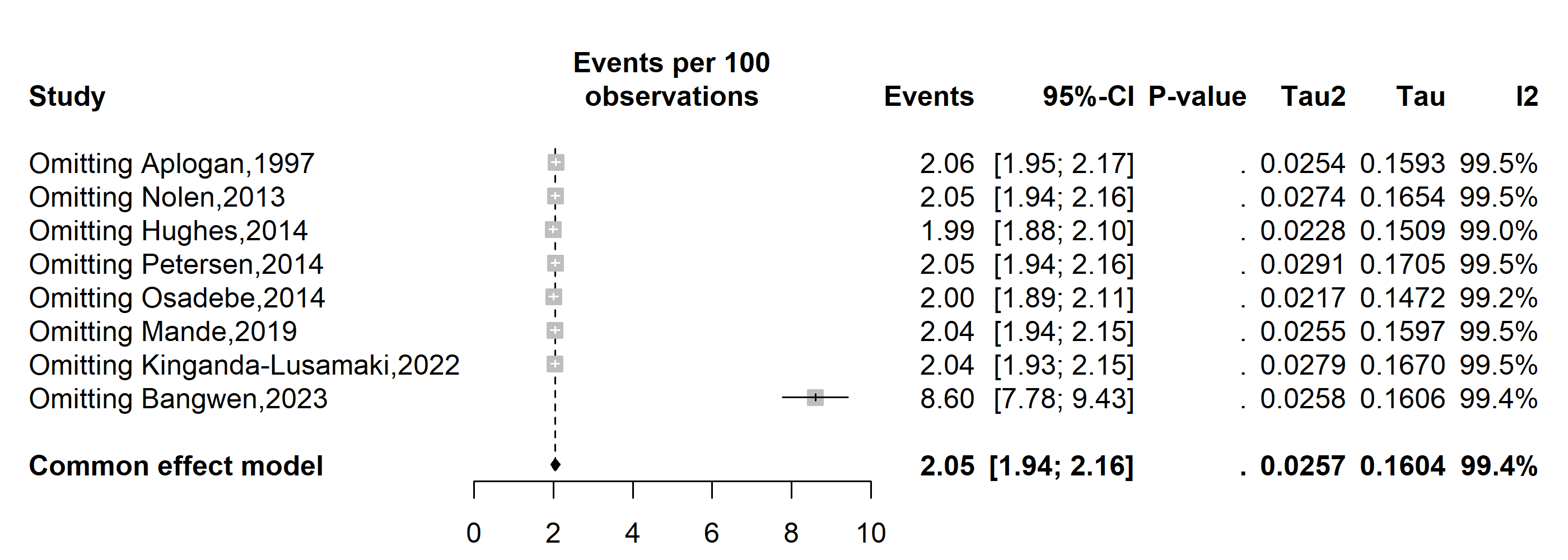

Supplementary Fig. 9 Sensitivity analysis of the pooled proportion estimate of varicella-zoster virus cases among suspected Mpox cases in DRC, 1970-2024
