## Additional Files 3 for "Mpox clinical features and varicella-zoster virus coinfection in the Democratic Republic of Congo: a systematic review and meta-analysis (1970–2024)"

Year of study

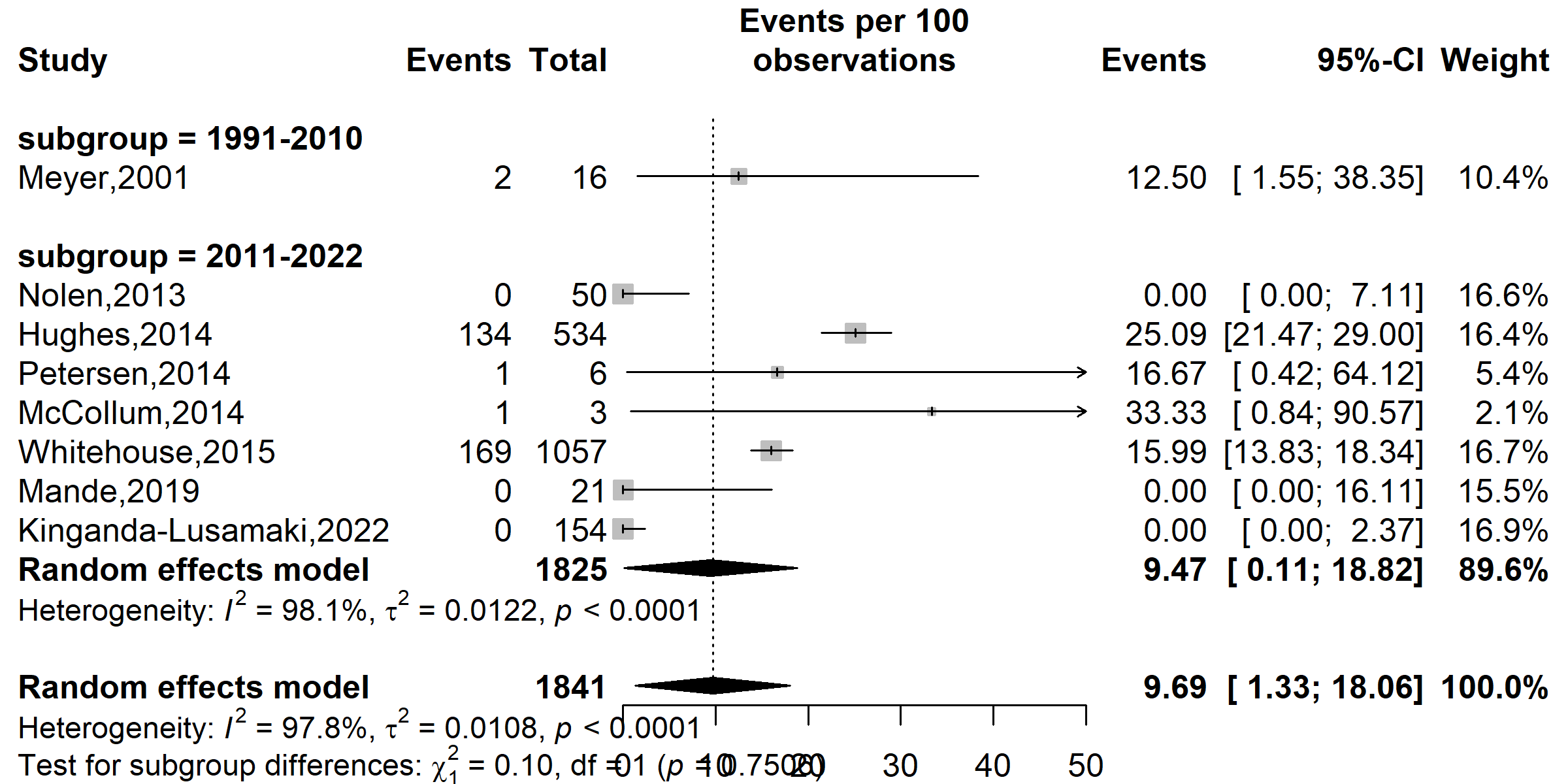

**Event rate (%)**

**Coinfection proportion (%)**

**Supplementary Fig. 1** Subgroup analysis of the proportion of varicella-zoster virus and Mpox coinfection in DRC, 1970-2024 *(based on policy or healthcare system changes: 1970–1990: Limited healthcare infrastructure in endemic regions; 1991-2010: Improvements in healthcare access and disease surveillance; 2011-2024: Strengthened global health initiatives and response systems)*

Study location

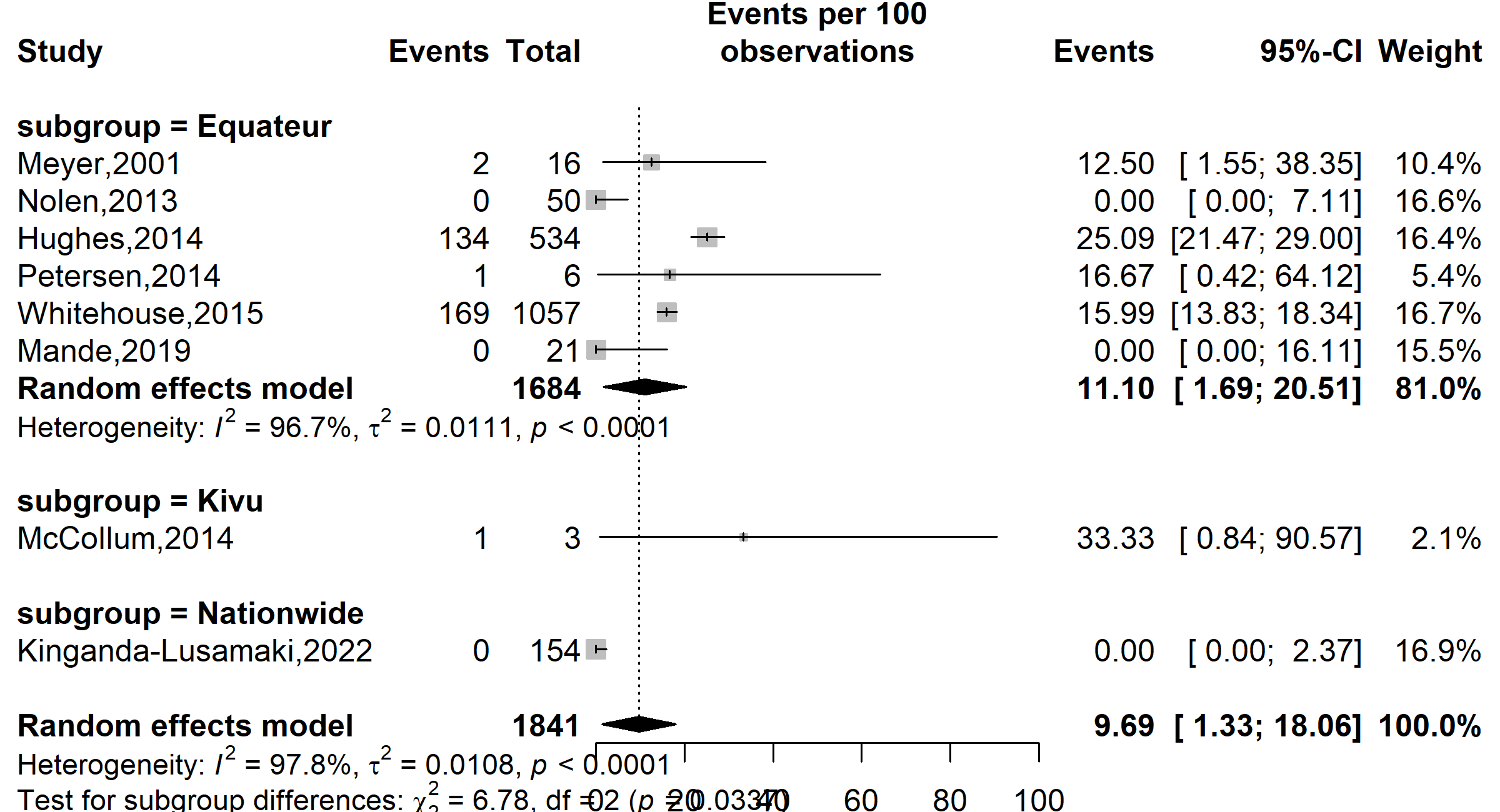

**Coinfection proportion (%)**

**Event rate (%)**

Supplementary Fig. 2 Subgroup analysis of the proportion of varicella-zoster virus and Mpox coinfection in DRC, 1970-2024 *(based on geographical location: Equateur= Equateur, Tshuapa and Bas-Uélé, Kivu (North and South))*

Type of participants

**Coinfection proportion (%)**

**Event rate (%)**

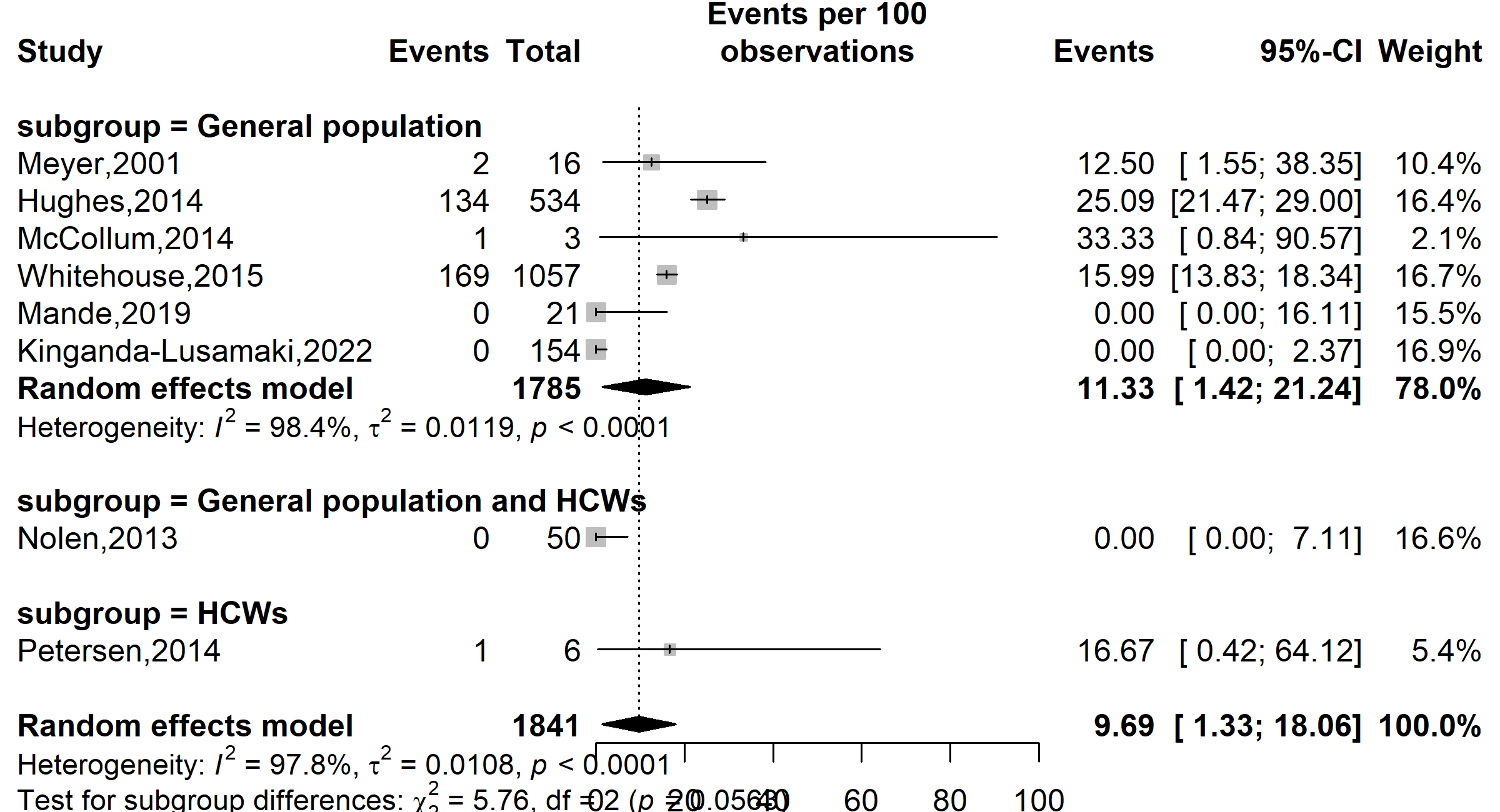

Supplementary Fig. 4 Subgroup analysis of the proportion of varicella-zoster virus and Mpox coinfection in DRC, 1970-2024 *(based on the type of study participants, HCW: Healthcare Worker)*

Disease burden

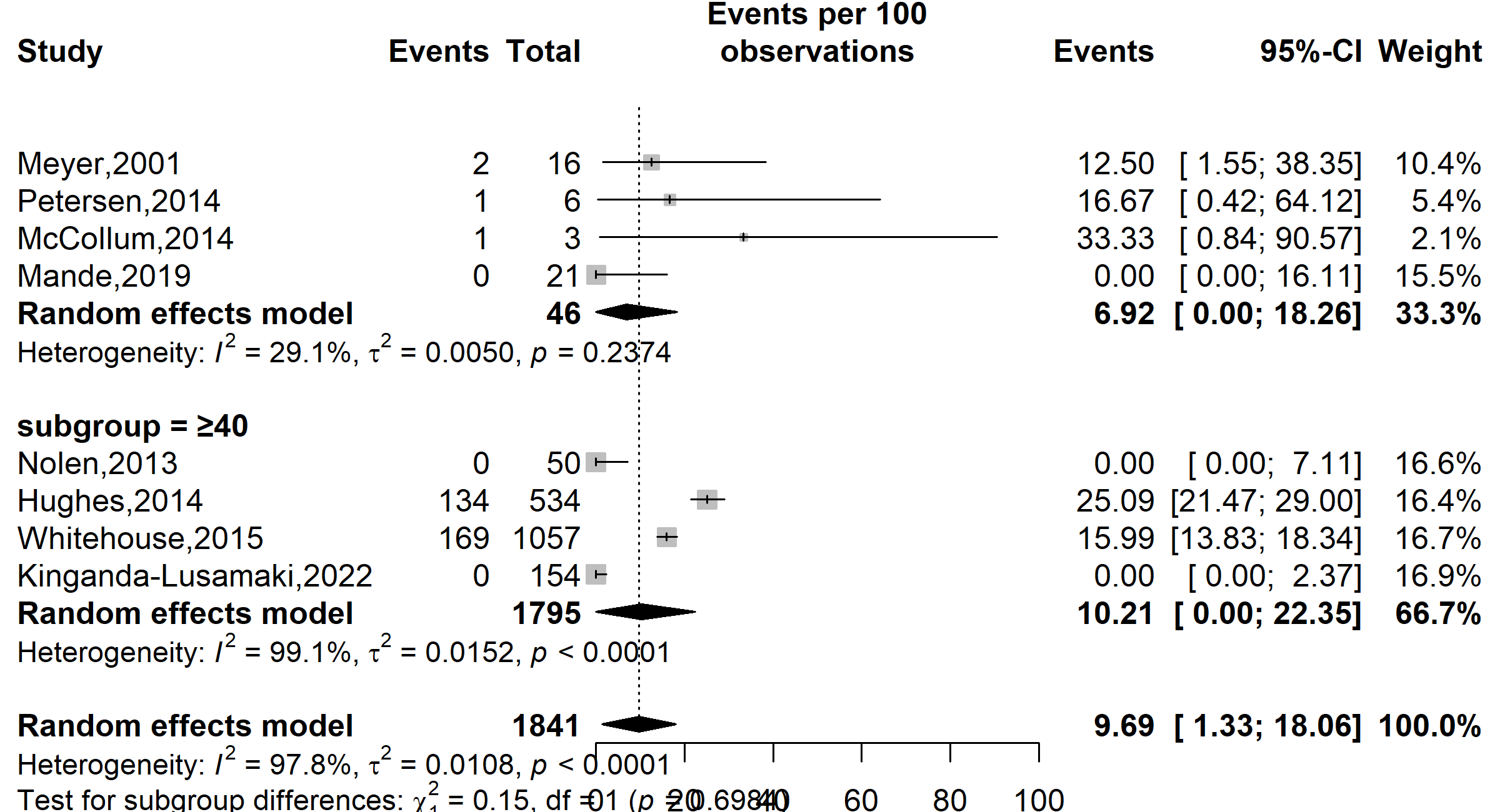

**Event rate (%)**

**Coinfection proportion (%)**

**Subgroup = ˂40**

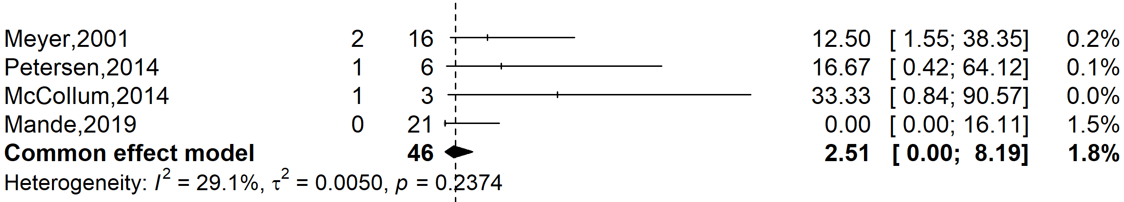

Supplementary Fig. 5 Subgroup analysis of the proportion of varicella-zoster virus and Mpox coinfection in DRC, 1970-2024 *(based median confirmed Mpox prevalence)*

### Publication bias assessment

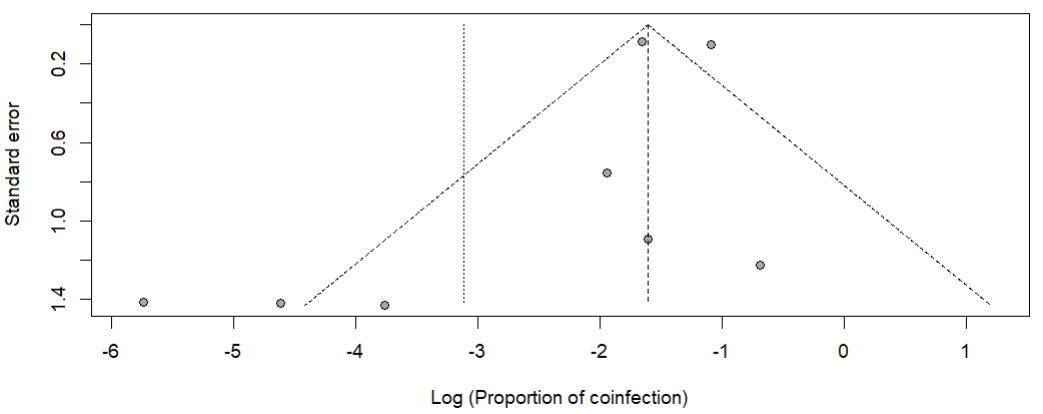

Supplementary Fig. 6 Funnel plot with pseudo 95% confidence limits and tests assessing the publication bias studies included

**Event rate (%)**

**Coinfection proportion (%)**

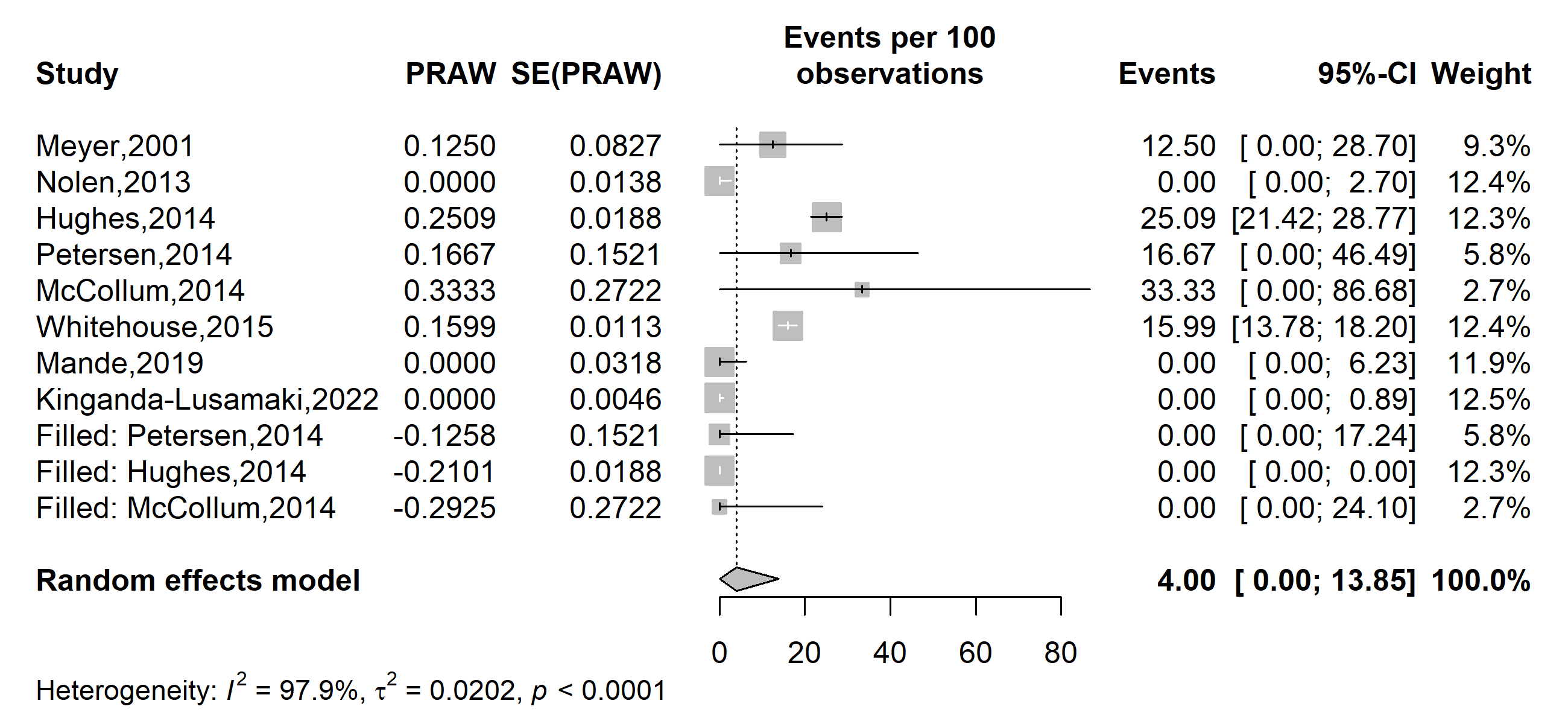

Supplementary Fig. 7 Funnel plot of the trim-and-fill method addressing publication bias

### Sensitivity analysis

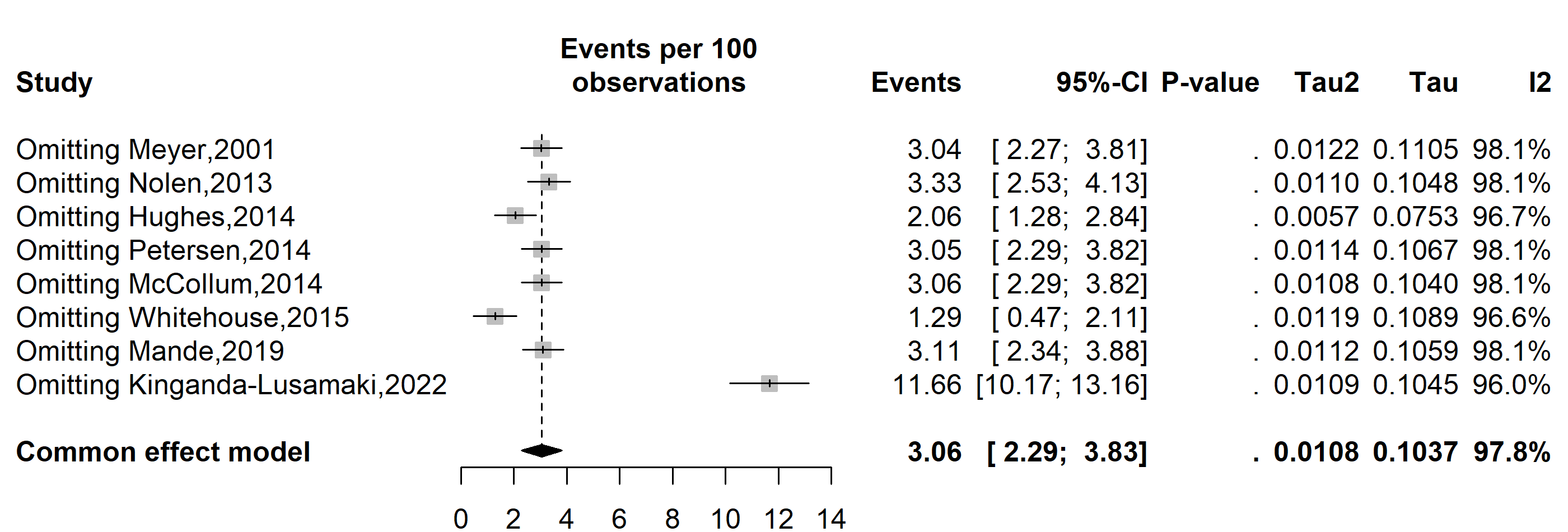

**Coinfection proportion (%)**

**Event rate (%)**

Supplementary Fig. 9 Sensitivity analysis of the pooled proportion estimate of varicella-zoster virus coinfection among confirmed Mpox cases in DRC, 1970-2024
