## Additional Files 4 for "Mpox clinical features and varicella-zoster virus coinfection in the Democratic Republic of Congo: a systematic review and meta-analysis (1970–2024)"

Year of study

**Event rate (%)**

**Coinfection proportion (%)**

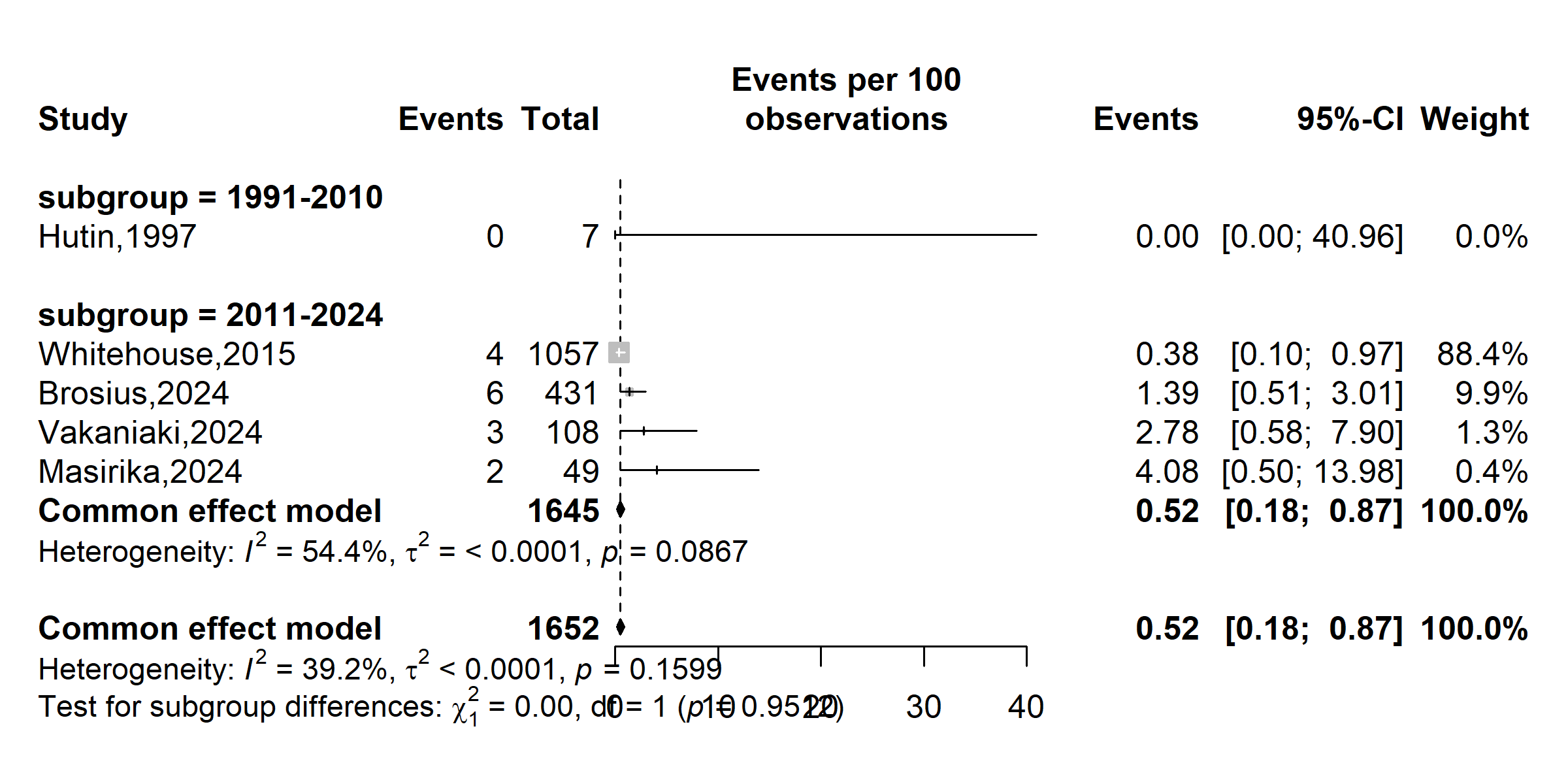

**Supplementary** **Fig. 1** Subgroup analysis of the HIV and Mpox coinfection in DRC, 1970-2024 *(based on policy or healthcare system changes: 1970–1990: Limited healthcare infrastructure in endemic regions; 1991-2010: Improvements in healthcare access and disease surveillance; 2011-2024: Strengthened global health initiatives and response systems)*

Study location

**Event rate (%)**

**Coinfection proportion (%)**

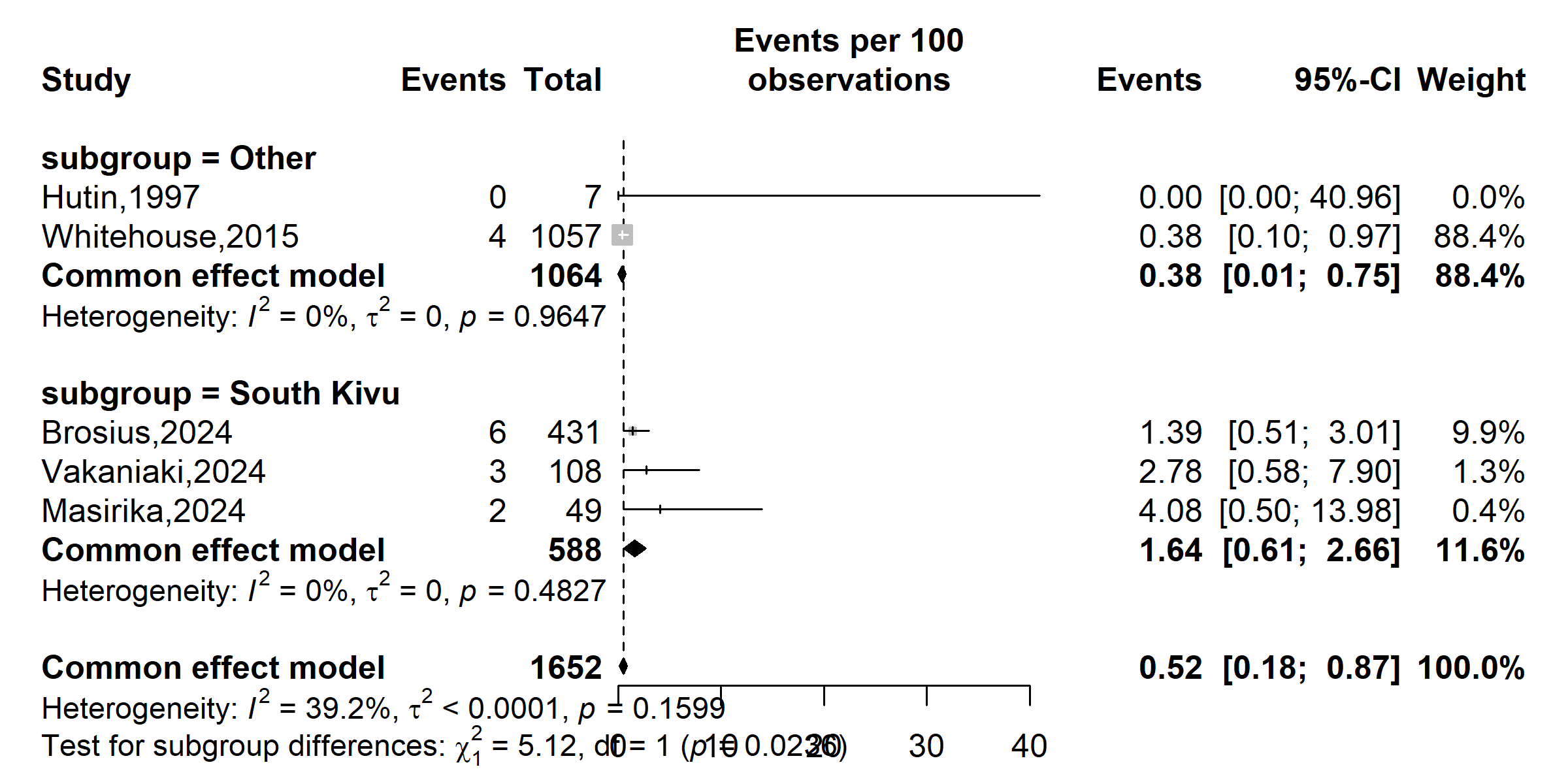

**Supplementary** **Fig. 2** Subgroup analysis of the HIV and Mpox coinfection in DRC, 1970-2024 *(based on geographical location: Other= Sankuru, Tshuapa)*

Study design

**Event rate (%)**

**Coinfection proportion (%)**

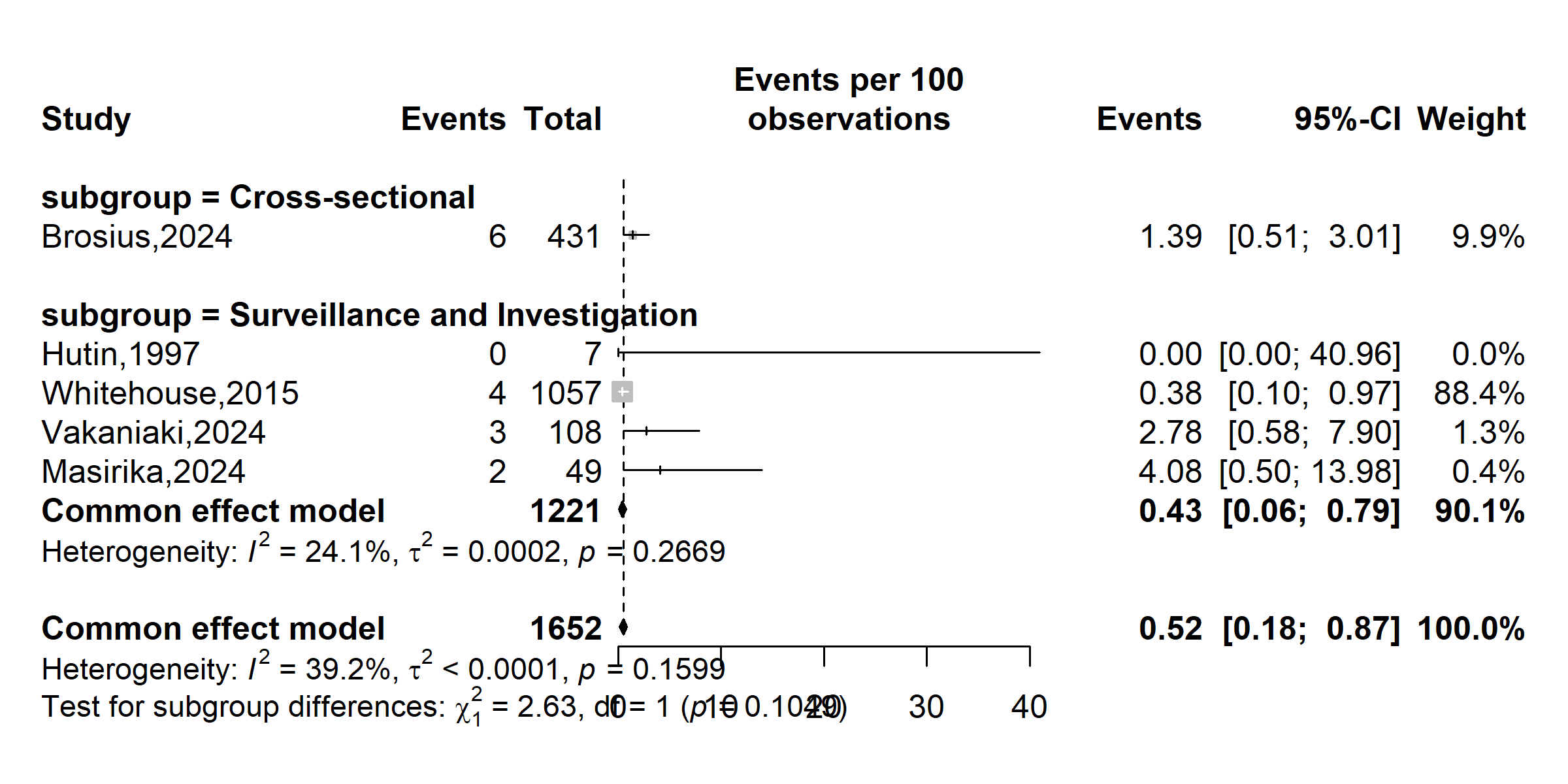

**Supplementary** **Fig. 3** Subgroup analysis of the HIV and Mpox coinfection in DRC, 1970-2024 *(based on study design)*

Study setting

**Event rate (%)**

**Coinfection proportion (%)**

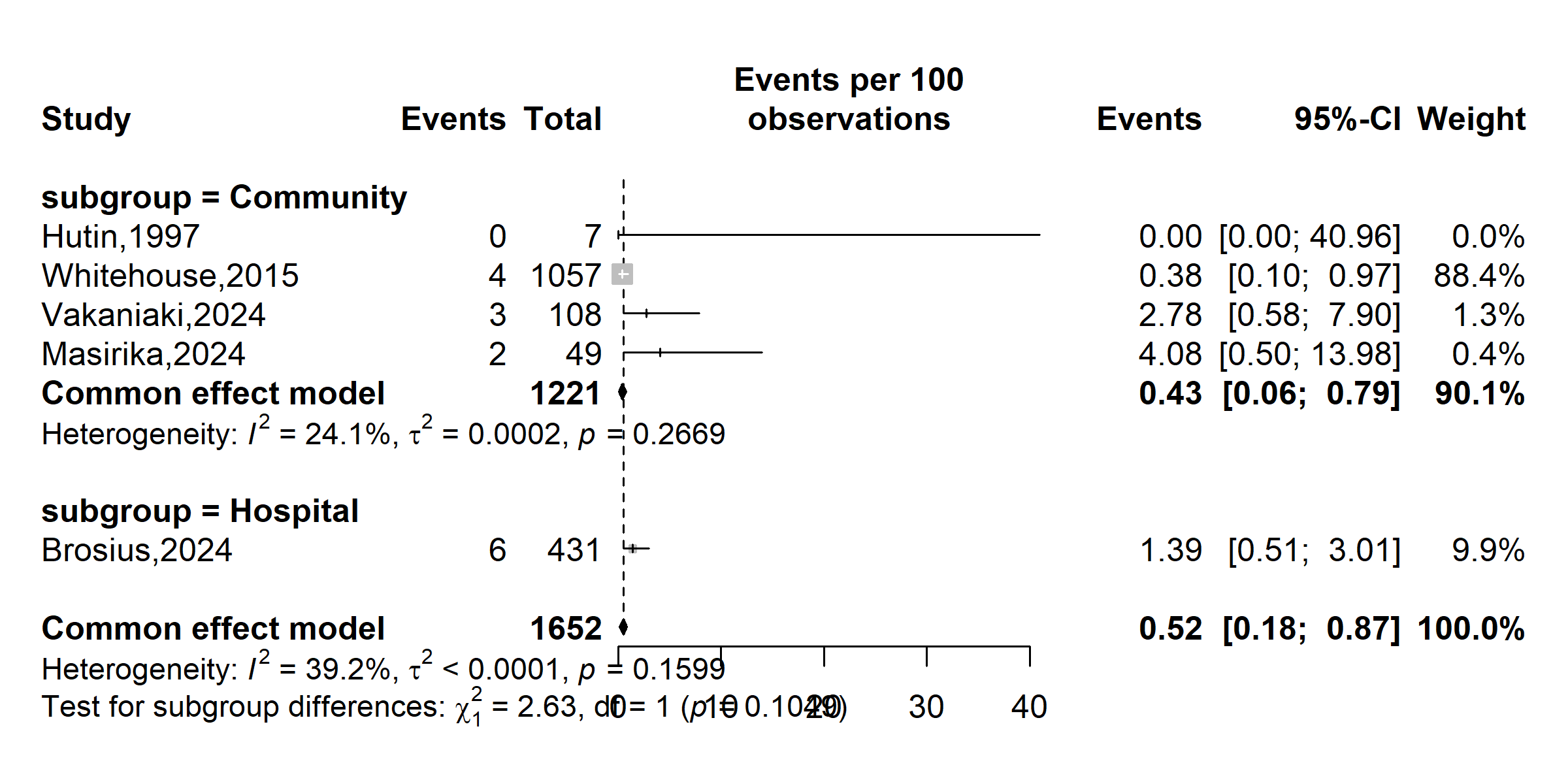

**Supplementary** **Fig. 4** Subgroup analysis of the HIV and Mpox coinfection in DRC, 1970-2024 *(based on study setting)*

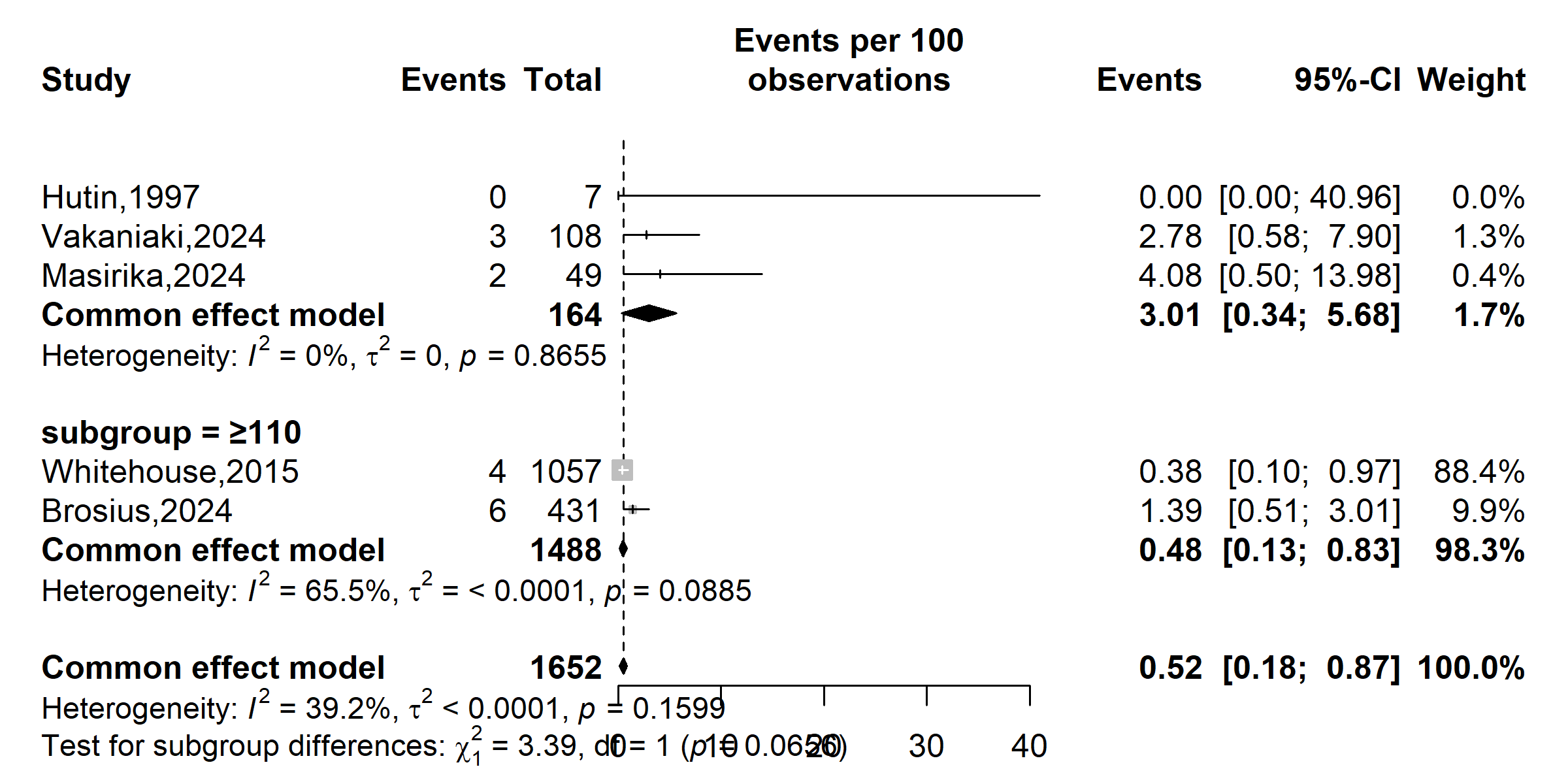

**Subgroup=˂110**

**Event rate (%)**

**Coinfection proportion (%)**

**Supplementary** **Fig. 5** Subgroup analysis of the HIV and Mpox coinfection in DRC, 1970-2024 *(based on study disease burden)*

### Publication bias assessment

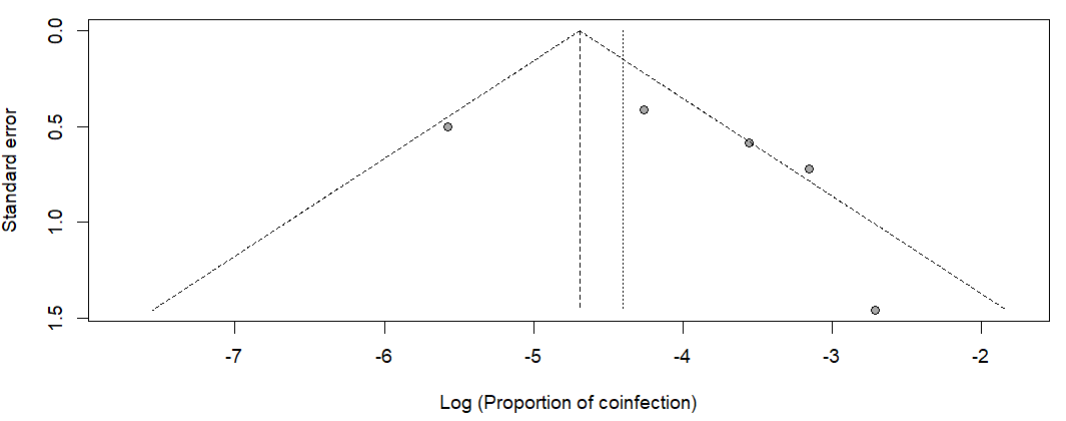

Supplementary Fig. 6 Funnel plot with pseudo 95% confidence limits and tests assessing the publication bias studies included

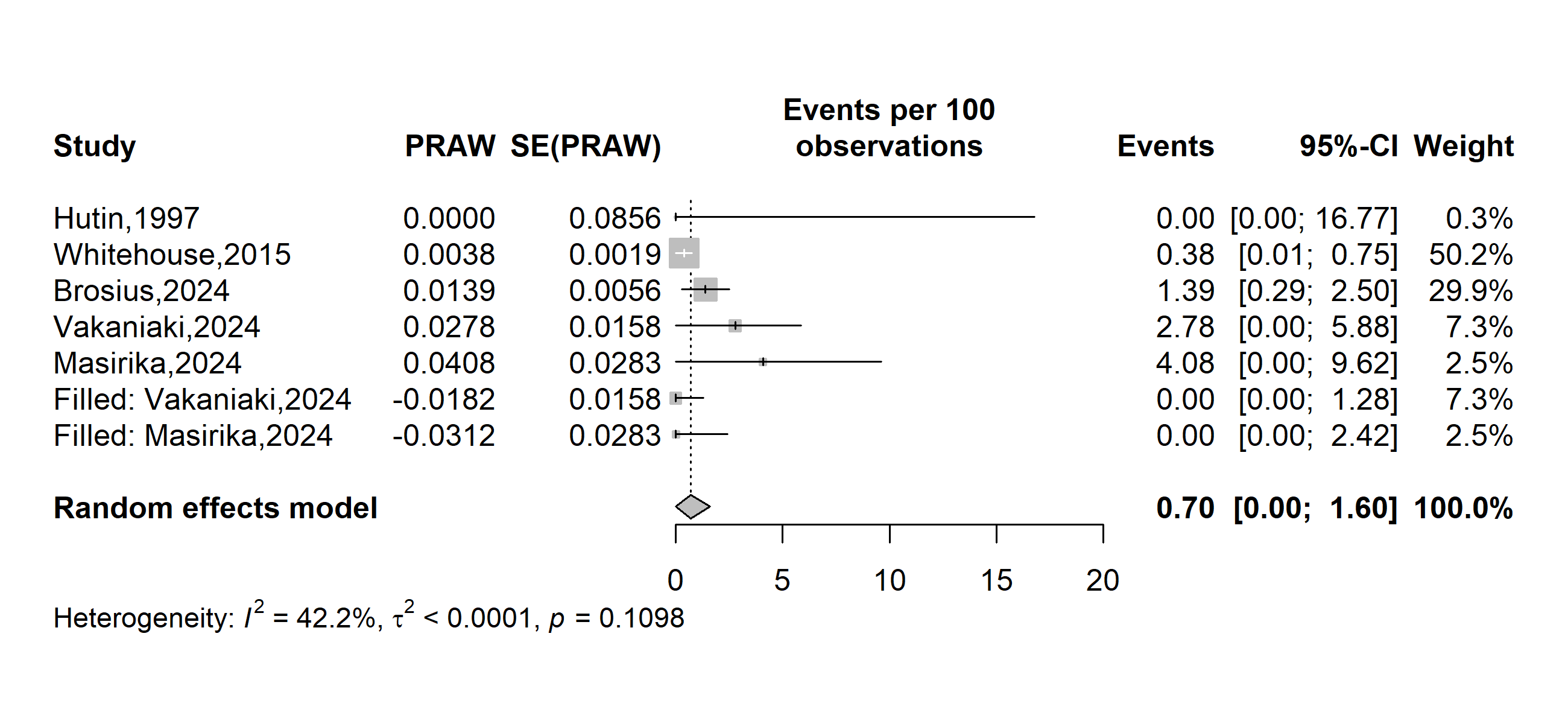

Supplementary Fig. 7 Funnel plot of the trim-and-fill method addressing publication bias

### Sensitivity test

**Coinfection proportion (%)**

**Event rate (%)**

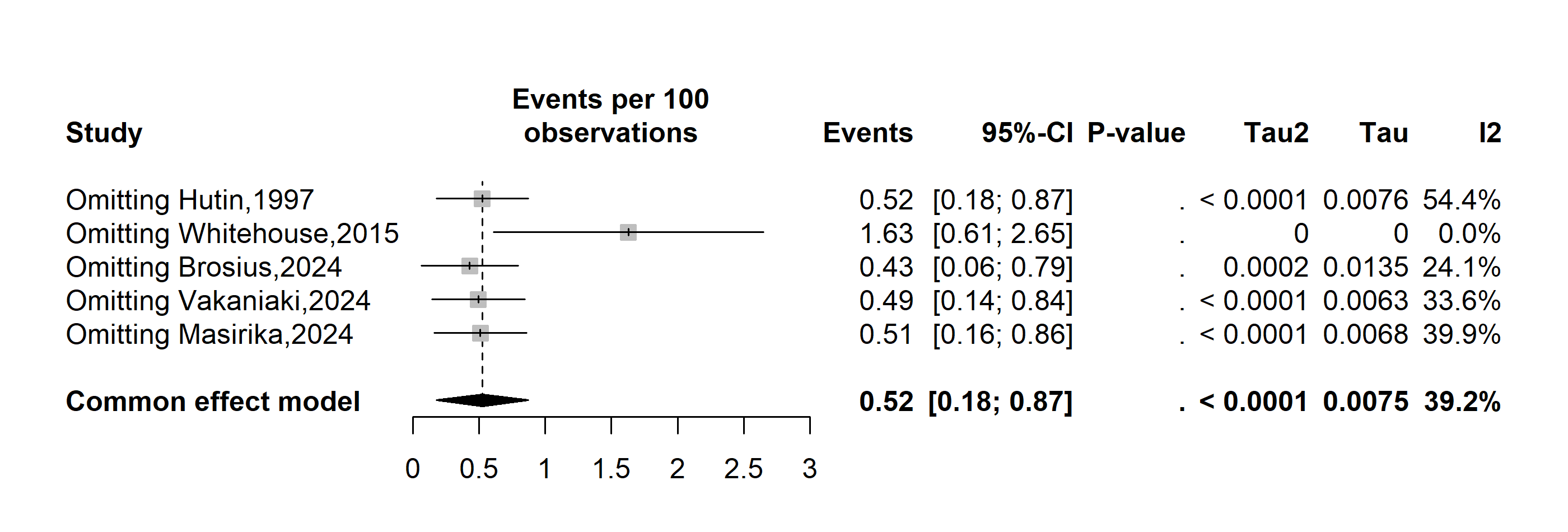

Supplementary Fig. 6 Sensitivity analysis of the pooled proportion estimate of varicella-zoster virus coinfection among confirmed Mpox cases in DRC, 1970-2024
