## Additional Files 5 for "Mpox clinical features and varicella-zoster virus coinfection in the Democratic Republic of Congo: a systematic review and meta-analysis (1970–2024)"

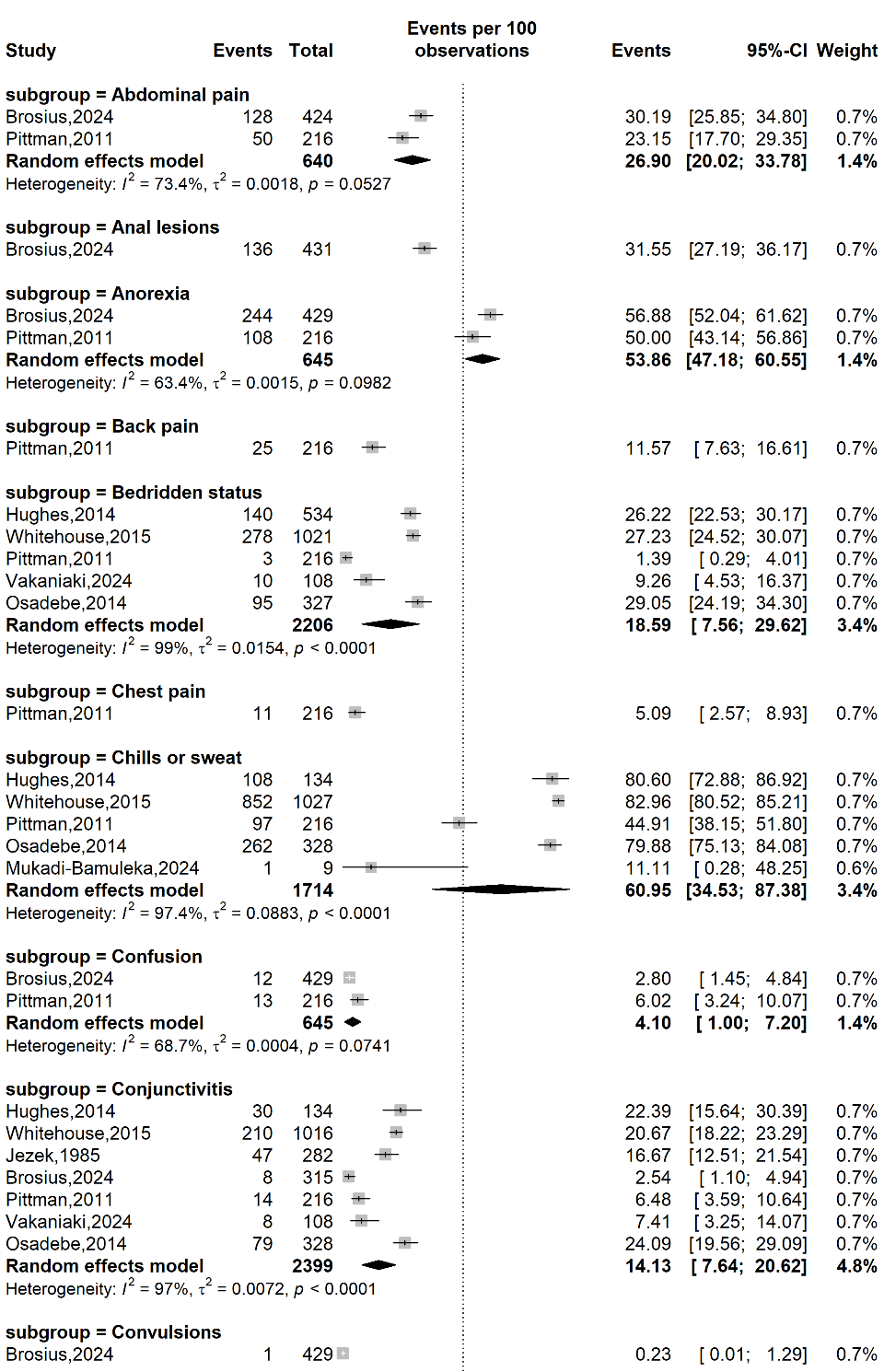

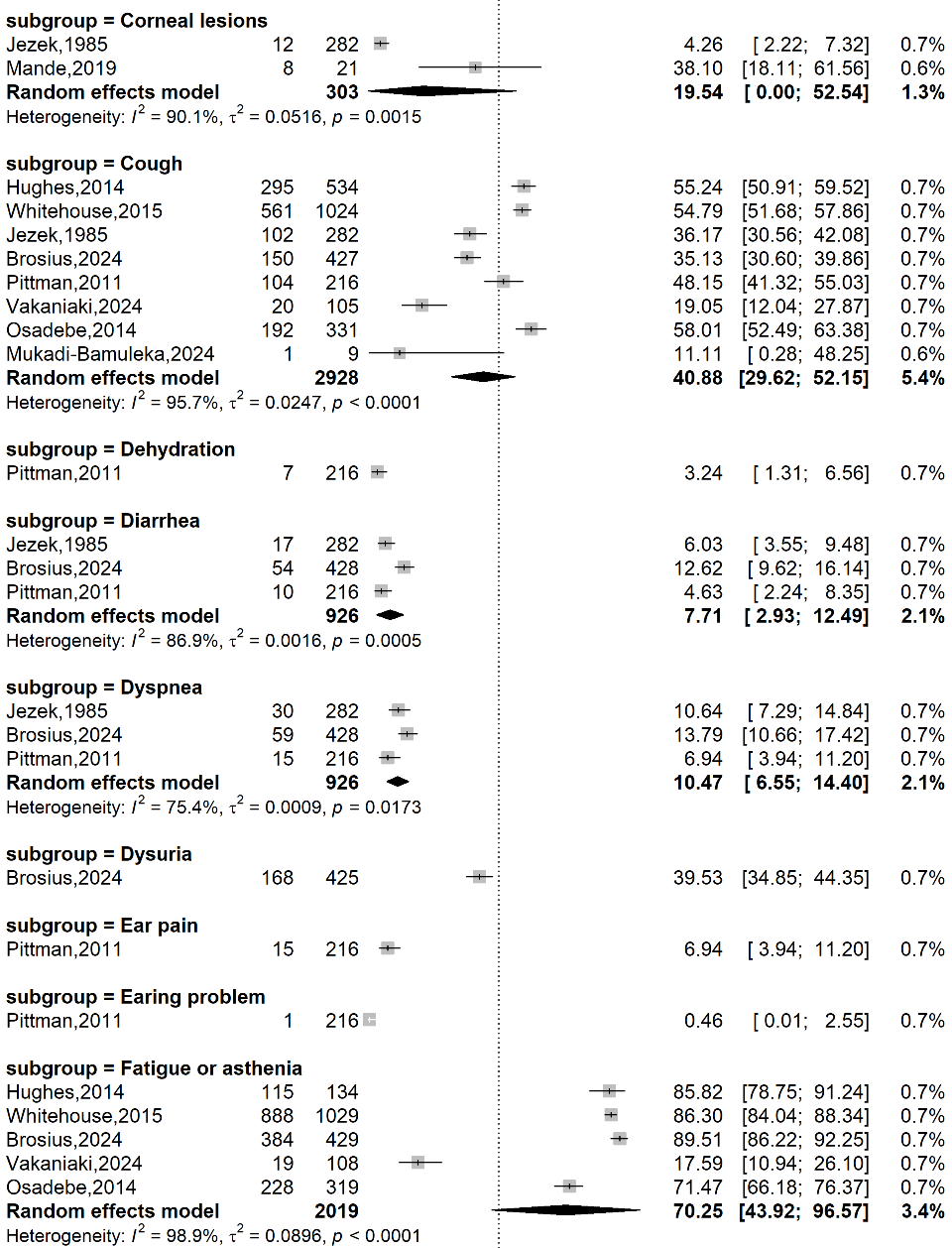

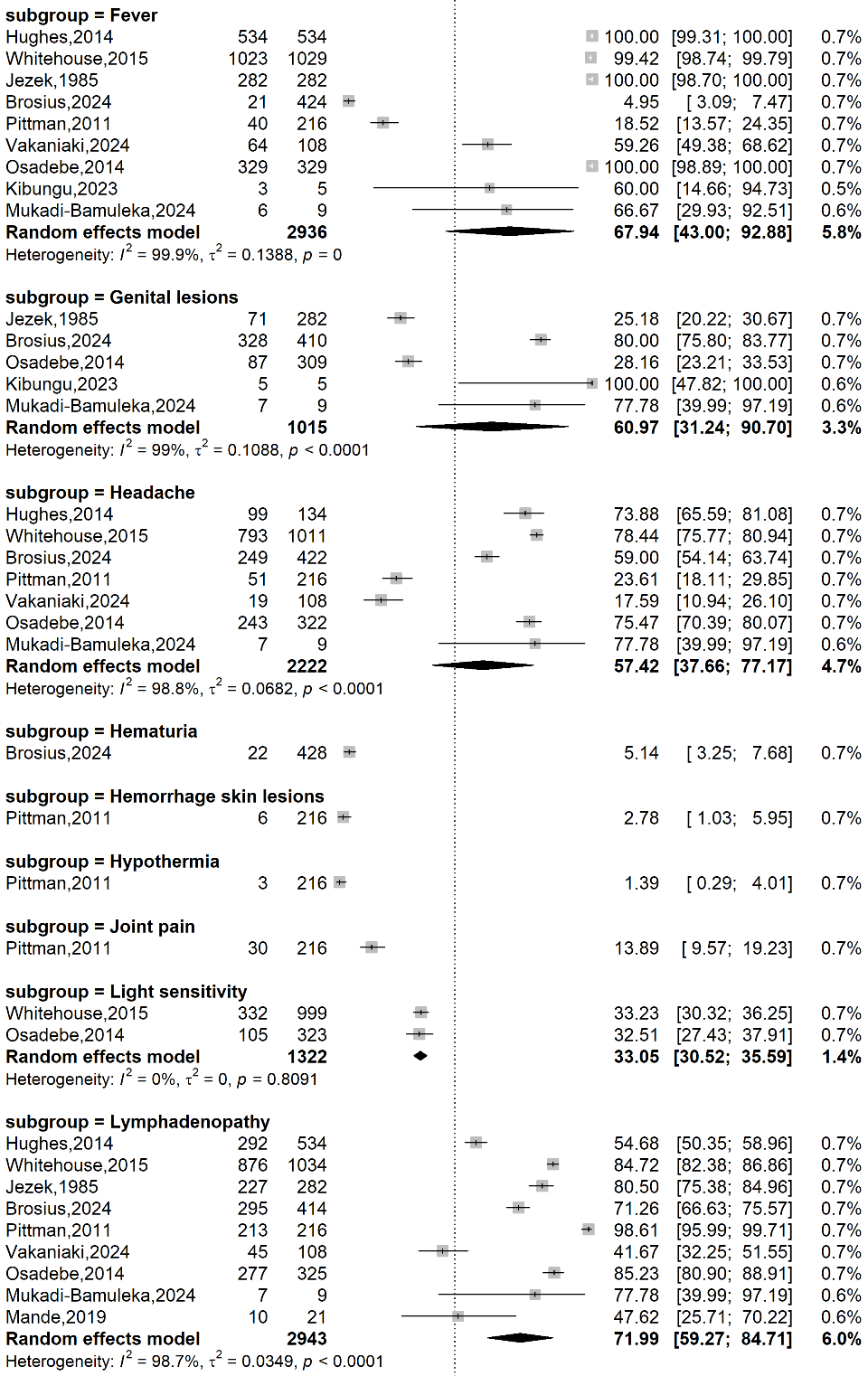

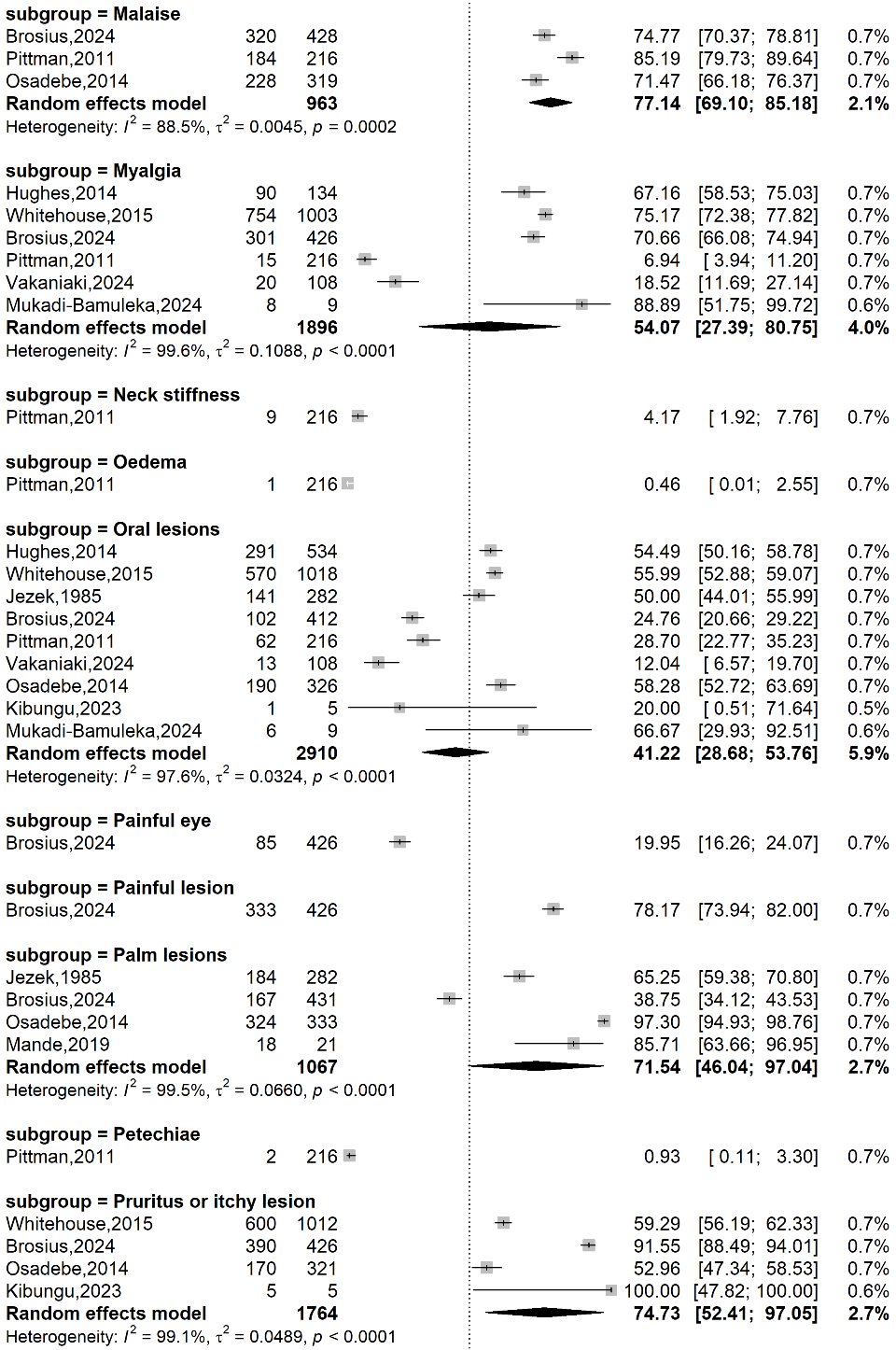

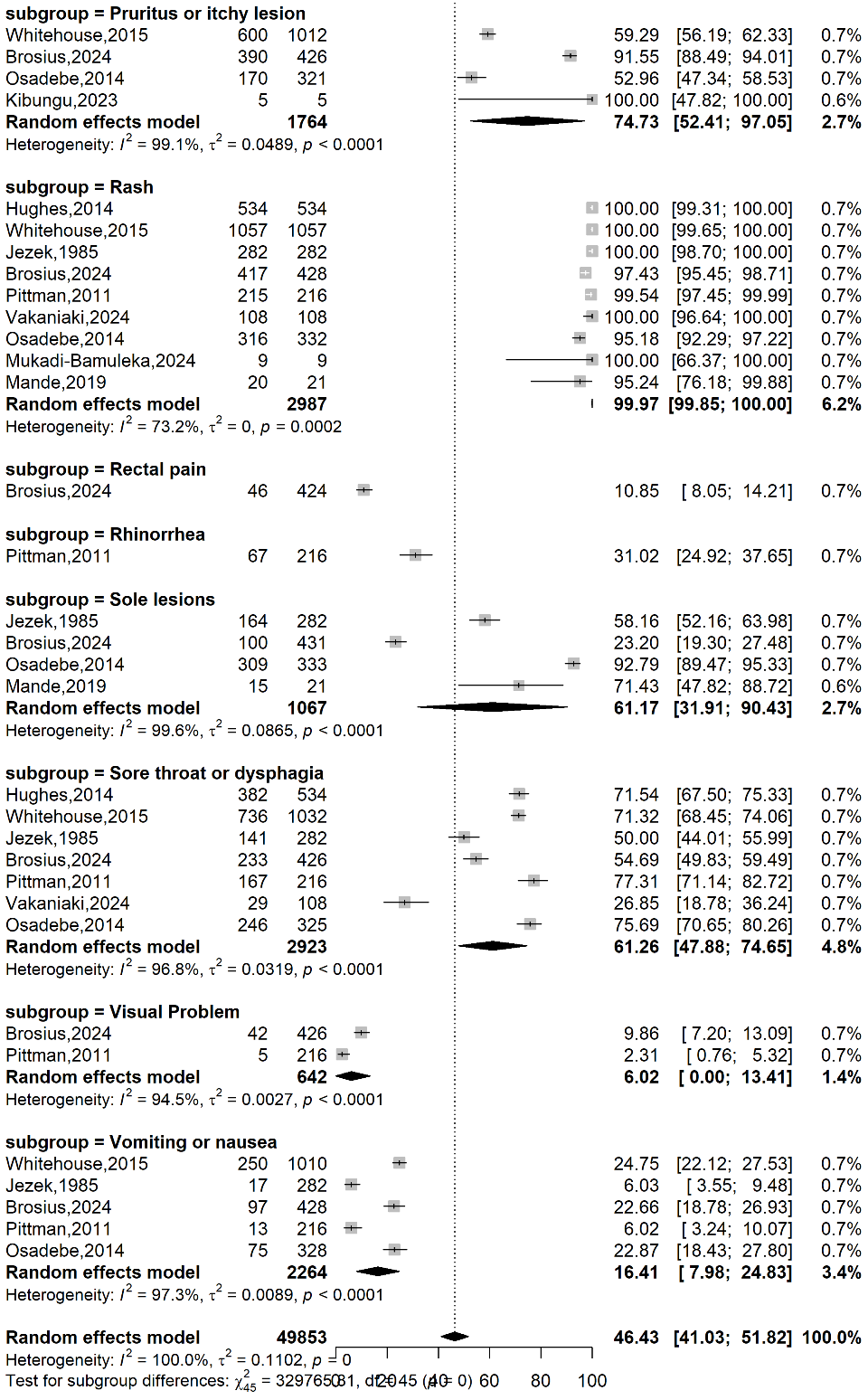

Supplementary Fig. 1 Clinical feature of confirmed Mpox cases in DRC, 1970-2024
