## Additional Files 6 for "Mpox clinical features and varicella-zoster virus coinfection in the Democratic Republic of Congo: a systematic review and meta-analysis (1970–2024)"

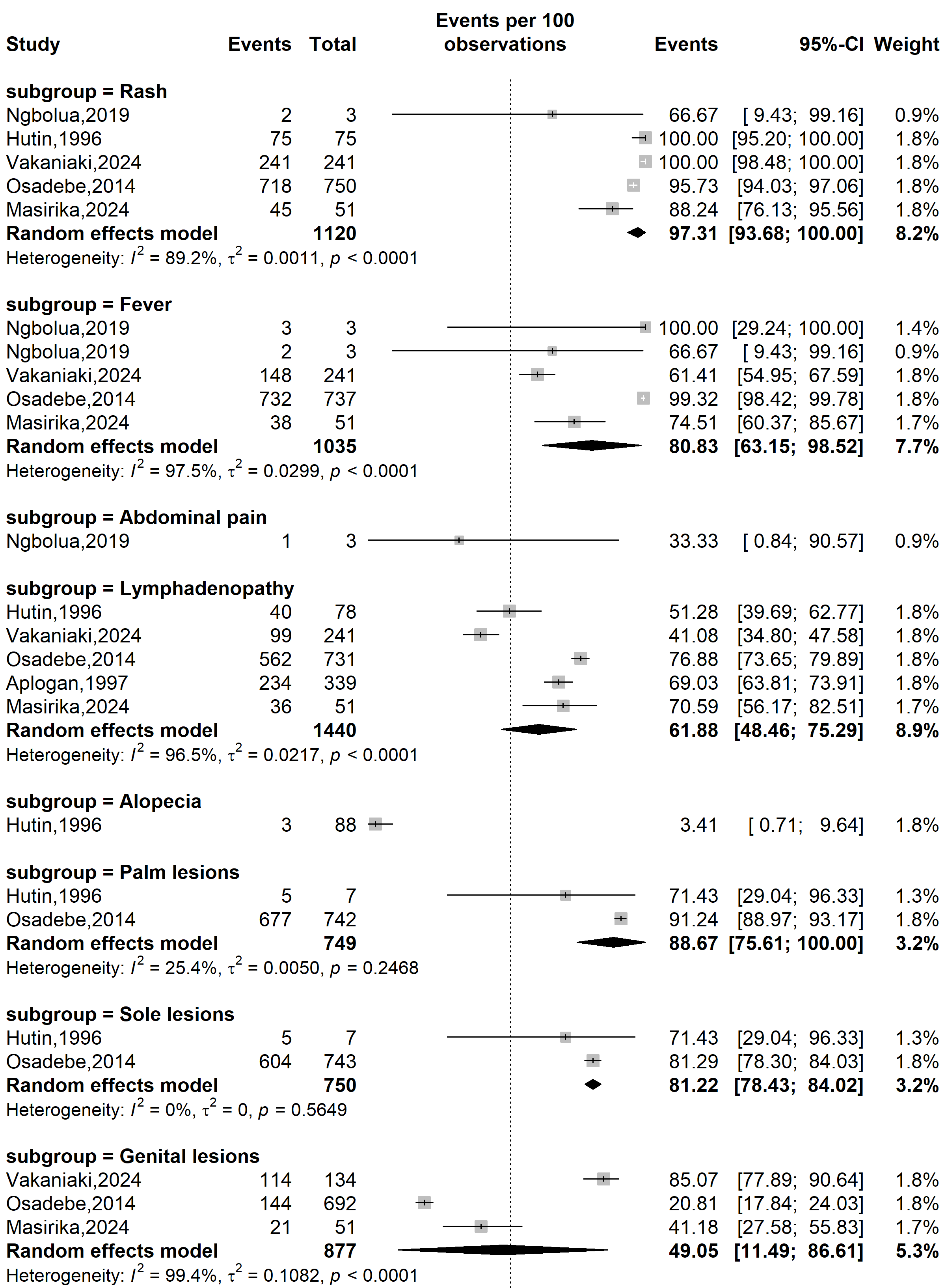

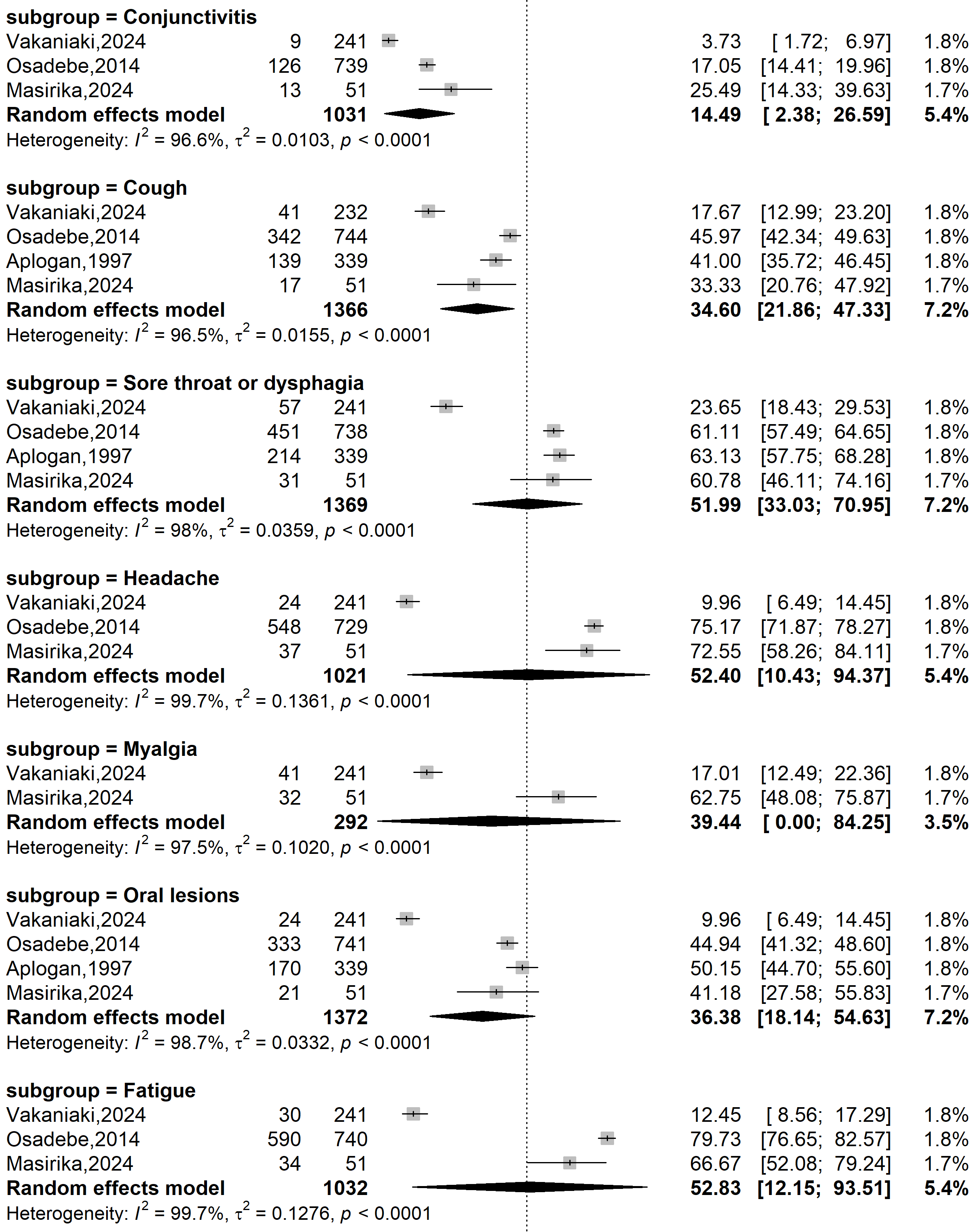

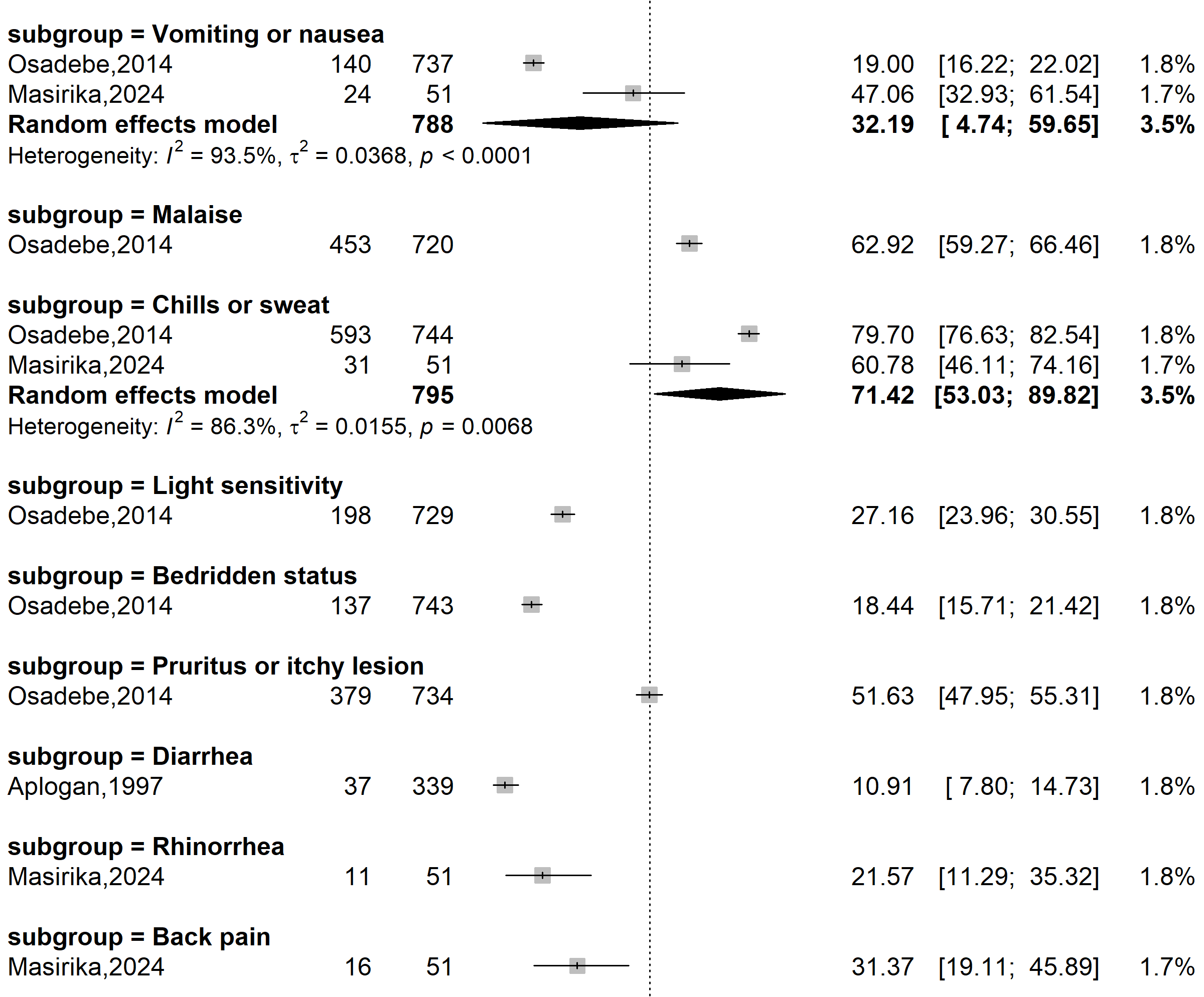

Supplementary Fig. 1 Clinical feature of suspected Mpox cases in DRC, 1970-2024
