## Additional Files 1 for "Mpox clinical features and varicella-zoster virus coinfection in the Democratic Republic of Congo: a systematic review and meta-analysis (1970–2024)"

Supplementary Table 1 Characteristic of studies assessing Mpox in DRC, 1970-2024

| **Author** | **Study**  **Year*^1^*** | **Region** | **Setting** | **Study population** | **Study type** | **Sampling** | **Risk of bias** | **Outcome of interest** | **VZV among**  **confirmed Mpox case** | **HIV among confirmed Mpox case** | **Summary of findings** |
| --- | --- | --- | --- | --- | --- | --- | --- | --- | --- | --- | --- |
| Jezek *et al.* [1] | 1985 | Nationwide | Community | General population | Surveillance and investigation report | Non-probabilistic | Low | Clinical feature | NR | NR | unvaccinated individuals, particularly young children, experienced an 11-15% case-fatality rate, while vaccinated individuals showed no deaths and altered disease presentation. |
| Hutin *et al.* [2] | 1997 | Sankuru | Community | General population | Surveillance and investigation report | Non-probabilistic | Low | Clinical feature | NR | Yes, *n*=0 | This outbreak presented with notable attack and case-fatality rates. The cessation of smallpox vaccination in 1983, following global eradication, contributed to an increased population susceptibility to monkeypox. |
| Aplogan *et al.* [3] | 1997 | Kasai Oriental | Community | General population | Surveillance and investigation report | Non-probabilistic | Low | VZV coinfection | No | NR | There was a high prevalence of Mpox among children and a mix of primary and secondary transmission patterns across numerous villages. The outbreak was localized outbreaks and travel, close  contact within households and neighborhoods facilitated the secondary case  transmission. |
| Meyer *et al.* [4] | 2001 | Equateur | Community | General population | Surveillance and investigation report | Non-probabilistic | Low | VZV coinfection | Yes*, n*=2 | NR | Two outbreaks were confirmed as monkeypox (4 deaths), two as co-infection of monkeypox and chickenpox (1 death), two as chickenpox (no deaths), and one outbreak yielded no viral evidence (no deaths). |
| Pittman *et al.* [5] | 2022 | Sankuru | Hospital | General population | Cross-sectional study | Non-probabilistic | Moderate | Clinical feature | NR | NR | Clinical course of Mpox in 216 PCR-confirmed cases revealed a 1.4% mortality rate and significant fetal loss in pregnant patients. Key findings included a high prevalence of rash and lymphadenopathy, with younger children exhibiting higher lesion counts and severe disease being associated with hypoalbuminemia and elevated viral load. |
| Nolen *et al.* [6] | 2012 | Tshuapa | Hospital | General population and healthcare worker | Surveillance and investigation report | Non-probabilistic | Low | VZV coinfection | Yes*, n*=0 | NR | Half of possible cases were lab-confirmed monkeypox. 50% household attack rate, multiple family transmissions, and a mean 8-day incubation period (4-14 days) were observed. |
| Hughes *et al.* [7] | 2014 | Tshuapa | Community | General population | Surveillance and investigation report | Non-probabilistic | Low | VZV coinfection and Clinical feature | Yes, *n*=134 | NR | A significant proportion of mpox cases were co-infected with VZV, exhibiting atypical clinical presentations and highlighting the complex interplay between these two viruses. |
| Petersen et al. [8] | 2014 | Tshuapa | Community | Healthcare worker | Surveillance and investigation report | Non-probabilistic | Low | VZV coinfection | Yes*, n*=1 | NR | These findings suggest that while a smallpox vaccination scar may offer some protection against monkeypox, healthcare workers remain at risk of infection. The presence of co-infections further complicates the picture. |
| Osadebe *et al.* [9] | 2014 | Tshuapa | Community | General population | Surveillance and investigation report | Non-probabilistic | Low | VZV coinfection and Clinical feature | No | NR | Laboratory-confirmed Mpox and VZV cases presented with many of the same signs and symptoms, and the analysis here emphasized the utility of including 12 specific signs/symptoms when investigating Mpox cases |
| McCollum *et al.* [10] | 2014 | Kivu (North and South) | Community | General population | Surveillance and investigation report | Non-probabilistic | Low | VZV coinfection | Yes, *n*=1 | NR | Report of the first confirmed cases of monkeypox (MPX) in forested areas of North and South Kivu Provinces, which aligns with ecological predictions for suitable Mpox transmission zones. |
| Whitehouse *et al.* [11] | 2014 | Tshuapa | Community | General population | Surveillance and investigation report | Non-probabilistic | Low | VZV coinfection and Clinical feature | Yes, *n*=169 | Yes, *n*=4 | Increased incidence compared to previous decades, likely due to waning smallpox immunity. While males generally had higher infection rates, females reported frequent contact with symptomatic individuals. Animal exposures were most common in males. |
| Mande *et al.* [12] | 2019 | Bas-Uélé | Community | General population | Surveillance and investigation report | Non-probabilistic | Low | VZV coinfection and Clinical feature | Yes, *n*=0 | NR | Among 77 suspected cases, PCR revealed 27.3% monkeypox, 58.4% chickenpox, and 14.3% negative. Monkeypox cases showed distinct skin lesions. |
| Ngbolua *et al.* [13] | 2019 | North Ubangui | Hospital | General population | Surveillance and investigation report | Non-probabilistic | High | Clinical feature | NR | NR | Three cases of monkeypox in young males with similar clinical presentations, including fever, rash, itching, and abdominal pain. No case of death. |
| Kinganda-Lusamaki *et al.* [14] | 2022 | Nationwide | Community and Hospital | General population | Surveillance and investigation report | Non-probabilistic | Low | VZV coinfection and Clinical feature | Yes, *n*=0 | NR | Adding antibody testing to PCR nearly doubled Mpox detection rates in the DRC (48% vs 34% with PCR alone), revealing hidden outbreaks across 14 additional health zones and proving current surveillance misses over 40% of cases. |
| Kibungu *et al.* [15] | 2023 | Kwango | Community | General population | Surveillance and investigation report | Non-probabilistic | Low | Clinical feature | NR | NR | A cluster of clades I monkeypox cases in the DRC shows sexual transmission, indicating this route is not limited to clade IIb. |
| Bangwen *et al.* [16] | 2023 | Nationwide | Community and Hospital | General population | Surveillance and investigation report | Non-probabilistic | Low | VZV coinfection | No | NR | There was a four-fold increase in incidence between 2010 and 2023, wider geographic spread, and high fatality rate in young children. |
| Brosius *et al.* [17] | 2024 | South Kivu | Hospital | General population | Cross-sectional study | Non-probabilistic | Moderate | Clinical feature | NR | Yes, *n*=6 | Most suspected cases were PCR-positive for Mpox. Most cases reported contact with known Mpox, primarily spouses/partners in adults and family in children. Genital lesions were common in adults. Hospitalized mortality was low. |
| Mukadi-Bamuleka e*t al.* [18] | 2024 | North Kivu | Community | General population | Surveillance and investigation report | Non-probabilistic | Low | Clinical feature | NR | NR | Clade Ib monkeypox was introduced into North Kivu, including displacement camps, with suspected non-intimate contact transmission, affecting children. |
| Vakaniaki *et al*. [19] | 2024 | South Kivu | Community | General population | Surveillance and investigation report | Non-probabilistic | Low | Clinical feature | NR | Yes, *n*=3 | The Mpox outbreak in eastern DRC was caused by a distinct Clade I Mpox lineage, differing from historical zoonotic patterns. The outbreak, predominantly affected young adults including a significant proportion of female sex workers, suggests a shift towards human-to-human transmission, potentially involving sexual contact. |
| Masirika *et al.* [20] | 2024 | South Kivu | Community | General population | Surveillance and investigation report | Non-probabilistic | Low | Clinical feature | NR | Yes, *n*=2 | The findings suggest heterosexual close contact as the main transmission route, highlighting the increased risk for sex workers and their clients in this region. |
| *^1^* Date of study completion; VZV: Varicella-Zoster Virus; Surveillance and Investigation Report (Case reports and Case series); NR: Not Reported; DRC: Democratic Republic of Congo; PCR: Polymeras Chain Reaction | | | | | | | | | | | |

Supplementary Table 2 Searching Strategies for online databases

| Database | Search Term |
| --- | --- |
| PubMed | (monkeypox) OR (monkeypox virus) OR (human monkeypox) OR (Mpox) OR (mpox) OR (MPX) OR (epidemiology) OR (surveillance) OR (varicella-zoster virus) OR (HIV) (characteristics) OR (clinical characteristics) OR (severe) AND (DRC) OR (Democratic Republic of Congo) OR (Zaire) |
| Google Scholar | (monkeypox) OR (monkeypox virus) OR (human monkeypox) OR (Mpox) OR (mpox) OR (MPX) OR (epidemiology) OR (surveillance) OR (varicella-zoster virus) OR (HIV) OR (characteristics) OR (clinical characteristics) OR (severe) AND (DRC) OR (Democratic Republic of Congo) OR (Zaire) |
| ScienceDirect | (monkeypox) OR (monkeypox virus) OR (human monkeypox) OR (Mpox) OR (mpox) OR (MPX) OR (epidemiology) OR (surveillance) OR (varicella-zoster virus) OR (HIV) OR (characteristics) OR (clinical characteristics) OR (severe)) AND (DRC) OR (Democratic Republic of Congo) OR (Zaire) |

**References**

1.	Jezek Z, Szczeniowski M, Paluku KM, Mutombo M. Human Monkeypox: Clinical Features of 282 Patients. *Journal of Infectious Diseases.* 1987;156(2):293–8.

2.	Hutin YJF, Williams RJ, Malfait P, Pebody R, Loparev VN, Ropp SL, *et al.* Outbreak of Human Monkeypox, Democratic Republic of Congo, 1996 to 1997. *Emerg Infect Dis*. 2001;7(3):434–8.

3.	Aplogan A, Mangindula V, Muamba P, Mwema G, Okito L, Pebody R, *et al*. Human monkeypox -- Kasai Oriental, Democratic Republic of Congo, February 1996-October 1997. *MMWR Morb Mortal Wkly Rep*. 1997;46(49):1168–71.

4.	Meyer H, Perrichot M, Stemmler M, Emmerich P, Schmitz H, Varaine F, *et al.* Outbreaks of Disease Suspected of Being Due to Human Monkeypox Virus Infection in the Democratic Republic of Congo in 2001. *J Clin Microbiol.* 2002;40(8):2919–21.

5.	Pittman PR, Martin JW, Kingebeni PM, Tamfum JJM, Mwema G, Wan Q, *et al.* Clinical characterization and placental pathology of mpox infection in hospitalized patients in the Democratic Republic of the Congo. Bowman N, editor. *PLoS Negl Trop Dis*. 2023;17(4): e0010384.

6.	Nolen LD, Osadebe L, Katomba J, Likofata J, Mukadi D, Monroe B, *et al.* Extended Human-to-Human Transmission during a Monkeypox Outbreak in the Democratic Republic of the Congo. *Emerg Infect Dis.* 2016;22(6):1014–21.

7.	Hughes CM, Liu L, Davidson WB, Radford KW, Wilkins K, Monroe B, *et al.* A Tale of Two Viruses: Coinfections of Monkeypox and Varicella Zoster Virus in the Democratic Republic of Congo. *The American Journal of Tropical Medicine and Hygiene*. 2021;104(2):604–11.

8.	Petersen BW, Kabamba J, McCollum AM, Lushima RS, Wemakoy EO, Muyembe Tamfum JJ, *et al.* Vaccinating against monkeypox in the Democratic Republic of the Congo. *Antiviral Research.* 2019; 162:171–7.

9.	Osadebe L, Hughes CM, Shongo Lushima R, Kabamba J, Nguete B, Malekani J, *et al*. Enhancing case definitions for surveillance of human monkeypox in the Democratic Republic of Congo. Kasper M, editor. *PLoS Negl Trop Dis*. 2017;11(9): e0005857.

10.	McCollum AM, Nakazawa Y, Ndongala GM, Pukuta E, Karhemere S, Lushima RS, *et al.* Human Monkeypox in the Kivus, a Conflict Region of the Democratic Republic of the Congo. *The American Society of Tropical Medicine and Hygiene.* 2015;93(4):718–1.

11.	Whitehouse ER, Bonwitt J, Hughes CM, Lushima RS, Likafi T, Nguete B, *et al.* Clinical and Epidemiological Findings from Enhanced Monkeypox Surveillance in Tshuapa Province, Democratic Republic of the Congo During 2011–2015. *The Journal of Infectious Diseases*. 2021;223(11):1870–8.

12.	Mande G, Akonda I, Weggheleire AD, Brosius I, Liesenborghs L, Bottieau E, *et al.* Enhanced surveillance of monkeypox in Bas-Uélé, Democratic Republic of Congo: the limitations of symptom-based case definitions. *International Journal of Infectious Diseases*. 2022; 122:647–55.

13.	Koto-te-Nyiwa Ngbolua, Guy Kumbali Ngambika, Blaise Mbembo-wa-Mbembo, Kohowe Pagerezo Séraphin, Kogana Kapalata Fabrice, Gédéon Ngiala Bongo, *et al*. First Report on Three Cases of Monkey pox in Nord Ubangi Province (Democratic Republic of the Congo). *bioex*. 2020;2(1):120–5.

14.	Kinganda-Lusamaki E, Baketana LK, Ndomba-Mukanya E, Bouillin J, Thaurignac G, Aziza AA, *et al*. Use of Mpox Multiplex Serology in the Identification of Cases and Outbreak Investigations in the Democratic Republic of the Congo (DRC). *Pathogens*. 2023;12(7):916.

15.	Kibungu EM, Vakaniaki EH, Kinganda-Lusamaki E, Kalonji-Mukendi T, Pukuta E, Hoff NA, *et al.* Clade I–Associated Mpox Cases Associated with Sexual Contact, the Democratic Republic of the Congo. *Emerg Infect Dis.* 2024 ;30(1):172–6.

16.	Bangwen E, Diavita R, Vos ED, Vakaniaki EH, Nundu SS, Mutombo A, *et al.* Suspected and confirmed mpox cases in DR Congo: a retrospective analysis of national epidemiological and laboratory surveillance data, 2010–23. *The Lancet.* 2025;405(10476):408–19.

17.	Brosius I, Vakaniaki EH, Mukari G, Munganga P, Tshomba JC, Vos ED, *et al.* Epidemiological and clinical features of mpox during the clade Ib outbreak in South Kivu, Democratic Republic of the Congo: a prospective cohort study. *The Lancet.* 2025;405(10478):547–59.

18.	Mukadi-Bamuleka D, Kinganda-Lusamaki E, Mulopo-Mukanya N, Amuri-Aziza A, O’Toole Á, Modadra-Madakpa B, *et al*. First imported Cases of MPXV Clade Ib in Goma, Democratic Republic of the Congo: Implications for Global Surveillance and Transmission Dynamics. *medRxiv*. 2024. doi: 10.1101/2024.09.12.24313188

19.	Vakaniaki EH, Kacita C, Kinganda-Lusamaki E, O’Toole Á, Wawina-Bokalanga T, Mukadi-Bamuleka D, *et al.* Sustained human outbreak of a new MPXV clade I lineage in eastern Democratic Republic of the Congo. *Nat Med.* 2024;30(10):2791–5.

20.	Masirika LM, Udahemuka JC, Ndishimye P, Martinez GS, Kelvin P, Nadine MB, *et al.* Epidemiology, clinical characteristics, and transmission patterns of a novel Mpox (Monkeypox) outbreak in eastern Democratic Republic of the Congo (DRC): an observational, cross-sectional cohort study. *medRxiv*; 2024. doi: 10.1101/2024.03.05.24303395v1.
